# Ultrasound and biochemical predictors of pregnancy outcome at diagnosis of early-onset fetal growth restriction

**DOI:** 10.1101/2023.01.27.23285087

**Authors:** Rebecca Spencer, Kasia Maksym, Kurt Hecher, Karel Maršál, Francesc Figueras, Gareth Ambler, Harry Whitwell, Nuno Rocha Nené, Neil J. Sebire, Stefan R. Hansson, Anke Diemert, Jana Brodszki, Eduard Gratacós, Yuval Ginsberg, Tal Weissbach, Donald M Peebles, Ian Zachary, Neil Marlow, Angela Huertas-Ceballos, Anna L. David

**Affiliations:** UCL Elizabeth Garrett Anderson Institute for Women’s Health, University College London, London, UK; Leeds Institute of Cardiovascular and Metabolic Medicine, University of Leeds, Leeds, UK; Department of Obstetrics and Fetal Medicine, University Medical Center Hamburg-Eppendorf, Hamburg, Germany; Department of Obstetrics and Gynaecology, Institute of of Clinical Sciences Lund, Skane University Hospital, Lund University, Lund, Sweden; Institut D’Investigacions Biomèdiques August Pi í Sunyer, University of Barcelona, Barcelona Center for Maternal-Fetal and Neonatal Medicine, Barcelona, Spain; Department of Statistical Science, University College London, London, UK; National Phenome Centre and Imperial Clinical Phenotyping Centre, Department of Metabolism, Digestion and Reproduction, Imperial College London, London, UK; Section of Bioanalytical Chemistry, Division of Systems Medicine, Department of Metabolism, Digestion and Reproduction, Imperial College London, London, UK; Population, Policy & Practice Department, Great Ormond Street Institute of Child Health, University College London, London, UK; Department of Obstetrics and Gynecology, Rambam Medical Centre, Haifa, Israel; Department of Obstetrics and Gynecology, Sheba Medical Center Tel Hashomer, Tel Aviv, Israel; Division of Medicine, Faculty of Medical Sciences, University College London, UK; Neonatal Department, University College London Hospitals NHS Foundation Trust, London, UK

## Abstract

**Background:** Severe early-onset fetal growth restriction (FGR) causes significant fetal and neonatal mortality and morbidity. Predicting the outcome of affected pregnancies at the time of diagnosis is difficult, preventing accurate patient counselling. We investigated the use of maternal serum protein and ultrasound measures at diagnosis to predict fetal or neonatal death and three secondary outcomes: fetal death or delivery ≤28+0 weeks; development of abnormal umbilical artery Doppler velocimetry; slow fetal growth.

**Methods:** Women with singleton pregnancies (n=142, estimated fetal weights [EFWs] <3^rd^ centile, <600g 20+0-26+6 weeks of gestation, no known chromosomal, genetic or major structural abnormalities), were recruited from four European centres. Maternal serum from the discovery set (n=63) was analysed for seven proteins linked to angiogenesis, 90 additional proteins associated with cardiovascular disease and five proteins identified through pooled liquid chromatography tandem mass spectrometry. Patient and clinician stakeholder priorities were used to select models tested in the validation set (n=60), with final models calculated from combined data.

**Results:** The most discriminative model for fetal or neonatal death included EFW z-score (Hadlock 3 formula/Marsal chart), gestational age and umbilical artery Doppler category (AUC 0.91, 95%CI 0.86-0.97) but was less well calibrated than the model containing only EFW z-score (Hadlock3/Marsal). The most discriminative model for fetal death or delivery ≤28+0 weeks included maternal serum placental growth factor (PlGF) concentration and umbilical artery Doppler category (AUC 0.89, 95%CI 0.83-0.94).

**Conclusion:** Ultrasound measurements and maternal serum PlGF concentration at diagnosis of severe early-onset FGR predict pregnancy outcomes of importance to patients and clinicians.

**Trial registration:** ClinicalTrials.gov NCT02097667

**Funding:** European Union, Rosetrees Trust, Mitchell Charitable Trust.

## Introduction

The survival and growth of a fetus depends on placental provision of nutrients and waste exchange with the mother. When this system is impaired, by inadequate transformation of the uteroplacental circulation or deficits in the structure or function of the placenta, the fetus fails to reach their growth potential (1, 2). The resulting fetal growth restriction (FGR) may be diagnosed antenatally on the basis of an ultrasound determined low estimated fetal weight (EFW) for gestational age; either <3^rd^ centile or <10^th^ centile with abnormal Doppler ultrasound indices in the uterine (UtA) and/or umbilical (UmA) arteries (3) (4, 5). Early-onset FGR, occurring before 32 weeks of gestation, carries significant risks of stillbirth, neonatal morbidity and mortality, neurodevelopmental impairment, and long-term health problems (6-13). There is currently no treatment that can improve fetal growth in utero; instead, management involves monitoring the pregnancy and timing delivery to balance the risks of stillbirth and prematurity (4, 14-16).

An important question when developing novel therapies for early-onset FGR is which pregnancies to include in early-phase clinical trials. There is a balance to be struck between identifying pregnancies that are sufficiently severely affected to justify the possible risks of the intervention but not so severely affected that there is no potential to determine efficacy. Numerous studies have investigated predictive markers for the development of FGR (17-22), but far fewer have studied the prediction of pregnancy outcome when early-onset FGR is diagnosed. Lack of knowledge about pregnancy outcome in FGR makes it difficult to optimise the inclusion criteria for clinical trials, but it also leaves pregnant patients and their partners with a considerable burden of uncertainty (23, 24).

The EVERREST Project aims to carry out a phase I/IIa trial of maternal vascular endothelial growth factor (VEGF) gene therapy for early-onset FGR (25). The greatest potential for benefit is in pregnancies at the threshold of viability, for whom our current management option, preterm delivery, is not possible or is very high risk. In preparation for a clinical trial of a novel therapeutic, we established a multicentre prospective study to characterise the natural history of early-onset FGR, choosing an extreme phenotype in which the estimated fetal weight (EFW) was <3^rd^ centile and <600g between 20+0 and 26+6 weeks of gestation (henceforth referred to as ‘severe’ early-onset FGR) (26).

The aim of this work was to prospectively identify and validate ultrasound and serum biochemical factors that could be used to predict fetal or neonatal death in pregnancies affected by severe early-onset FGR. These could subsequently be used to select the most appropriate women for a first-in-human study of a novel therapeutic to treat FGR, and to better counsel women and their partners about pregnancy outcome. To this end, we asked patients and clinicians to assess the value of our primary and secondary outcomes, based on which we then selected models for validation. Unsupervised parenclitic network analysis by pregnancy outcome and functional network analysis of proteins associated with pregnancy outcomes were also performed to maximise the utility of the proteomic data with the aim of providing insights into the underlying pathophysiology.

## Results

The discovery set, recruited between March 2014 and September 2016, comprised 63 pregnant participants (Figure 1 & Table 1; supplementary data Table 1). Follow-up for the ascertainment of study outcomes was completed in November 2016. The validation set, recruited between October 2016 and January 2020, comprised 60 pregnant participants, with follow-up for the ascertainment of study outcomes completed in March 2020. There were no significant differences in maternal demographics, pregnancies characteristics or pregnancy outcomes between the discovery and validation sets (Table 1; supplementary data Table 2 & Figure 1). Overall, 42 (34%) of the pregnancies ended in the primary outcome of fetal or neonatal death (within the first 28 days of life). For the three secondary outcomes, only fetal death or delivery ≤28+0 weeks of gestation could be ascertained for all pregnancies, occurring in 58 cases (47%). The UmA Doppler velocimetry was normal (<95^th^ centile for gestation (27)) at enrolment in 46 participants, of whom 21 (46%) subsequently developed abnormal UmA Doppler measurements. Fetal growth trajectory (based on change in percentage weight deviation over a period or two weeks or more (28)) could be assessed for 104 pregnancies (85%), with the remaining pregnancies ending in fetal death or delivery before a 2-week interval was reached. Forty-one of these 104 fetuses (39%) demonstrated slow fetal growth (worsening of weight deviation of >10 percentage points). A smaller proportion of fetuses demonstrated slow fetal growth in the validation set (31%) than in the discovery set (47%).

**Table 1:** Characteristics and outcomes of the discovery, validation and combined participant sets. Discovery and validation sets were compared using 2-sided t tests for symmetrical continuous variables, Mann-Whitney U tests for skewed continuous variables and chi square or Fisher’s exact tests (where specified) for categorical variables.

|  | Discovery<br>n=63 | Validation<br>n=60 | p value | Combined<br>n=123 |
| --- | --- | --- | --- | --- |
| Maternal characteristics |  |  |  |  |
| Maternal age, mean years (SD) | 33.7 (6.0) | 33.3 (6.7) | 0.72 | 33.5 (6.3) |
| Primiparous, n (%) | 43 (68) | 37 (62) | 0.44 | 80 (65) |
| BMI, median (IQR) | 24.9 (22.7-28.6) | 26.4 (22.8-31.0) | 0.36 | 25.7 (22.8-30.0) |
| Ethnicity, n (%)<sup>1</sup> |  |  |  |  |
| White | 42 (67) | 27 (46) | 0.16 | 69 (57) |
| Black | 10 (16) | 15 (25) |  | 25 (20) |
| Asian | 10 (16) | 15 (25) |  | 25 (20) |
| Other | 1 (2) | 2 (3) |  | 3 (2) |
| Essential hypertension, n (%) | 10 (16) | 6 (10) | 0.33 | 16 (13) |
| Pre-eclampsia at enrolment, n (%)<sup>#</sup> | 4 (6) | 5 (8) | 0.47 | 9 (7) |
| Enrolment ultrasound measurements |  |  |  |  |
| Gestational age at enrolment, median weeks+days (IQR) [range] | 23+6<br>(22+3-25+1)<br>[20+1-26+5] | 23+5<br>(22+3-24+4)<br>[20+4-26+4] | 0.48 | 23+5<br>(22+3-24+5)<br>[20+1-26+5] |
| EFW at enrolment, median grams (IQR) (29) | 392 (281-503) | 387 (280-448) | 0.61 | 389 (281-484) |
| EFW z-score at enrolment, median (IQR) (29, 30) | -3.0<br>(-3.6 to -2.5) | -3.2<br>(-3.8 to -2.7) | 0.17 | -3.1<br>(-3.7 to -2.6) |
| Mean UtA PI >95<sup>th</sup> centile at enrolment, n (%)<sup>2</sup> (27) | 49 (79) | 49 (82) | 0.71 | 98 (80) |
| UmA PI >95<sup>th</sup> centile at enrolment, n (%)<sup>3</sup> (27) | 32 (51) | 36 (60) | 0.26 | 78 (55) |
| Absent or reversed UmA end-diastolic flow at enrolment, n (%)<sup>3</sup> | 18 (29) | 25 (42) | 0.11 | 43 (35) |
| MCA PI <5<sup>th</sup> centile at enrolment, n (%)<sup>4</sup> (27) | 10 (16) | 7 (12) | 0.41 | 17 (15) |
| DV a wave absent or reversed at enrolment, n (%)<sup>5#</sup> | 5 (8) | 4 (7) | 0.51 | 9 (8) |
| Pregnancy outcomes |  |  |  |  |
| Pre-eclampsia at any point in pregnancy, n (%)<sup>6</sup> | 24 (39) | 17 (33) | 0.51 | 41 (36) |
| Gestational age at diagnosis of stillbirth or delivery of live birth, median weeks+days (IQR) [range] | 28+2<br>(26+3-34+0)<br>[21+4-39+3] | 28+2<br>(26+4-33+2)<br>[22+2-39+6] | 0.90 | 28+2<br>(26+3-33+2)<br>[22+2-39+6] |
| Female fetus / infant, n (%)<sup>7</sup> | 36 (57) | 24 (43) | 0.19 | 60 (50) |
| Caesarean delivery, n (%) | 42 (67) | 37 (63) | 0.65 | 79 (65) |
| Live births, n (%) | 47 (75) | 43 (72) | 0.71 | 90 (73) |
|  | n=47 | n=43 |  | n=90 |
| Live births ≤28+0 weeks, n (%) | 15 (32) | 14 (33) | 0.95 | 29 (32) |
| Live births >37 weeks, n (%) | 8 (17) | 10 (23) | 0.46 | 18 (20) |
| Caesarean delivery for live births, n (%) | 42 (89) | 37 (88) | 0.85 | 79 (89) |
| Neonatal deaths, n (%)<sup>#</sup> | 5 (11) | 4 (9) | 0.56 | 9 (10) |
| Birth weight z-score for live births, mean (SD) (30) | -3.5 (1.1) | -3.5 (0.9) | 0.97 | -3.5 (1.0) |
| Study outcomes |  |  |  |  |
|  | *n*=63 | *n*=60 |  | *n*=123 |
| Fetal or neonatal death, n (%) | 21 (33) | 21 (35) | 0.85 | 42 (34) |
| Death or delivery ≤28+0 weeks of gestation, n (%) | 30 (48) | 28 (47) | 0.92 | 58 (47) |
|  | *n*=55 | *n*=49 |  | *n*=104 |
| Slow fetal growth, n (%) | 26 (47) | 15 (31) | 0.083 | 41 (39) |
|  | *n*=26 | *n*=20 |  | *n*=46 |
| Development of UmA PI >95<sup>th</sup> centile, n (%) | 12 (46) | 9 (45) | 0.94 | 21 (46) |

<sup>1</sup>*n*=1 missing from validation, <sup>2</sup>*n*=1 missing from discovery, <sup>3</sup>*n*=1 missing from validation, <sup>4</sup>*n*=5 missing from discovery and *n*=1 missing from validation, <sup>5</sup>*n*=3 missing from discovery and *n*=1 missing from validation, <sup>6</sup>*n*=1 missing from discovery and *n*=8 missing from validation, <sup>7</sup>*n*=5 missing from validation, <sup>#</sup>Fisher's exact test. DV=ductus venosus, EFW=estimated fetal weight, MCA=middle cerebral artery, PI=pulsatility index, UmA=umbilical artery, UtA=uterine artery.

**Figure 1:**
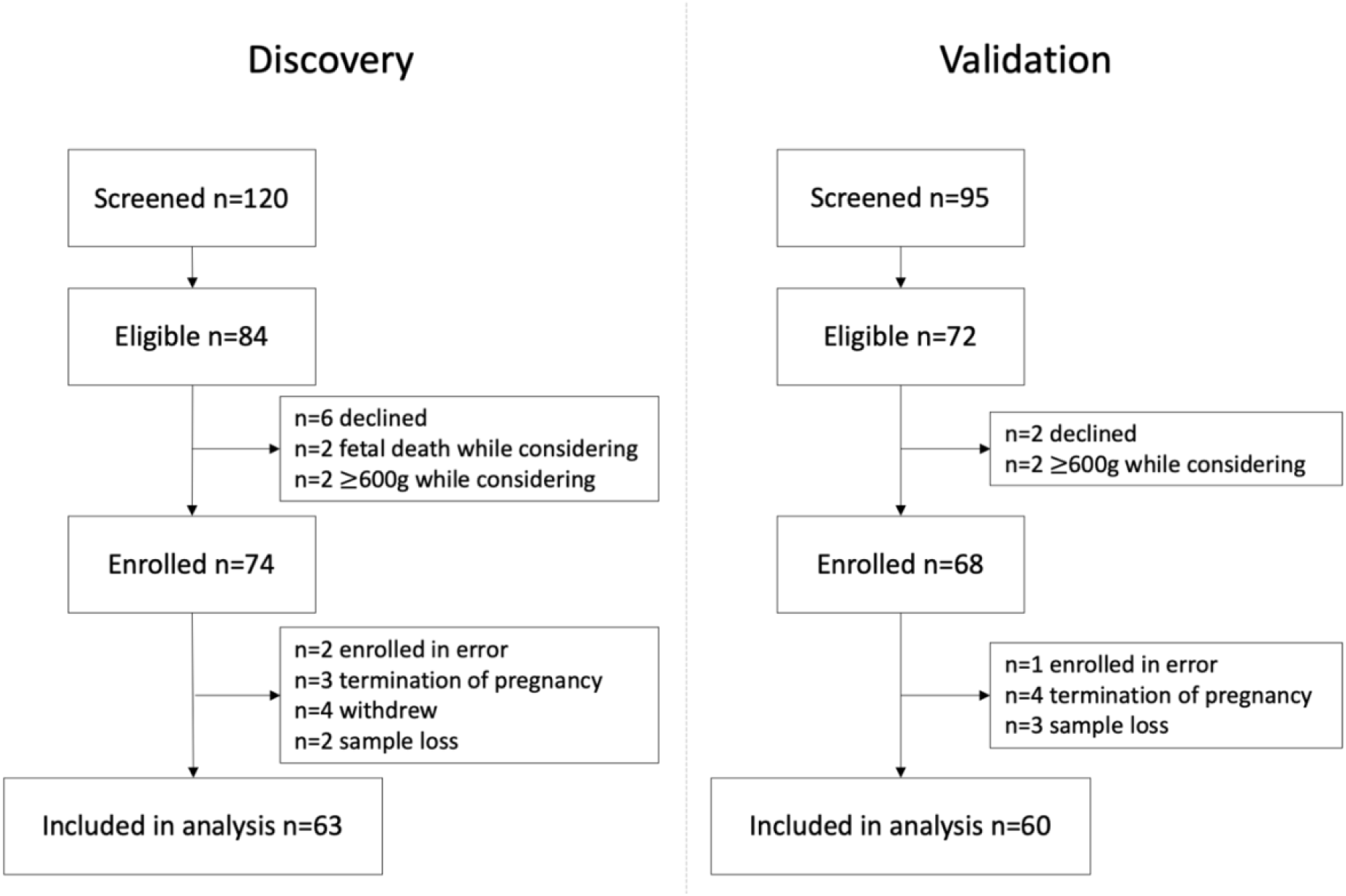
Flow diagram of participant eligibility and enrolment across the four EVERREST Prospective Study centres from 10th March 2014 to 30th January 2020 for the discovery and validation sets.

### Ultrasound measurements as predictors of fetal or neonatal death and death or delivery ≤28+0 weeks of gestation in the discovery set

The best ultrasound predictor of fetal or neonatal death was EFW z-score, either as calculated using the Hadlock 3 formula and Marsal chart (EFW_HM_: AUC 0.81, 95% CI 0.69-0.93) or the Intergrowth formula and chart (EFW_Int_: AUC 0.83, 95% CI 0.71-0.95). UmA category (<95th centile; >95th centile with positive EDF; absent EDF; reversed EDF. AUC 0.75, 95% CI 0.62-0.88) and slow fetal growth (AUC 0.70, 95% CI 0.56-0.83) were also fair predictors. UmA category was the best predictor of death or delivery ≤28+0 weeks (AUC 0.80, 95% CI 0.70-0.91), with mean UtA PI (AUC 0.77, 95% CI 0.65-0.89) and Intergrowth EFW z-score (AUC 0.73, 95% CI 0.60-0.85) also fair predictors (supplementary data Tables 3 & 4).

### Proteomics

Mass spectrometry (MS) profiling of pooled samples gave quantitative information for 200 protein groups (sets of proteins that cannot be distinguished based on peptide sequences), from which human placental lactogen (HPL), fibronectin, pregnancy-specific beta-1 glycoprotein (PSG1), serum amyloid A (SAA) and leucyl-cystinyl aminopeptidase (LNPEP) were selected for individual validation, based on the scoring system outlined in the methods.

### Univariate associations between maternal serum protein concentrations and outcomes in the discovery set

Four proteins were undetectable in most of the samples: VEGF-A, natriuretic peptides B (BNP), melusin, poly[ADP-ribose] polymerase 1 (PARP1). These were excluded from further prediction analyses. The associations between the remaining 98 proteins and the four pregnancy outcomes are summarised in Figure 2. Placental growth factor (PlGF) and HPL concentration were significantly associated with fetal or neonatal death (after Benjamini-Hochberg correction), with fold-changes of 0.52 in pregnancies ending in fetal or neonatal death compared with pregnancies ending in neonatal survival. The concentrations of nine proteins were significantly associated with death or delivery ≤28+0 weeks (after correction). The greatest magnitudes of fold-changes were seen for PlGF (0.28), HPL (0.45), and PSG1 (0.48).

**Figure 2:**
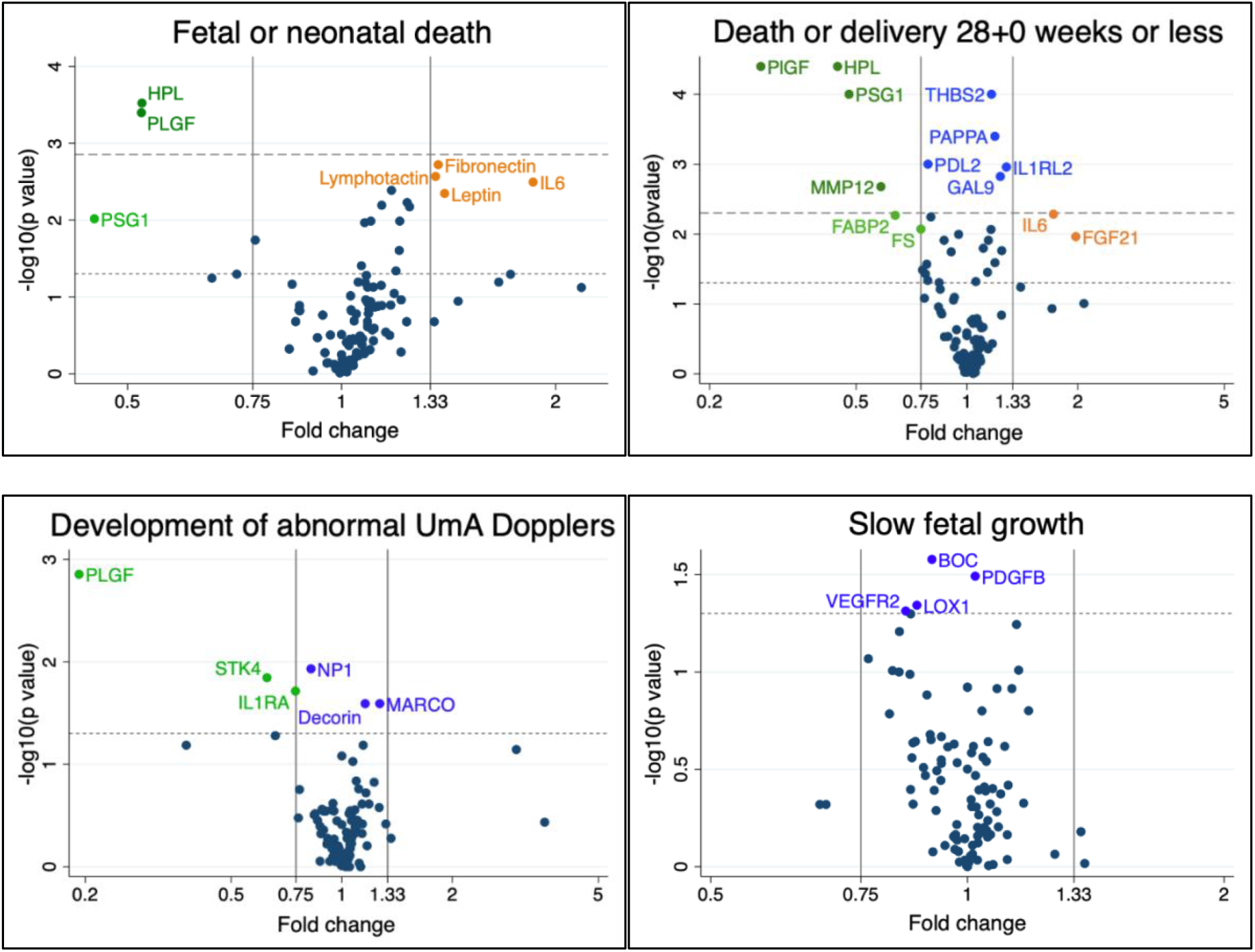
Volcano plots showing the statistical significance and magnitude of the associations between the 98 proteins and four pregnancy outcomes in the discovery set. Associations tested with 2-sided t tests for symmetrical data and Mann-Whitney U tests for skewed data. Dotted line indicates a p value of 0.05. Dashed line indicates the Benjamini-Hochberg cut-off with a 5% false discovery rate (p=0.0015 for fetal or neonatal death, p=0.005 for death or delivery ≤28+0 weeks). See supplementary data Table 5 for full protein names.

### Parenclitic network analysis of the discovery set

Both the networks for fetal or neonatal death and death or delivery ≤28+0 weeks contained clusters centred around HPL (clusters 6 and 4, Figure 3). These clusters also contained pentraxin-related protein PTX3, spondin-2 (SPON2) and thrombomodulin (TM) and contained or were linked to decorin (DCN). The network for death or delivery ≤28+0weeks also contained a cluster centred around PlGF (cluster 2). For all three of the networks that included fetal sex, similar clusters emerged that contained renin (REN), angiopoietin 1 (ANG1), dickkopf-related protein 1 (DKK1) and platelet-derived growth factor subunit beta (PDGFB), contained or were linked to pregnancy-associated plasma protein A (PAPP-A) and in two of the three networks included lectin-like oxidized LDL receptor 1 (LOX1). Networks and associated dendrograms for the development of abnormal umbilical artery Dopplers and slow fetal growth are provided in supplementary data Figures 4 & 5.

**Figure 3:**
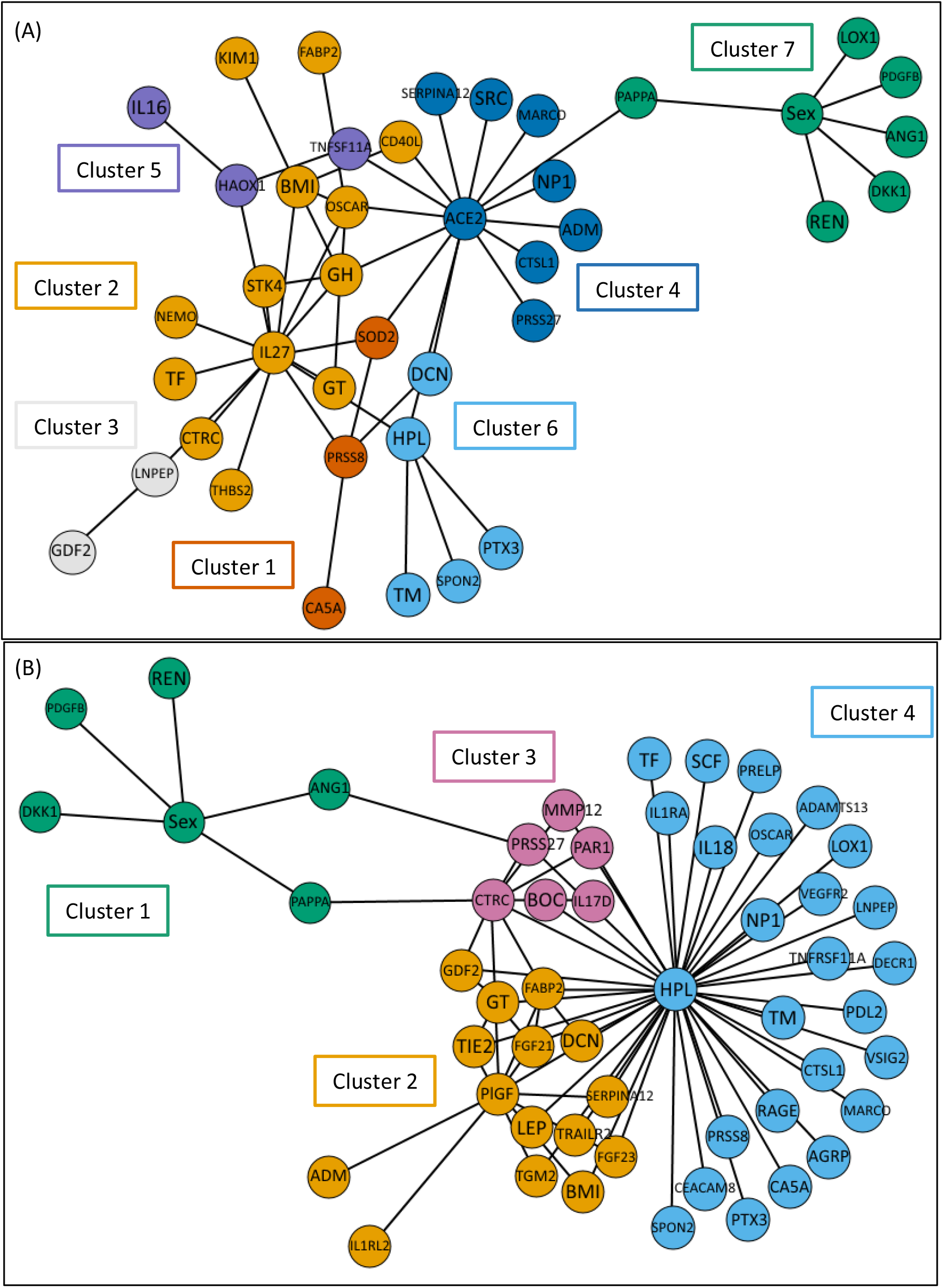
Parenclitic networks for clusters generated based on: (A) fetal or neonatal death (B) death or delivery ≤28+0 weeks. See supplementary data Table 5 for full protein names and supplementary data Figures 2 & 3 for associated dendrograms.

### Model selection by stakeholders

None of the single variable or multivariable models for predicting slow fetal growth performed well enough to warrant validation (AUCs <0.70). For the three remaining outcomes, an online survey was performed to ascertain the priorities of clinicians and patients in predicting outcomes. Forty-five clinicians from 18 countries (of 173 contacted, 26%) and seven patients from the UK who had experienced a pregnancy complicated by severe early-onset FGR (of 36 contacted, 19%) responded (supplementary data Table 6). The prediction of fetal or neonatal death and death or delivery ≤28+0 weeks were considered important or very important by all patients and were also rated highly by clinicians for the purposes of patient counselling and clinical management (Figure 4). Patients and clinicians marginally prioritised sensitivity over specificity for most outcomes (supplementary data Figure 6). For the prediction of the development of abnormal UmA PI, patients universally prioritised sensitivity while clinicians marginally prioritised specificity for patient counselling.

**Figure 4:**
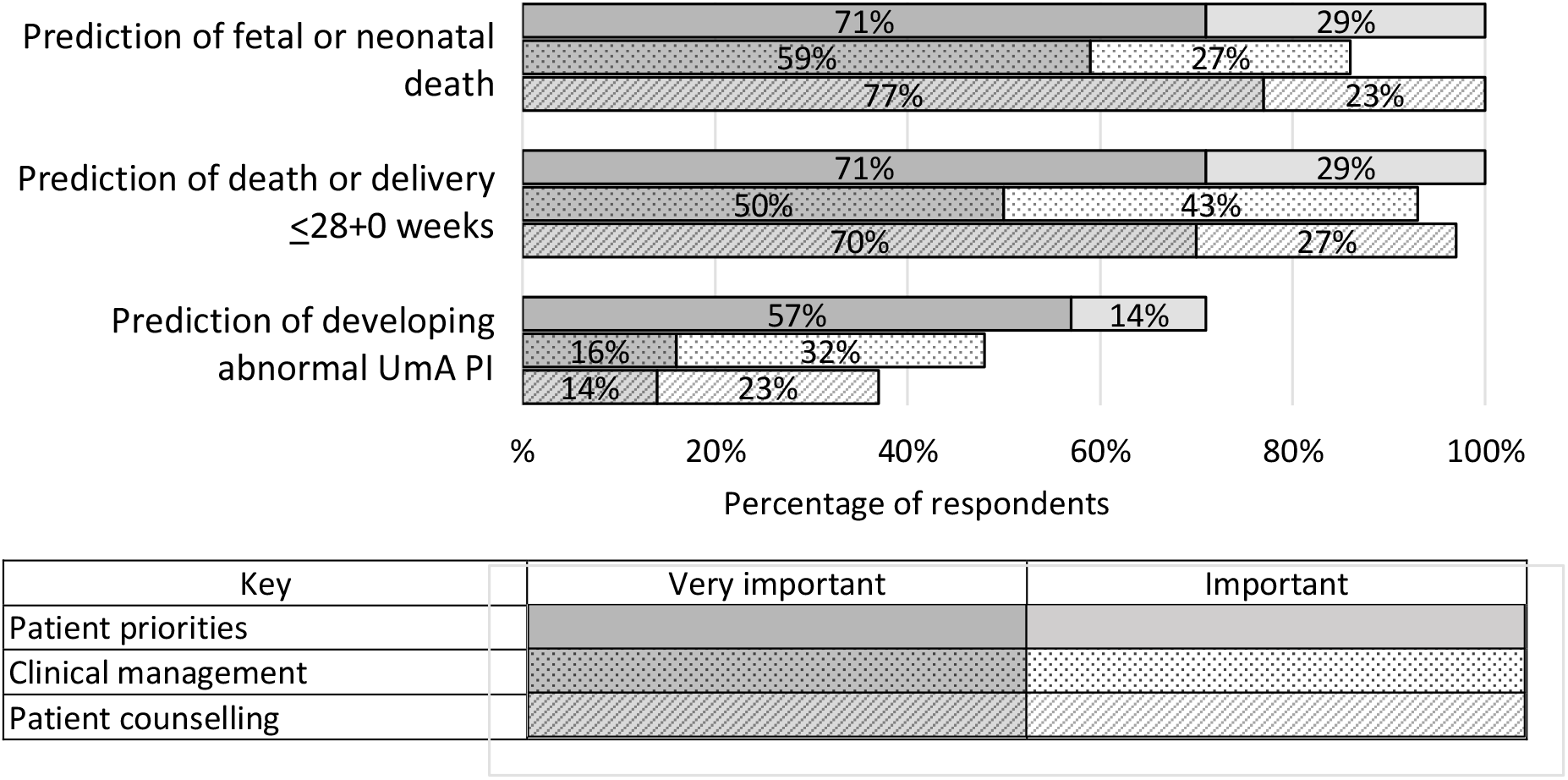
The proportion of patients who ranked our three pregnancy outcomes as ‘important’ or ‘very important’ and the proportion of clinicians who ranked them as ‘important’ or ‘very important’ for clinical management and for patient counselling. UmA PI=umbilical artery pulsatility index.

Based on the survey results, the model performance metrics and the assay reliability, models containing the variables listed in Tables 2 and 3 were selected for validation. For the prediction of death or delivery ≤28+0 weeks, models including HPL marginally outperformed models including PlGF. However, possibly related to the short processing time, the commercial HPL ELISA had high intra-assay variability in our hands (mean coefficient of variation 8.0%, SD 7.3%, 28% requiring repeat analysis for coefficient of variation >10%). Because of this, and the existence of clinically approved tests for PlGF, models including PlGF were selected for validation.

**Table 2:** Model validation for predicting adverse pregnancy outcomes: the seven pre-specified models containing maternal serum proteins. The model not validated is shaded. Validated models include estimates from the combined discovery and validation sets.

| Outcomes | Variable(s) | Discovery<br>(with LOOCV) |  | Validation |  | Combined |  |
| --- | --- | --- | --- | --- | --- | --- | --- |
|  |  | AUC | 95% CI | AUC | 95% CI | AUC | 95% CI |
| Fetal or neonatal death | PIGF | 0.75 | 0.62-0.88 | 0.83 | 0.72-0.95 | 0.81 | 0.73-0.89 |
|  | PIGF & lymphotactin | 0.84 | 0.73-0.95 | 0.75 | 0.62-0.88 | 0.83 | 0.75-0.91 |
|  | PIGF, lymphotactin & fibronectin | 0.85 | 0.74-0.96 | 0.69 | 0.55-0.83 |  |  |
| Death or delivery $\leq 28+0$ weeks | PIGF | 0.86 | 0.76-0.96 | 0.76 | 0.64-0.88 | 0.82 | 0.75-0.89 |
|  | PIGF & pre-eclampsia |  |  |  |  | 0.84 | 0.77-0.91 |
|  | PIGF & PSG1* | 0.91 | 0.82-0.99 | 0.80 | 0.69-0.91 | 0.86 | 0.80-0.93 |
|  | PIGF, PSG1 & pre-eclampsia |  |  |  |  | 0.88 | 0.82-0.94 |
| Development of abnormal UmA PI | PIGF | 0.84 | 0.50-0.95 | 0.64 | 0.38-0.89 | 0.78 | 0.64-0.91 |
|  | PIGF & fibronectin | 0.88 | 0.74-1.00 | 0.65 | 0.39-0.90 | 0.80 | 0.67-0.93 |

\*Discovery AUC 0.906, validation 95% CI 0.688-0.912. LOOCV=leave-one-out cross-validation, PI=pulsatility index, PIGF=placental growth factor, PSG1=pregnancy-specific beta-1 glycoprotein, UmA=umbilical artery. Models were generated and tested using the natural log of PIGF in pg/ml and centered and scaled values for lymphotactin normalised protein expression (NPX) on a log2 scale.

**Table 3:**
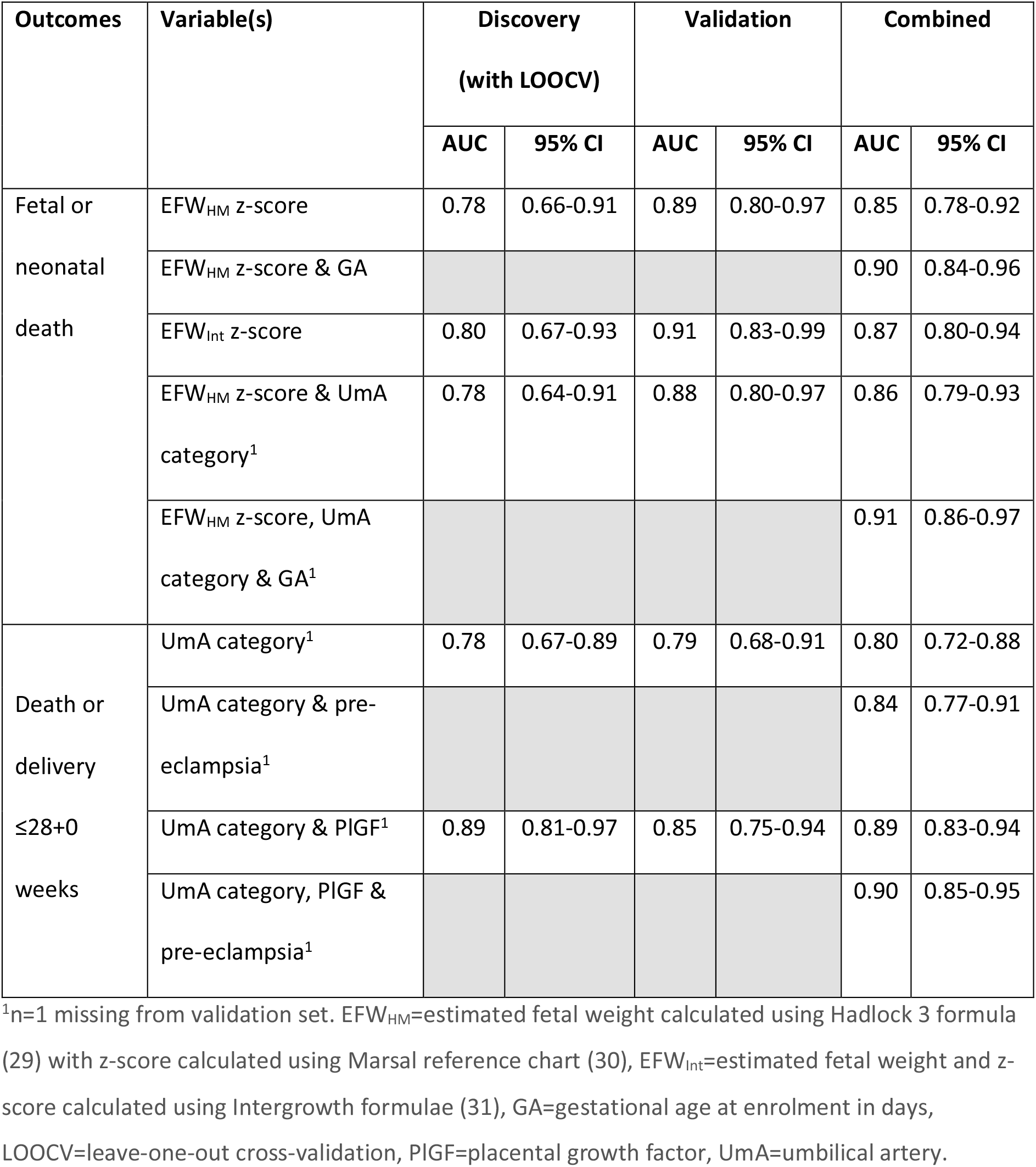
Model validation for predicting adverse pregnancy outcomes: models containing ultrasound measurements, maternal serum protein concentrations and pregnancy characteristics, and their final AUCs from the combined discovery and validation sets.

| Outcomes | Variable(s) | Discovery<br>(with LOOCV) |  | Validation |  | Combined |  |
| --- | --- | --- | --- | --- | --- | --- | --- |
|  |  | AUC | 95% CI | AUC | 95% CI | AUC | 95% CI |
| Fetal or neonatal death | EFW<sub>HM</sub> z-score | 0.78 | 0.66-0.91 | 0.89 | 0.80-0.97 | 0.85 | 0.78-0.92 |
|  | EFW<sub>HM</sub> z-score & GA |  |  |  |  | 0.90 | 0.84-0.96 |
|  | EFW<sub>Int</sub> z-score | 0.80 | 0.67-0.93 | 0.91 | 0.83-0.99 | 0.87 | 0.80-0.94 |
|  | EFW<sub>HM</sub> z-score & UmA category<sup>1</sup> | 0.78 | 0.64-0.91 | 0.88 | 0.80-0.97 | 0.86 | 0.79-0.93 |
|  | EFW<sub>HM</sub> z-score, UmA category & GA<sup>1</sup> |  |  |  |  | 0.91 | 0.86-0.97 |
| Death or delivery ≤28+0 weeks | UmA category<sup>1</sup> | 0.78 | 0.67-0.89 | 0.79 | 0.68-0.91 | 0.80 | 0.72-0.88 |
|  | UmA category & pre-eclampsia<sup>1</sup> |  |  |  |  | 0.84 | 0.77-0.91 |
|  | UmA category & PlGF<sup>1</sup> | 0.89 | 0.81-0.97 | 0.85 | 0.75-0.94 | 0.89 | 0.83-0.94 |
|  | UmA category, PlGF & pre-eclampsia<sup>1</sup> |  |  |  |  | 0.90 | 0.85-0.95 |

<sup>1</sup>n=1 missing from validation set. EFW<sub>HM</sub>=estimated fetal weight calculated using Hadlock 3 formula (29) with z-score calculated using Marsal reference chart (30), EFW<sub>Int</sub>=estimated fetal weight and z-score calculated using Intergrowth formulae (31), GA=gestational age at enrolment in days, LOOCV=leave-one-out cross-validation, PlGF=placental growth factor, UmA=umbilical artery.

### Model validation

Five of the seven protein models (Table 2) and all five of the models containing ultrasound measurements (Table 3), generated in the discovery set, were successfully validated, with AUCs included in the AUC 95% CIs generated from the discovery cross-validation estimates.

### Addition of pregnancy characteristics

Validated models were not significantly improved by the addition of maternal BMI, maternal age, maternal ethnicity or fetal sex. Adding gestational age at enrolment significantly improved the models containing EFW_HM_ alone (LR test p=0.0001) and EFW_HM_ with UmA category (LR test p<0.00005) to predict fetal or neonatal death (supplementary data Table 7). The addition of ‘pre-eclampsia at enrolment’ significantly improved all validated models predicting fetal death or delivery ≤28 weeks of gestation (supplementary data Table 7).

### PlGF values for maximum likelihood ratios

Receiver operating characteristic (ROC) curves are shown in Figures 5 and 6. Model constants and coefficients, along with optimal cut points for positive and negative likelihood ratios and correct classification are provided in supplementary data Tables 8 and 9. Serum PlGF concentration <14.2 pg/ml predicted fetal or neonatal death with a positive likelihood ratio of 18.3, sensitivity of 45% and specificity of 98% and correctly classified 80% of participants. Serum PlGF concentration <14.5 pg/ml predicted death or delivery ≤28+0 weeks with a positive likelihood ratio of 24.7, sensitivity of 38% and specificity of 98% and correctly classified 70% of participants.

**Figure 5:**
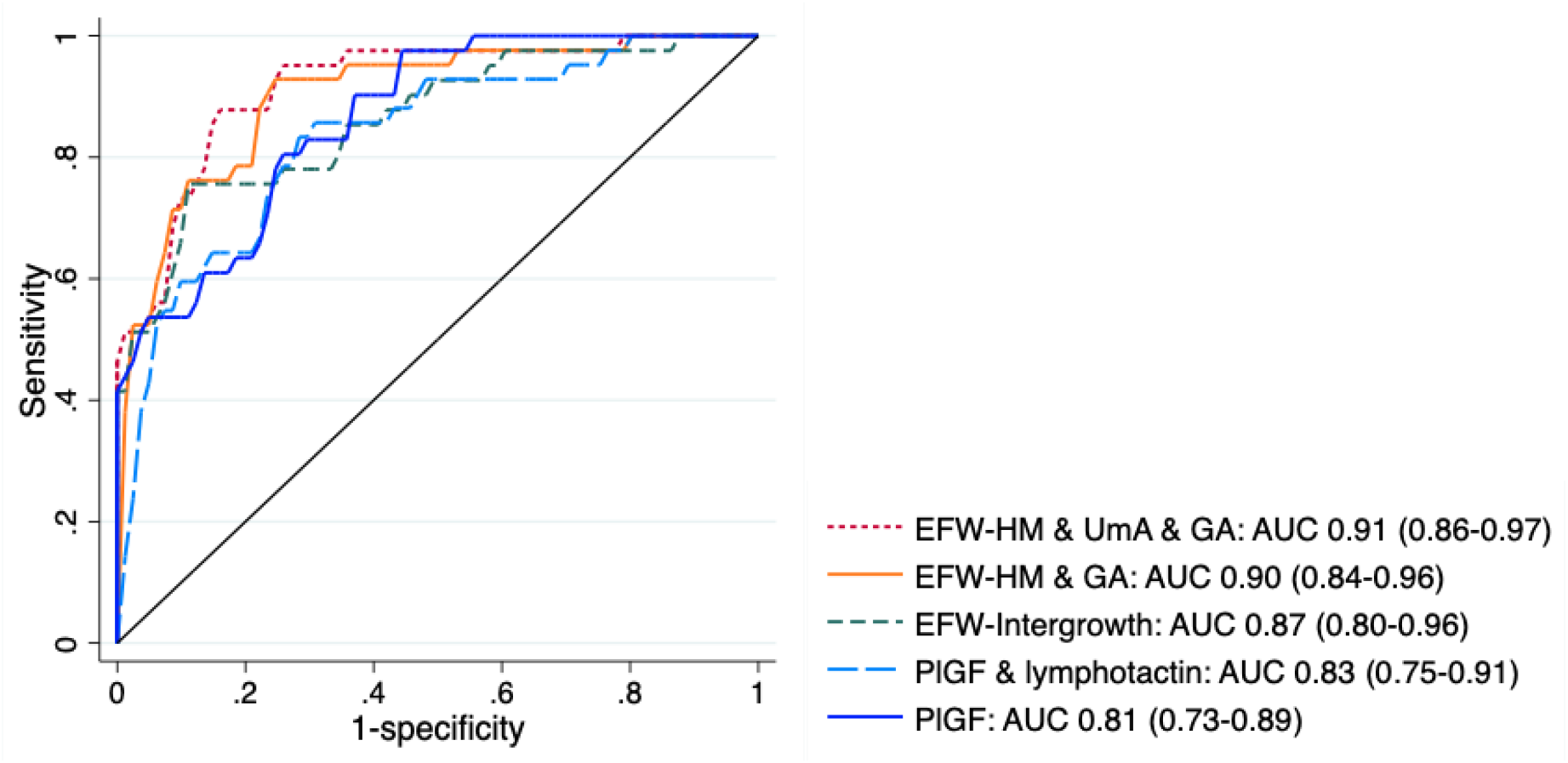
Comparison of the receiver operating characteristic (ROC) curves for the models predicting fetal or neonatal death. EFW-HM=estimated fetal weight calculated using Hadlock 3 formula (29) with z-score calculated using Marsal reference chart (30), EFW-Intergrowth=estimated fetal weight and z-score calculated using Intergrowth formula and reference chart (31), GA=gestational age at enrolment, PlGF=placental growth factor concentration, UmA=umbilical artery Doppler category (0=pulsatility index <95th centile, 1=pulsatility index >95th centile, 2=absent end-diastolic flow, 3=reversed end-diastolic flow).

**Figure 6:**
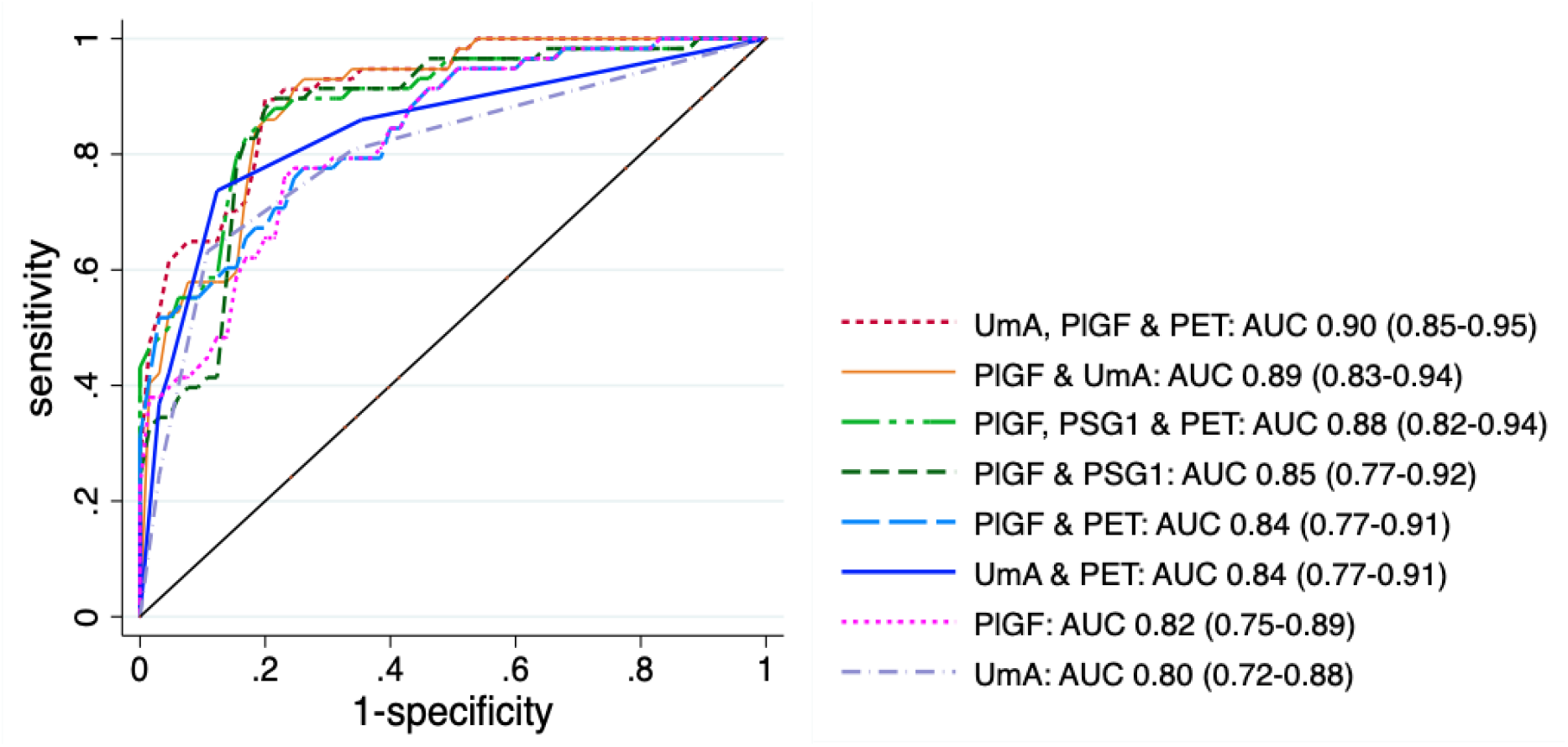
Comparison of the receiver operating characteristic (ROC) curves for the models predicting fetal death or delivery ≤28+0 weeks of gestation. PET=pre-eclampsia at enrolment, PlGF=placental growth factor concentration, PSG1=pregnancy-specific glycoprotein 1 normalised protein expression, UmA=umbilical artery Doppler category (0=pulsatility index <95th centile, 1=pulsatility index >95th centile, 2=absent end-diastolic flow, 3=reversed end-diastolic flow).

### Alternative EFW formulae

Although the EFW z-score calculated using the Intergrowth formula and chart gave the highest AUC for predicting fetal or neonatal death, the Intergrowth formula for estimating fetal weight performed poorly in our sample, especially at lower fetal weights. For the 21 livebirths with a birthweight <600g and an EFW performed within seven days of delivery, the Intergrowth formula overestimated birthweight by a mean of 47% (SD 14%), in contrast to the Hadlock 3 formula which overestimated birthweight by a mean of 25% (SD 10%, supplementary data Figure 8). For all 67 livebirths with an EFW performed within seven days of delivery, the Intergrowth formula overestimated birthweight by a mean of 29% (SD 20%) and the Hadlock 3 formula overestimated birthweight by a mean of 15% (SD 13%). As might be expected, using the EFW calculated from one formula in the model derived from the other had a substantial negative impact on calibration (supplementary data Figure 9).

### Re-analysis of the combined sets

Combining the centred and scaled data from the discovery and validation sets, the strongest associations with both fetal or neonatal death, and death or delivery ≤28+0 weeks were the previously observed negative associations with PlGF (p=1.4×10^−8^ and p=3.0×10^−11^) and HPL (p=1.3×10^−7^ and p=1.4×10^−10^) (Figure 7). The evidence for the negative association between PSG1 and both outcomes was strengthened, as was the evidence for negative associations between matrix metalloproteinase-12 (MMP12) and programmed cell death 1 ligand 2 (PDL2) and death or delivery ≤28+0 weeks. None of the proteins showed an association with the development of abnormal UmA Dopplers or slow fetal growth at a Benjamini-Hochberg 5% false discovery rate (supplementary data Figure 10).

**Figure 7:**
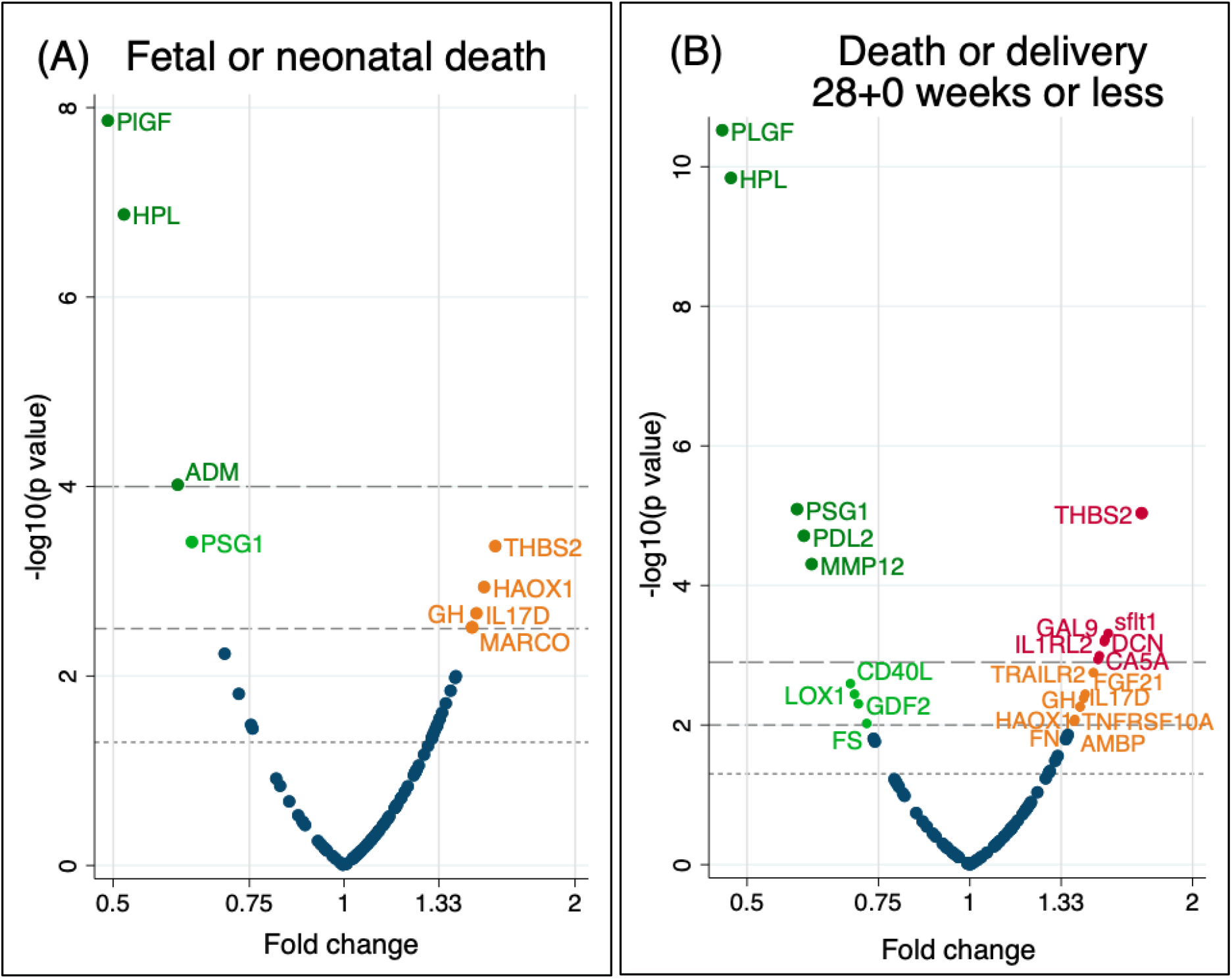
Volcano plots showing the statistical significance and magnitude of associations between fetal and neonatal death and death or delivery ≤28+0 weeks and the centred and scaled concentrations of the 93 proteins from the discovery and validation sets combined. Associations tested with 2-sided t tests. Dotted line indicates p=0.05, short-dashed line indicates Benjamini-Hochberg cut-off with a 5% false discovery rate (A p=0.0048, B p=0.012), long-dashed line indicates Benjamini-Hochberg cut-off with a 1% false discovery rate (A p=0.00032, B p=0.0013). See supplementary data Table 10 for full protein names and individual -log10 p values.

Functional analysis of the proteins associated with fetal or neonatal death at a 5% false discovery rate demonstrated co-expression of HPL and GH. Expanding the network to include intervening proteins resulted in clusters sharing GO biological processes of growth hormone receptor signalling, VEGF signalling and calcitonin family receptor signalling, with proteins in the latter two clusters also involved in angiogenesis and regulation of angiogenesis (Figure 8). Proteins associated with death or delivery ≤28+0 weeks showed multiple interactions, predominantly centred on fibronectin. Shared GO biological processes included those relating to growth (regulation of angiogenesis, cellular response to growth factors and growth hormone receptor signalling pathway via jak-stat), immune function (leucocyte migration, inflammatory response, and positive regulation of T cell activation) or both (regulation of cell adhesion, positive regulation of nik/NF-kappaB signalling) (Figure 9).

**Figure 8:**
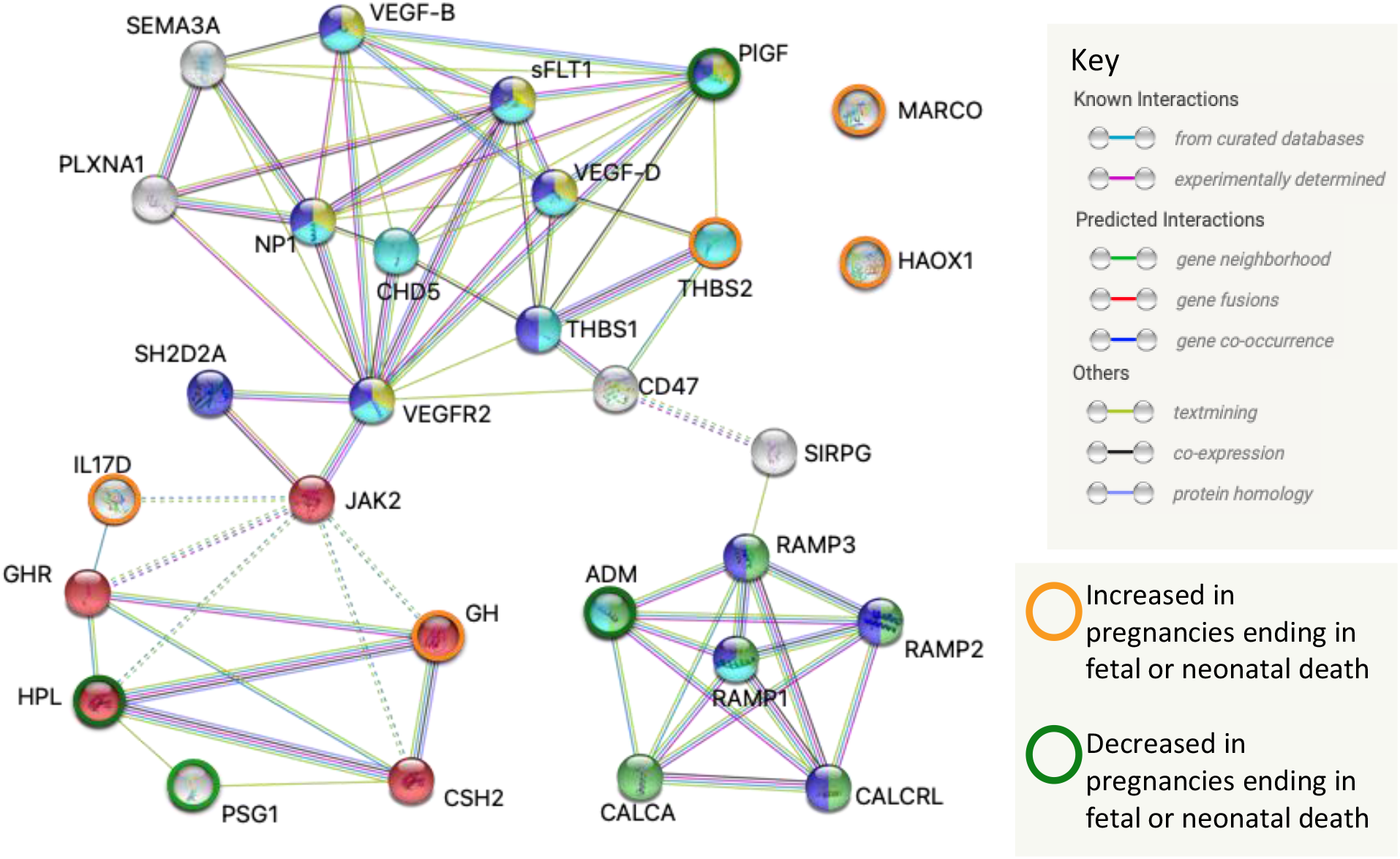
An expanded functional network demonstrating interactions and shared GO biological processes of the proteins associated with fetal or neonatal death in the combined data set at a Benjamini-Hochberg false discovery rate of 5%. Red=growth hormone receptor signalling pathway, yellow=vascular endothelial growth factor signalling pathway, green=calcitonin family receptor signalling pathway, light blue=regulation of angiogenesis, dark blue=angiogenesis. See supplementary data Table 5 for full protein names. Analysis, graphic and legend from STRING (Swiss Institute of Bioinformatics)(32).

**Figure 9:**
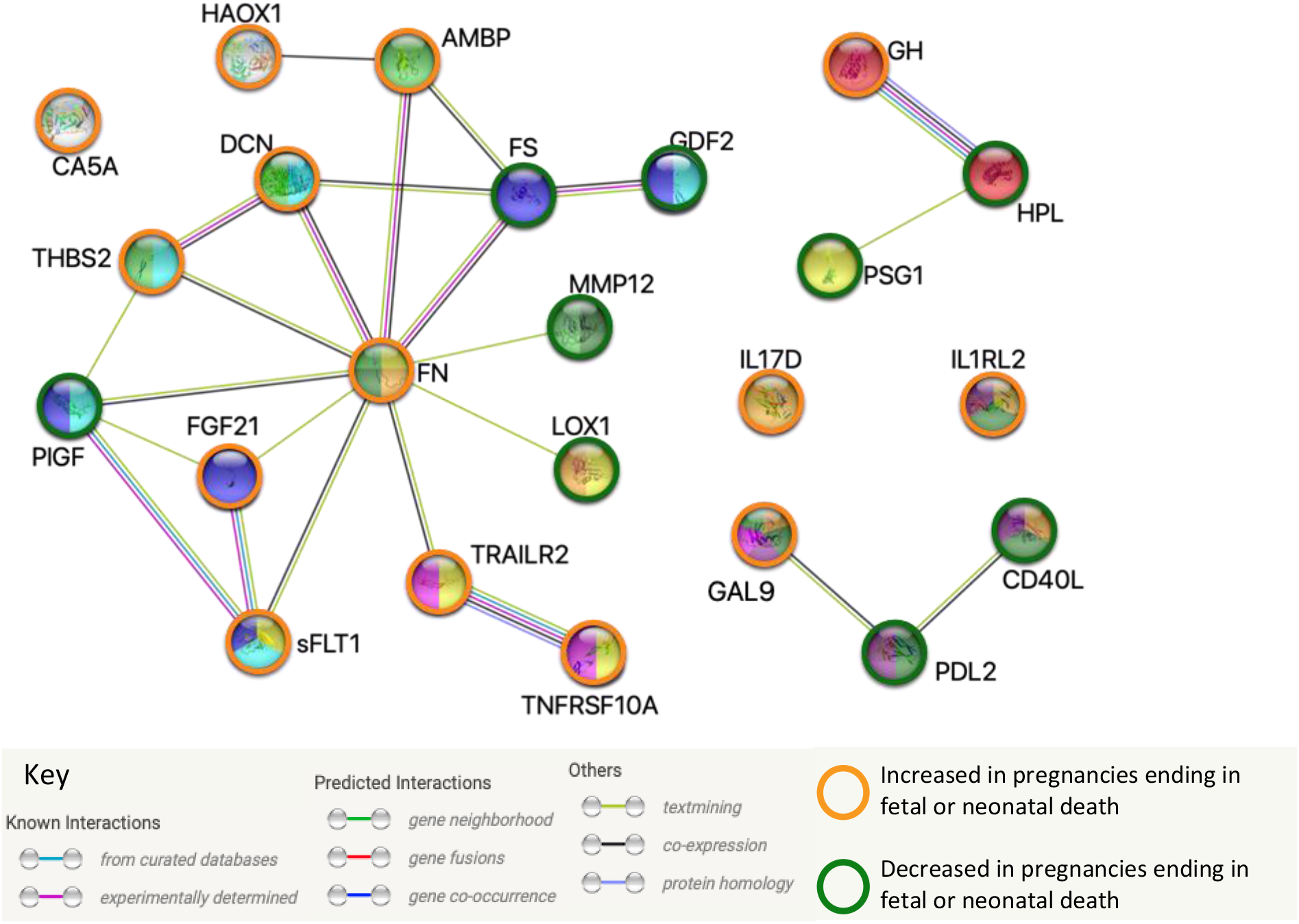
Functional interactions and shared GO biological processes of the proteins associated with death or delivery ≤28+0 weeks in the combined data set at a Benjamini-Hochberg false discovery rate of 5%. Red=growth hormone receptor signalling pathway via jak-stat, orange=inflammatory response, yellow=leukocyte migration, light green=extracellular matrix, dark green=regulation of cell adhesion, light blue=regulation of angiogenesis, dark blue=cellular response to growth factor stimulus, pink=positive regulation of nik/NF-kappaB signalling, purple=positive regulation of T cell activation. See supplementary data Table 5 for full protein names. Analysis, graphic and legend from STRING (Swiss Institute of Bioinformatics)(32).

The three best performing LOOCV models using the combined centred and scaled data all included pro-adrenomedullin (ADM) for predicting fetal or neonatal death, PlGF and HPL for predicting death or delivery ≤28+0 weeks and Platelet-derived growth factor subunit B (PDGFB) for predicting the development of abnormal UmA Doppler measurements (supplementary data Table 12). The emergence of ADM in the models predicting fetal or neonatal death was consistent with the significant association present in the combined but not the discovery sets. In contrast, PDGFB did not show significant univariate associations with the development of abnormal UmA Dopplers in the discovery, validation, or combined data sets.

### Predicting gestational age of livebirth or diagnosis of fetal death and interval from enrolment to livebirth or diagnosis of fetal death

Twelve protein and ultrasound measurements showed an association with the gestational age at which the pregnancies ended in livebirth or fetal death, at a 1% Benjamini-Hochberg false discovery rate (supplementary data Table 13). The best model to predict gestational age at livebirth or fetal death included PlGF and sflt1 concentrations, MMP12 and IL1RL2 NPX and UmA category at enrolment (Figure 10). Eight protein and ultrasound measurements showed an association with the interval between enrolment and either livebirth or the diagnosis of fetal death at a 1% Benjamini-Hochberg false discovery rate (supplementary data Table 13). The best model to predict the interval between enrolment and livebirth or fetal death included PlGF and sflt1 concentrations, MMP12 and decorin NPX, UmA category and gestational age at enrolment (Figure 10). Both models accounted for 68% of the variation in the outcomes they were predicting but had 95% prediction intervals of 40 days, limiting their clinical utility. Sparser models, including PlGF and sflt1 concentrations and UmA category to predict gestational age at livebirth or fetal death, and these same variables plus gestational age at enrolment to predict interval to livebirth or fetal death, had only slightly wider 95% prediction intervals of 42 days (supplementary data Figure 11).

**Figure 10:**
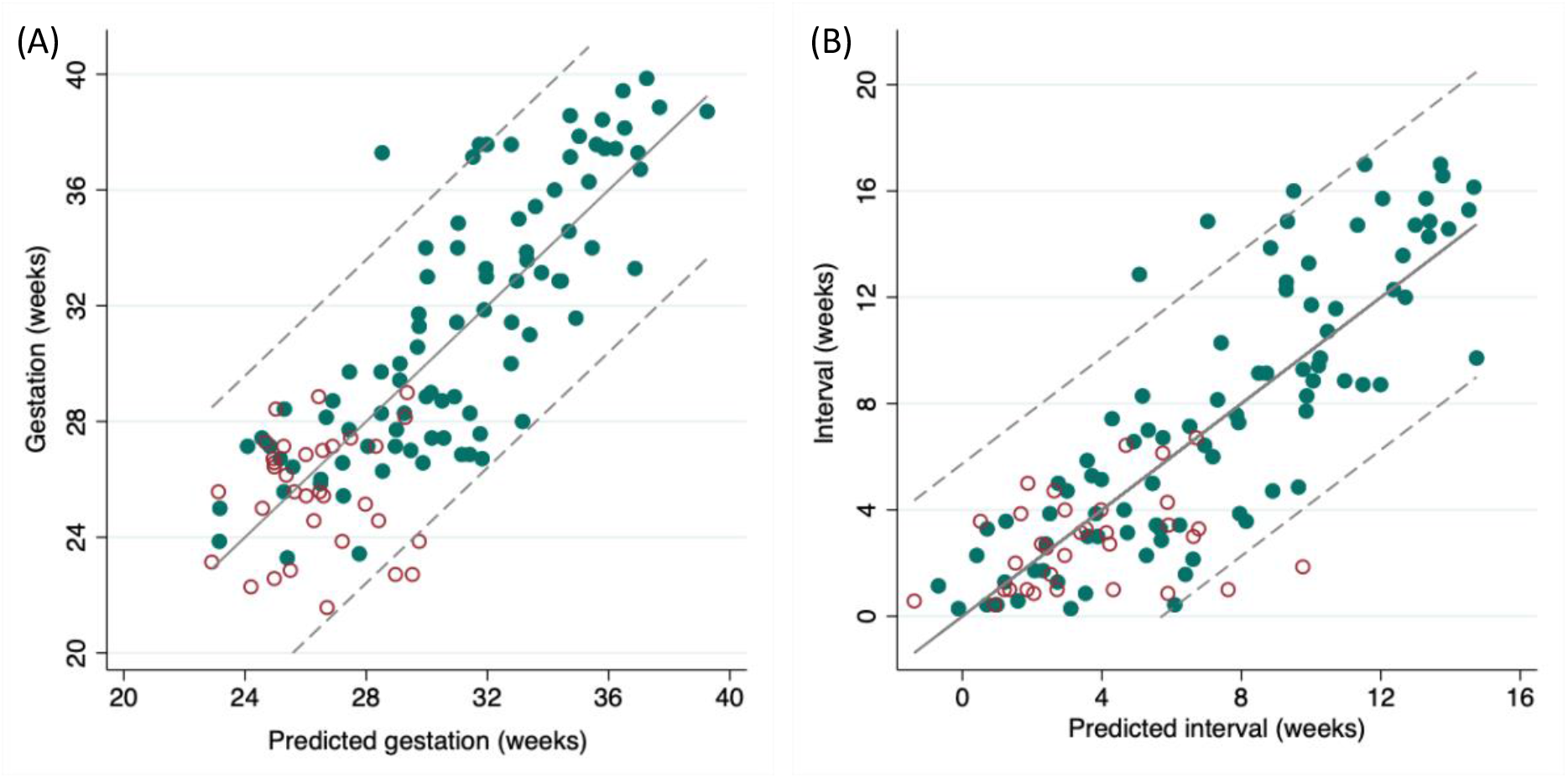
(A) Predicted versus actual gestational age of either livebirth or diagnosis of fetal death, based on the model containing PlGF and sflt1 concentrations, decorin and matrix metalloproteinase 12 normalised protein expression and umbilical artery Doppler category. (B) Predicted versus actual interval from enrolment to either livebirth or diagnosis of fetal death, based on the model containing PlGF and sflt1 concentrations, decorin and matrix metalloproteinase 12 normalised protein expression, umbilical artery Doppler category and gestational age at enrolment. Green filled circles=pregnancies ending in livebirth, red hollow circles=pregnancies ending in fetal death, dotted lines=95% prediction intervals.

### Placental histological classification

Placental samples for histological examination were available for 55 pregnancies (45%); these had similar characteristics and outcomes to the pregnancies without available samples (supplementary data Tables 14 & 15). The only statistically significant difference was a higher proportion of female fetuses among the pregnancies that had placental samples than those that did not (63% vs 41%, p=0.016). Forty-five (82%) placentas showed evidence of placental pathology, with 39 (71%) classified as maternal vascular malperfusion (MVM), three (5%) as villitis of unknown aetiology (VUE), one (2%) as fetal vascular malperfusion (FVM) and two (4%) as non-specific dysmorphic villi. Twelve of the 14 placental samples from pregnancies ending in stillbirth showed MVM, while the three available samples from pregnancies ending in neonatal death showed VUE, FVM and dysmorphic villi. Mean UtA PI, maternal serum PlGF and maternal serum PAPP-A at enrolment all differed significantly between pregnancies with subsequent MVM and pregnancies without MVM. In contrast, none of the umbilical artery parameters studied (UmA category at enrolment; the occurrence of UmA PI >95^th^ centile at any point before delivery; the occurrence of absent or reversed UmA end-diastolic flow at any point before delivery) showed evidence of an association with placental histological classification of MVM (supplementary data Table 16).

## Discussion

### Principle findings and significance

To our knowledge this is the first study to use a discovery science approach, combining ultrasound and biochemical parameters, to identify and validate prognostic markers at the time of diagnosis of severe early-onset FGR. These findings can be used to inform personalised counselling and management of affected pregnancies with outcomes of importance to patients and clinicians. Furthermore, by providing alternative thresholds that prioritise positive and negative likelihood ratios and maximum correct categorisation, eligibility criteria for clinical trials of novel therapeutics can be adapted depending on the perceived risk: benefit ratio of the intervention. Our secondary analyses, including parenclitic network analysis, functional enrichment analysis and triangulation with placental histological classification, provide a deeper characterisation of this unique case series. Some of these findings support and enhance our existing understanding of placental FGR, such as the interplay between angiogenesis, immune cells, and the extracellular matrix (33-35). Other findings offer new avenues for investigation, such as the parenclitic network cluster around fetal sex, which includes proteins related to pericyte function (36).

### Findings in the context of existing literature

Given that ultrasound assessment of biometry and Doppler velocimetry forms the mainstay of identifying and monitoring FGR, it is unsurprising that EFW z-score and umbilical artery category were validated as predictors of fetal or neonatal death and fetal death or delivery ≤28+0 weeks respectively (37-39). A secondary analysis of 105 pregnancies from the UK placebo-controlled trial of sildenafil citrate for early-onset FGR (STRIDER; EFW or AC <10^th^ centile with absent or reversed UmA end-diastolic flow at 22+0-29+6 weeks) identified EFW as an independent predictor of livebirth (OR per 100g 4.3, 95%CI 2.3-8.0, p<0.001) and overall survival (OR per 100g 2.9, 95%CI 1.8-4.4, p<0.001)(40). What is less expected is that absent or reversed ductus venosus a wave was a poor predictor of fetal or neonatal death in our participants (AUC 0.59, 95% CI 0.53-0.66, see supplementary material Table 4), in contrast to the results of previous studies (41, 42). This may reflect a change in clinical practice since these studies were published. Their findings led to ductus venosus waveform becoming an important factor in timing of delivery in extremely pre-term FGR (14, 43), which may have altered the natural history of the disease by prompting delivery before stillbirth could occur.

A limitation of using ultrasound parameters is their potential for variation. In the case of Doppler velocimetry this includes interobserver variability, temporal variation due to factors such as maternal and fetal movement, and variation in umbilical artery waveforms between arteries and along the length of the cord (44-47). There is also considerable variation between different Doppler reference ranges, both in terms of the values of their ‘normal’ ranges and their methodological quality(48). In the case of EFW z-score, variation arises from interobserver variability in measuring biometry, variation in formulae used to generate the EFW and variation in charts used to determine the z-score for gestational age (49-51). Despite the methodological limitations of the Hadlock 3 formula, a recent study of 65 pregnancies with early-onset FGR delivered within seven days of ultrasound assessment found it gave a better combination of systematic and random error than 20 other formulae tested (52).

Several recent studies have highlighted the potential utility of PlGF concentration to predict outcomes in SGA and FGR pregnancies. In a case series of 173 singleton pregnancies with a customised EFW <10^th^ centile between 20+0 and 31+6 weeks, the sflt1/PlGF ratio at diagnosis was an excellent predictor of delivery <30 weeks (AUC 0.96, 95% CI 0.93->0.99) and <34 weeks (AUC 0.94, 95% CI 0.89-0.98) and a good predictor of a composite adverse perinatal outcome (AUC 0.83, 95% CI 0.77-0.90) (53). Similarly, in 116 singleton pregnancies with early-onset FGR (customised EFW <3^rd^ centile or customised EFW <10^th^ centile with abnormal UmA and/or UtA Doppler velocimetry; <32+0 weeks) and positive UmA end-diastolic flow ending in livebirth, women with an sflt1/PlGF ratio ≥85 were significantly more likely to deliver within one, two, three and four weeks from ratio measurement than women with an sflt1/PlGF ratio <85 (54). Composite neonatal morbidity and neonatal admission were also significantly higher following pregnancies with a sflt1/PlGF ratio ≥85 (53.8% vs 28.6% p=0.04; 97.5% vs 67.9% p<0.01). More strikingly, in a series of 130 singleton pregnancies with SGA (AC or EFW <10^th^ centile), fetal demise only occurred in pregnancies with a PlGF <10^th^ centile for gestational age at any time between 16 and 36 weeks (12/65 vs 0/65, p<0.0001) (55).

While these studies revealed the PlGF results to the managing clinicians, similar results have been found in studies where PlGF was not revealed. The secondary analysis of STRIDER UK trial participants, mentioned above, found significant associations between pregnancy outcomes and both the sflt1/PlGF ratio and PlGF alone (40). Higher PlGF concentrations and a lower sflt1/PlGF ratios were associated with greater overall survival (PlGF coefficient 3.67, p<0.001; ratio coefficient 0.51, p=0.002) and later gestation at birth (PlGF coefficient 1.4, p<0.001; ratio coefficient -0.99, p<0.001). Similarly, in a multinational case series of 411 pregnancies, PlGF <5^th^ centile at the time of suspected FGR (AC <10^th^ centile from 20+0 weeks) had an 87.5% sensitivity and 62.8% specificity for predicting stillbirth (56). PlGF <12 pg/ml was associated with a shorter interval to delivery than PlGF >5^th^ centile (13.0 vs 29.5 days, p<0.0001).

Our finding that placental histological classification of maternal vascular malperfusion was significantly associated with lower maternal PlGF concentration and higher mean uterine artery PI at diagnosis of early-onset FGR, but not with UmA Doppler measurements, was in keeping with the results of previous studies (56-59). Agrawal et al. have recently reported that MVM, unlike other placental pathologies, is characterised by raised mean uterine artery PI and a gradual decline in PlGF as the pregnancy progresses (57). Triunfo et al. found in SGA pregnancies (EFW <10^th^) identified between 30 and 34 weeks of gestation, a pattern of placental histopathology they termed ‘placental underperfusion’, was most strongly associated with lower PlGF, measured at the time of diagnosis (58). Benton et al. also found low PlGF to be a better predictor of placental pathology than umbilical artery resistance index or abdominal circumference centile (56).

### Strengths and limitations

The strengths of this multicentre study are that it was carried out prospectively in academic health science centres with fetal medicine experts trained on a common ultrasound protocol, and level 3 perinatal care available for delivery. Participants and their fetuses/neonates were extensively phenotyped at study entry, for the duration of the pregnancy and postnatally, and we report temporally validated results. All pregnancies were managed according to local guidelines, although these were broadly consistent, in line with national and international guidelines (4, 16, 60, 61)and current RCT evidence (e.g. the TRUFFLE trial). This introduces variation, but potentially better reflects real-world practice and hence adds external validity. All serum analysis was carried out after pregnancy outcomes were obtained, using a proteomic discovery science approach which did not assume associations with outcome, but also included additional analysis of proteins anticipated to be related to pregnancy outcome in placental insufficiency. Placental histological classification was blinded to pregnancy outcomes and included control and non-FGR preterm placental samples, to remove some potential bias. Finally, our inclusion of stakeholders to guide model selection means that their predictive value is most important to patients and clinicians.

Our relatively narrow inclusion criteria are both a strength and a limitation, in that they have allowed us to focus on a specific clinical group but have limited our sample size and the generalisability of our findings. The sample size means our study was underpowered to demonstrate small or medium effects and our estimates have wider confidence intervals than larger studies (62). The exclusion of pregnancies <3^rd^ centile but >600g limits the number of pregnancies from 24+6 weeks of gestation to which our findings can be applied and the exclusion of pregnancies with known genetic, chromosomal, and structural differences means our findings cannot be applied to the whole spectrum of FGR. Generalisability is also limited to healthcare settings with comparable neonatal care provision and outcomes, given their impact on neonatal survival and decision making for iatrogenic preterm delivery. Clinicians managing the pregnancies were not blinded to ultrasound measurements, and indeed many management decisions will have been influenced by ultrasound findings. This could have biased the apparent associations between ultrasound variables and pregnancy outcomes, either artificially strengthening or weakening them.

### Future directions

Ideally, our findings should be independently and externally validated. Given the incidence of FGR ≤28+0 weeks this would require another multicentre study. Further research is also needed to determine whether the use of these models would have benefit in practice, both on the psychological wellbeing of parents and on the use of health resources. Future studies to identify and validate predictive models in early-onset SGA (EFW <10^th^ centile <32+0 weeks of gestation) would allow application to a wider population. This would complement the work currently being done in the PLANES study, which is investigating the impact of revealed PlGF in SGA from 32+0 weeks (63). Finally, our primary outcome of fetal or neonatal death provides only short-term information, and data collection for 2-year neurodevelopmental outcomes is on-going.

### Conclusion

In conclusion, our study provides validated models for predicting fetal or neonatal death and fetal death or delivery ≤28+0 weeks of gestation based on ultrasound and maternal serum protein measurements at the time of diagnosis of severe early-onset FGR. The EFW z-score and umbilical artery Doppler velocimetry were the best performing ultrasound parameters, but are vulnerable to inter-rater variability, variation in formulae and reference ranges and temporal variation. The biomarker PlGF was the best performing maternal serum protein for predicting both pregnancy outcomes and maternal vascular malperfusion. This identification of a specific pathological phenotype may be useful for targeting future potential therapies.

## Methods

This study is reported according to the ‘strengthening the reporting of observational studies in epidemiology’ (STROBE) guidelines (64) for cohort studies and the ‘transparent reporting of a multivariable prediction model for individual prognosis or diagnosis’ (TRIPOD) guidelines (65).

### Study design and setting

The EVERREST Prospective Study was a multicentre prospective cohort study recruiting pregnant women from four European tertiary referral centres: University College London Hospital, UK; University Medical Centre Hamburg-Eppendorf, Germany; Maternal-Fetal Unit Hospital Clinic Barcelona, Spain; Skane University Hospital, Lund, Sweden.

### Study population

Full details of the protocol have been published previously (26). In brief, pregnant women were eligible if they had a singleton fetus with an ultrasound estimated fetal weight (EFW) <600g and <3^rd^ centile according to local criteria between 20+0 and 26+6 weeks of gestation. Exclusion criteria were a known abnormal karyotype or major fetal structural abnormality at enrolment (66), indication for immediate delivery, preterm rupture of membranes before enrolment, maternal HIV or hepatitis B or C infection, maternal age under 18 years, any medical or psychiatric condition which compromised a woman’s ability to participate and lack of capacity to consent. Pregnancies with a known congenital infection were not recruited and for the purposes of this analysis pregnancies ending in termination were excluded.

### Outcomes

The primary outcome was fetal or neonatal death (≤28 days of life). Secondary outcomes were: fetal death or delivery ≤28+0 weeks of gestation; slow fetal growth, defined as a worsening of weight deviation of ≥10 percentage points over a two-week interval (including before and after enrolment) or equivalent trajectory over a longer period (28); and the development of abnormal UmA Dopplers, defined as development of UmA PI >95^th^ centile in pregnancies where UmA PI was ≤95^th^ centile at enrolment (27). Ascertainment for outcomes of this study was possible, at the latest, by 29 days of life. Follow-up for neonatal morbidity and infant health and neurodevelopment to the age of 2 years continues.

All pregnancies were managed according to the local fetal medicine unit protocols. Pre-eclampsia was defined according to International Society for the Study of Hypertension in Pregnancy (ISSHP) criteria (67), meaning that, given the presence of FGR, any woman developing new onset hypertension after 20+0 weeks of gestation was classified as having pre-eclampsia rather than pregnancy-induced hypertension. Formalin-fixed placental samples were classified according to Amsterdam consensus criteria by a single assessor (NS) (68). To minimise bias, study placental samples were mixed with placental samples from healthy term pregnancies and pregnancies delivering spontaneously preterm, with NS blinded to pregnancy phenotype and outcome during the assessment.

### Ultrasound measurements

All ultrasound examinations were performed by staff trained and validated to the common EVERREST prospective study protocol (26). At each ultrasound scan Doppler velocimetry of the umbilical artery (UmA), uterine artery (UtA), middle cerebral artery (MCA), ductus venosus (DV) and umbilical vein was performed (69). Local EFW formulae and centile charts were used to determine study eligibility, but for consistency all EFWs were recalculated using the Hadlock 3 formula (incorporating head circumference, abdominal circumference and femur length), with z-scores recalculated using the Marsal chart for descriptive data (supplementary data Equations 1-3) (29, 30). EFWs and z-scores were also recalculated using Intergrowth formulae for analysis (supplementary data Equations 4 & 5) (31). The effect of alternative Doppler reference charts was explored, with similar results to those presented (70-74).

### Blinding

Maternal serum protein concentrations were not available to clinicians, participants or researchers during the pregnancy, as all samples were analysed after complete primary outcome data had been ascertained. Serum PlGF and sflt-1 concentrations were not used as part of clinical care in any of the study centres during the period of recruitment.

### Sample collection

Maternal blood was collected at study enrolment in BD Vacutainer® serum separating tubes and processed according to the manufacturer’s instructions. 500μl serum aliquots were frozen and stored at -80°C. Placental samples, for Amsterdam criteria categorisation, were collected from two areas of each placenta, midway between the cord insertion and margin in areas free from macroscopic infarcts or lesions. Samples were rinsed in PBS, formalin-fixed, wax-embedded, sectioned and stained with H&E.

### Measurement of a priori candidate biomarkers in maternal serum

PlGF and sflt-1 concentrations were measured using Elecsys® electrochemiluminescence immunoassays on a Cobas® e411 analyser (Roche Diagnostics). The normalised protein expression (NPX) of 90 additional proteins associated with cardiovascular disease was measured using the Olink® Cardiovascular II proximity extension assay (full list of proteins in supplementary data Table 17). In the discovery set but not the validation set VEGFA, VEGFD, VEGFR2, neuropilin 1 and endoglin were measured in triplicate using Quantikine® colorimetric sandwich ELISAs (R&D Systems).

### Identification of novel candidate biomarkers in maternal serum using liquid chromatography and tandem mass spectrometry

Five pooled serum samples were created on the following basis: (1) pregnancies ending in fetal or neonatal death (2) pregnancies ending in neonatal survival with delivery <37+0 weeks of gestation (3) pregnancies ending in neonatal survival with delivery 37+0 weeks of gestation or more (4) slow fetal growth trajectory (5) normal fetal growth trajectory. Pooled serum samples were depleted of 12 high-abundance proteins using Proteome Purify™ 12 resin, as per the manufacturer’s instructions (R&D Systems), concentrated using Vivaspin© 500 5kDa Molecular Weight Cut-Off columns (GE Healthcare), reduced with 10mM tris(2-carboxyethyl)phosphine hydrochloride then alkylated with 7.5mM iodoacetamide. Pooled samples were digested using a trypsin/Lys-C mix, labelled with Tandem Mass Tags™ (Thermo Fisher Scientific) and combined(75). The combined sample underwent two-dimensional high-performance reverse-phase liquid chromatography and tandem mass spectrometry. In the first dimension, samples were fractionated into 30 at high pH using a Poroshell 300 Extend C18 column (Agilent), following which fractions 1-4 were combined with fractions 27-30 respectively due to low abundance in the first four fractions. The second fractionation was performed on the Ultimate 3000 nano-liquid chromatography system using Acclaim™ PepMap™ 100 C18 pre-columns and Acclaim™ PepMap™ 100 C18 Nano-LC columns run in tandem with analysis on the LTQ (linear trap quadrupole) Orbitrap XL™ 2.5.5 (all Thermo Fisher Scientific). A blank calibration sample was run after every three fractions, and a standard sample of known mass run after every six fractions, for quality control.

Proteins were identified using Proteome Discover V1.4 software (ThermoFisher Scientific) to search the human Swiss-Prot database with the Mascot search engine (Matrix Science Ltd.). Proteins were scored on variability, peptide count, ubiquity, ratio between pools and consistent trend across pools (supplementary data Tables 18 & 19). Expression pattern clusters, based on standardised and raw quantification ratios, were generated using the Graphical Proteomics Data Explorer (GPRoX) platform. Based on their scores and expression clusters, five candidate proteins were selected and measured in individual samples using ELISAs. Fibronectin, PSG1 (both R&D Systems) and HPL (DRG International) were measured in the discovery and validation sets while SAA (R&D Systems) and LNPEP (Cloud-clone) were measured in the discovery set only.

### Priority survey and model selection

An online survey was sent to patients and clinicians asking their opinion on the importance of different pregnancy outcomes and, for each outcome, whether they would prioritise sensitivity or specificity. Models were selected on the basis of the survey results and the model performance metrics described below. Protein models were published online prior to the validation data analysis.

### Sample size

Since this work involved discovery of novel biomarkers, a formal *a priori* sample size calculation was not possible. Before analysing the discovery set it was determined that this sample of n=63 with 21 fetal or neonatal deaths gave an 80% power to detect a standardised effect size of 0.9 (large) to a significance level of 0.05 (76).

### Statistics

Data analysis was performed using STATA/MP 16.1 software (StataCorp LLC, College Station, USA) unless otherwise specified. Descriptive and investigative variables were tested for skew and kurtosis (77, 78) and handled as symmetrical if there was no evidence of either. PlGF, sflt1, endoglin, VEGFD, NP1, HPL, SAA and LNPEP were transformed to their natural logs and multiplex data was analysed as provided, on a log2 scale. Characteristics of the discovery and validation sets were compared using chi-square tests (categorical data), Fisher’s exact tests (binary data with sparse outcomes), 2-sided t tests (symmetrical continuous data) and Mann-Whitney U tests (skewed continuous data).

Missing data for BMI (n=5) were imputed using chain equations. Umbilical artery (UmA) PI at enrolment was systematically missing (n=16), with most missing cases having absent or reversed end-diastolic flow (EDF, n=15). Umbilical artery Doppler velocimetry was therefore handled as an interval variable, ‘UmA PI category’,, where 0=UmA PI ≤95^th^ centile, 1=UmA >95^th^ centile with positive EDF, 2=absent EDF, 3=reversed EDF. Where uterine artery (UtA) PI at enrolment was missing (n=10) a mean UtA PI below or above the 95^th^ centile could be inferred in nine cases where the mean UtA PI was consistently normal (n=1) or abnormal (n=8) respectively at scans prior to and after enrolment and UtA PI values were imputed using multiple imputation. Associations between ultrasound measurements and both fetal or neonatal death and death or delivery ≤28+0 weeks of gestation, were analysed using logistic regression. Univariate associations between protein concentrations or NPX and outcomes were assessed using 2-sided t tests, Mann-Whitney U tests and logistic regression, with Benjamini-Hochberg procedures to account for multiple comparisons.

### Model development

Two-protein models for the development of abnormal UmA Dopplers and two- and three-protein models for the other three pregnancy outcomes, with internal validation using leave-one-out cross-validation (LOOCV), were compared based on AUC, specificity for 90% sensitivity, sensitivity for 90% specificity, F1 score, Matthews correlation coefficient (MCC) and precision-recall characteristics (PRROC) AUC. ROC curves were generated with the *pROC* R package (version 1.18.0, https://cran.r-project.org/web/packages/pROC/index.html). 95% confidence intervals for AUCs were determined by stratified bootstrapping. PRROC curves were generated with the MLmetrics R package (version 1.1.1, https://cran.r-project.org/web/packages/MLmetrics/index.html). Models with variance inflation factors of five or more were excluded using the following R package: https://cran.r-project.org/web/packages/car/index.html, version 3.1-0. Two-variable models predicting fetal or neonatal death and death or delivery ≤28+0 weeks of gestation containing ultrasound parameters with or without PlGF or HPL (as the proteins showing the strongest associations with these outcomes) were compared in the same way. Outcomes and protein models to be validated were published on the study registry prior to analysis of the validation data.

### Parenclitic network analysis

Parenclitic networks of the 102 proteins were generated for each of the four pregnancy outcomes. For each outcome, two-dimensional kernel density estimations were generated for every pair-combination of variables in ‘controls’ (pregnancies without the outcome). Individual networks were then generated for each ‘case’, with linkages created if a pair-wise relationship of variables differed from the control distribution by more than a given threshold(79). These individual case networks were then combined. VEGF-A, BNP, PARP1 and melusin were included as binary variables of ‘detectable’ or ‘not detectable’. Booking body mass index (BMI) and fetal sex were included as variables in the networks, except for ‘development of abnormal UmA Dopplers’, where the sample was not large enough to accommodate them.

### Model validation

Concentrations of HPL and PSG1, as measured by ELISA, and NPX values for the Olink multiplex proteins showed substantial variation in centrality and spread between the discovery and validation sets. To account for this, values of each protein were centred to a mean of 0 and scaled to an SD of 1 in the discovery set and validation set separately. These centred and scaled values were used for subsequent analyses, including model validation. Concentrations of PlGF, sflt1 and fibronectin did not require transformation.

Models generated from the discovery set were run on data from the validation set and were considered validated if the 95% CI for the validation estimate of AUC included the LOOCV AUC estimate from the discovery set. For validated models, data from both sets were combined to give final test characteristics. Likelihood ratio tests were used to determine whether the addition of pregnancy characteristics (maternal BMI, maternal age, maternal ethnicity, fetal sex, gestational age at enrolment and pre-eclampsia at enrolment) significantly improved the validated models. Model calibration was assessed by plotting predicted probability against observed frequency of outcome.

### Functional interactions

Centred and scaled data from the discovery and validation sets were combined to retest univariate associations with the primary and secondary outcomes. Proteins showing a significant association at a 5% Benjamini-Hochberg false discovery rate were explored for physical and functional interactions and for enrichment of GO biological processes, relative to the background of all proteins measured, using STRING (Swiss Institute of Bioinformatics)(32). Where no enrichment was detected shared GO biological processes were identified through comparison to the whole genome.

### Modelling pregnancy duration

Protein and ultrasound measurements from the combined discovery and validation sets were tested for their association with gestational age at livebirth or diagnosis of fetal death and interval from enrolment to livebirth or diagnosis of fetal death using linear regression. Variables showing a significant association at a 1% Benjamini-Hochberg false discovery rate were used to create linear models predicting these outcomes in a stepwise fashion. Maternal age, BMI, ethnicity, gestational age at enrolment, pre-eclampsia at enrolment and fetal sex were tested for model improvement. Model fit was tested by assessing variance inflation factors for multicollinearity, assessing the distribution of the residuals for heteroscedasticity and outliers, and looking for observations with high leverage. UmA and UtA Doppler velocimetry, PlGF concentration, HPL concentration and PAPP-A NPX were tested for their associations with placental histological classification using logistic regression.

### Study approval

Ethical approval was provided by the National Research Ethics Service Committee London - Stanmore in the UK (REC reference: 13/LO/1254), the Hospital Clinic of Barcelona’s Clinical Research Ethics Committee in Spain (Reg: HCB/2014/0091), the Regional Ethical Review Board in Lund for Sweden, (DNr 2014/147) and the Ethics Committee of Hamburg Board of Physicians in Germany (PV4809). This study was conducted according to the Declaration of Helsinki principles and written informed consent was given by all participants before enrolment.

## Data Availability

The full data set will not be made publicly available because the degree of detailed phenotyping could allow individual patient identification. Limited data sharing may be possible, with the agreement of the consortium, on request to RS or ALD.

## Acknowledgements

The research leading to these results has received funding from the European Union Seventh Framework Programme (FP7/2007-2013) under grant agreement no. 305823, the Rosetrees Trust and the Mitchell Charitable Trust in memory of Shoshana Mitchell Glynn. This research has been supported by the National Institute for Health Research University College London Hospitals Biomedical Research Centre (RS, NM, ALD). NRN would like to thank the support from Cancer Research UK (C12077/A26223).

This work would not have been possible without the contribution of the late Professor John Timms, Elizabeth Garrett Anderson Institute for Women’s Health, University College London. His contribution to the design, conduct and analysis of the study was invaluable, and we would have been honoured to have him as a co-author. We would also like to thank all of the women and families who took part in this study as well as staff at the UCL Comprehensive Clinical Trials Unit (Anna Morka, Jade Dyer, Helen Knowles, Steve Hibbert, Kate Maclagan), Gina Buquis, Jade Okell, Dr Mark Lees, Dr Carlo Rossi, Dr Tara Krishnan, Dr Roberta Morris, Dr Sarah Guillon, Richard Gunu, Dr Eva Sedlak, Professor Alexei Zaikin and Dr Oleg Blyuss.

