## Supplementary data for "Ultrasound and biochemical predictors of pregnancy outcome at diagnosis of early-onset fetal growth restriction"

**Contents of Supplementary Data**

Figure 11: (A) Predicted versus actual gestational age of either livebirth or diagnosis of fetal death, based on the sparser model containing PlGF and sflt1 concentration and umbilical artery Doppler category (B) Predicted versus actual interval from enrolment to either livebirth or diagnosis of fetal death, based on the sparser model containing PlGF and sflt1 concentrations, umbilical artery Doppler category and gestational age at enrolment. ...................................................................... 30

### Results

Table 1: Recruitment by study centre for the discovery and validation sets.

| Centre | Discovery (n %) | Validation (n %) | Combined (n %) |
|---|---|---|---|
| University College London Hospital, UK | 48 (76) | 52 (87) | 100 (81) |
| Maternal-Fetal Unit Hospital Clinic Barcelona, Spain | 6 (10) | 4 (7) | 10 (8) |
| University Medical Center Hamburg-Eppendorf, Germany | 5 (8) | 2 (3) | 7 (6) |
| Skane University Hospital, Lund, Sweden | 4 (6) | 2 (3) | 6 (5) |

Table 2: Core maternal and neonatal outcomes recommended for reporting by the COSGROVE (Core Outcome Set for the prevention and treatment of fetal GROwth restriction: deVeloping Endpoints) consensus process. Childhood outcomes at 2 years of age are still being assessed and will be reported in due course.

| Domain | Outcome | |
|---|---|---|
| Maternal (n=123) | Eclampsia (n) | 0 |
| | Maternal death (n) | 0 |
| Neonatal (n=90) | Birthweight (g, median IQR) | 953 (590-1650) |
| | Birthweight <3rd centile (n %) | 83 (92%) |
| | Birthweight <10th centile (n %) | 88 (98%) |
| | Need for mechanical ventilation (n %) | 66 (73%) |
| | Bronchopulmonary dysplasia / chronic lung disease (n%)* | 47 (59%) |
| | Necrotising enterocolitis (n %) | 10 (11%) |

*n=9 neonatal deaths before 36 weeks corrected age, n=2 missing.

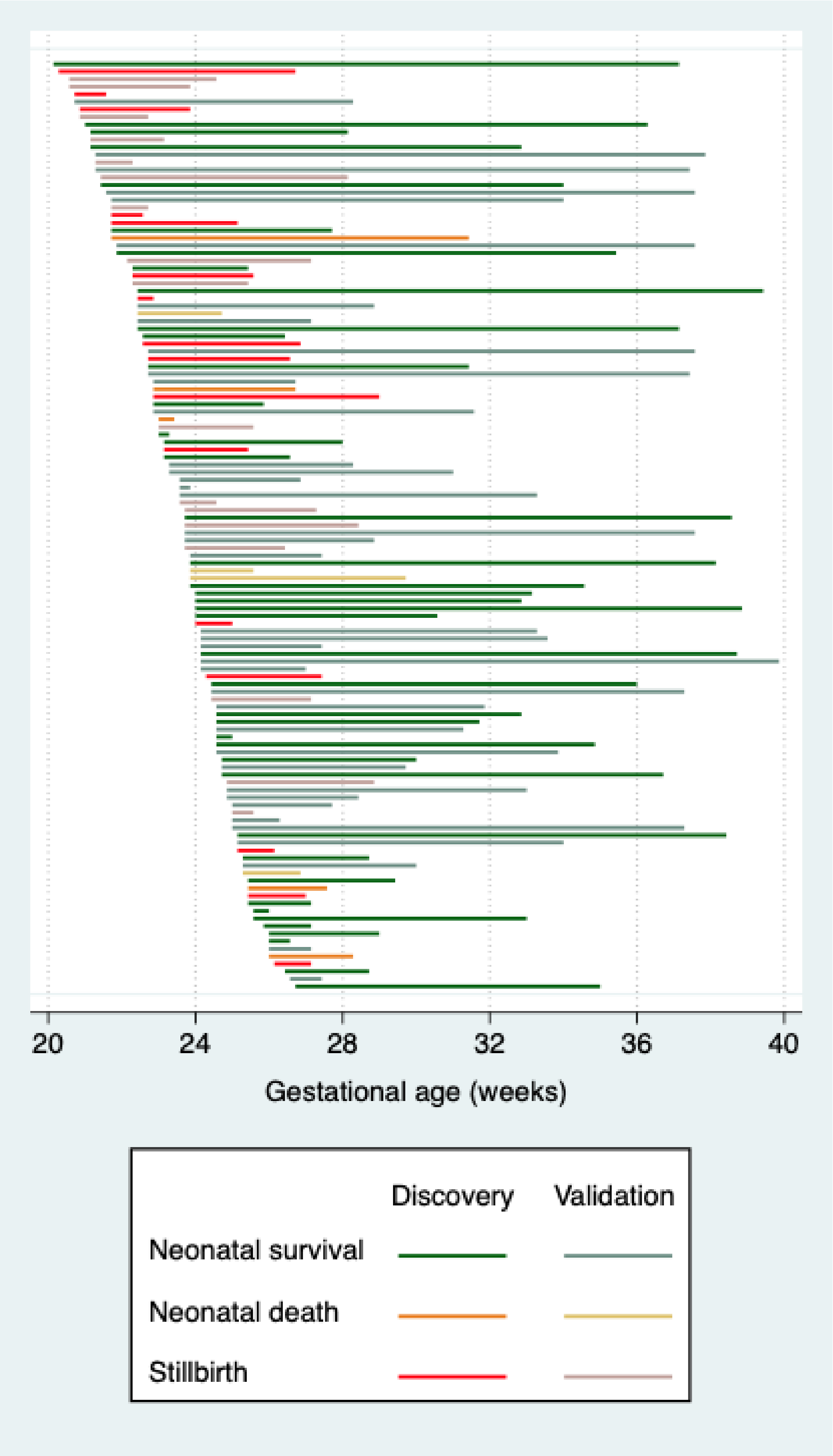

Figure 1: Gestational age range from enrolment to either live birth or diagnosis of intrauterine fetal death for all participants, sorted by gestational age at enrolment.

Table 3: Associations between ultrasound measurements at enrolment and fetal or neonatal death and death or delivery ≤28+0 weeks of gestation in the discovery set.

| Ultrasound variables | | Fetal or neonatal death | | | | Death or delivery ≤28+0 weeks of gestation | | | |
|---|---|---|---|---|---|---|---|---|---|
| | | OR (95% CI) | p value | RR (95% CI) | AUC (95% CI) | OR (95% CI) | p value | RR (95% CI) | AUC (95% CI) |
| UmA PI >95th centile (Schaffer and Staudach 1997) | n=63 | 5.2 (1.6-17.0) | 0.006 | 3.1 (1.4-8.5) | 0.69 (0.57-0.81) | 8.8 (2.8-27.4) | 0.0002 | 3.2 (1.4-7.4) | 0.75 (0.64-0.86) |
| Absent or reversed UmA EDF | n=63 | 5.5 (1.7-17.9) | 0.005 | 2.8 (1.7-6.5) | 0.68 (0.55-0.80) | 17.7 (3.6-87.7) | 0.0004 | 2.9 (1.4-5.9) | 0.74 (0.64-0.84) |
| UmA category[1] | n=63 | 2.8 (1.6-5.2) | 0.001 | 1.8 (1.2-2.6) | 0.75 (0.62-0.88) | 4.4 (2.1-9.3) | 0.0001 | 1.7 (1.2-2.3) | 0.80 (0.70-0.91) |
| MCA PI <5th centile (Schaffer and Staudach 1997) | n=58 | 1.3 (0.3-5.4) | 0.69 | 1.2 (0.4-3.6) | | 2.8 (0.64-12.0) | 0.18 | 1.5 (0.7-3.6) | |
| Absent or reversed DV a wave | n=60 | 10.7 (1.1-103.3) | 0.04 | 2.9 (1.0-8.8) | 0.59 (0.50-0.69) | 4.8 (0.50-45.8) | 0.18 | 1.8 (0.6-5.1) | |
| Mean UtA PI | n=62 | 1.7 (0.7-4.1) | 0.23 | 1.4 (0.7-2.8) | | 6.4 (2.1-19.3) | 0.001 | 2.1 (1.2-3.8) | 0.77 (0.65-0.89) |
| Mean UtA PI >95th centile (Schaffer and Staudach 1997) | n=62 | 1.8 (0.4-7.3) | 0.43 | 1.5 (0.4-5.1) | | 1.4 (1.1-1.7) | 0.002 | 7.4 (1.0-54.6) | 0.76 (0.64-0.89) |
| $EFW_{HM}$ z-score (Hadlock et al. 1985, Marsal et al. 1996) | n=63 | 0.17 (0.06-0.45) | 0.0003 | 0.56 (0.40-0.77) | 0.81 (0.69-0.93) | 0.35 (0.17-0.72) | 0.005 | 0.71 (0.52-0.95) | 0.69 (0.56-0.82) |
| $EFW_I$ z-score (Stirnemann et al. 2017) | n=63 | 0.37 (0.20-0.67) | 0.001 | 0.80 (0.71-0.91) | 0.83 (0.71-0.95) | 0.54 (0.36-0.83) | 0.004 | 0.87 (0.77-0.97) | 0.73 (0.60-0.85) |
| Slow fetal growth[2] | n=55 | 5.4 (1.4-19.8) | 0.012 | 3.3 (1.1-10.4) | 0.70 (0.56-0.83) | 2.6 (0.86-7.8) | 0.09 | 1.7 (0.8-4.0) | |

[1]Four levels: ≤95th centile, >95th centile with positive EDF, absent EDF, reversed EDF. [2]Slow fetal growth, defined as a worsening of percentage weight deviation by ≥10 percentage points over 2 weeks or equivalent (Marsal 2009), was assessed over a minimum of 2 weeks so included data from follow-up scans. DV=ductus venosus, EDF=end-diastolic flow, $EFW_{HM}$=estimated fetal weight calculated using Hadlock 3 formula with z-score calculated using Marsal reference chart, $EFW_I$=estimated fetal weight and z-score calculated using Intergrowth formulae, MCA=middle cerebral artery, OR=odds ratio, PI=pulsatility index, RR=risk ratio (estimated using Poisson regression), UmA=umbilical artery, UtA=uterine artery.

Table 4: Associations between ultrasound measurements at enrolment and (1) fetal or neonatal death and (2) death or delivery ≤28+0 weeks of gestation in the combined discovery and validation sets. This is provided for reference but was not available at the time of model selection as models were selected prior to validation set recruitment.

| Ultrasound variable | | Fetal or neonatal death | | | | Death or delivery ≤28+0 weeks of gestation | | | |
|---|---|---|---|---|---|---|---|---|---|
| | | OR (95% CI) | p value | RR (95% CI) | AUC (95% CI) | OR (95% CI) | p value | RR (95% CI) | AUC (95% CI) |
| UmA PI >95th centile (Schaffer and Staudach 1997) | n=122 | 6.7 (2.7-16.9) | 0.0001 | 3.9 (1.7-8.7) | 0.70 (0.63-0.78) | 8.2 (3.5-18.8) | <0.00005 | 3.3 (1.7-6.4) | 0.73 (0.66-0.81) |
| Absent or reversed UmA EDF | n=122 | 6.5 (2.8-15.0) | <0.00005 | 3.2 (1.7-6.0) | 0.71 (0.63-0.80) | 14.2 (5.5-36.8) | <0.00005 | 3.1 (1.8-5.4) | 0.76 (0.69-0.84) |
| UmA category[1] | n=122 | 2.8 (1.1-4.2) | <0.00005 | 1.8 (1.4-2.4) | 0.77 (0.69-0.86) | 3.5 (2.2-5.6) | <0.00005 | 1.7 (1.3-2.1) | 0.80 (0.72-0.88) |
| MCA PI <5th centile (Schaffer and Staudach 1997) | n=117 | 1.8 (0.64-5.10) | 0.27 | 1.4 (0.66-3.1) | | 2.2 (0.77-6.5) | 0.14 | 1.4 (0.74-2.8) | |
| Absent or reversed DV a wave | n=119 | 19.5 (2.3-162.3) | 0.006 | 3.1 (1.4-6.6) | 0.59 (0.53-0.66) | 4.4 (0.87-21.9) | 0.07 | 1.7 (0.79-3.9) | |
| Mean UtA PI | n=122 | 2.3 (1.1-4.7) | 0.017 | 1.7 (1.0-2.9) | 0.64 (0.53-0.75) | 4.1 (1.9-8.6) | 0.0002 | 1.9 (1.2-3.0) | 0.72 (0.62-0.81) |
| Mean UtA PI >95th centile (Schaffer and Staudach 1997) | n=122 | 2.2 (0.76-6.4) | 0.15 | 1.8 (0.69-4.5) | | 8.6 (2.4-30.7) | 0.0009 | 4.4 (1.4-14.1) | 0.64 (0.57-0.70) |
| $EFW_{HM}$ z-score (Hadlock et al. 1985, Marsal et al. 1996) | n=123 | 0.11 (0.048-0.25) | <0.00005 | 0.50 (0.39-0.65) | 0.85 (0.78-0.92) | 0.32 (0.18-0.56) | <0.00005 | 0.67 (0.52-0.86) | 0.64 (0.57-0.70) |
| $EFW_{I}$ z-score (Stirnemann et al. 2017) | n=123 | 0.29 (0.18-0.47) | <0.00005 | 0.84 (0.79-0.90) | 0.87 (0.80-0.94) | 0.53 (0.39-0.71) | <0.00005 | 0.89 (0.83-0.96) | 0.74 (0.66-0.83) |
| Slow fetal growth[2] | n=104 | 4.0 (1.68-9.74) | 0.0018 | 2.6 (1.3-5.2) | 0.67 (0.57-0.77) | 2.3 (1.0-5.1) | 0.049 | 1.6 (0.87-3.0) | 0.60 (0.50-0.70) |

[1]Four levels: ≤95th centile, >95th centile with positive EDF, absent EDF, reversed EDF. [2]Slow fetal growth, defined as a worsening of percentage weight deviation by ≥10 percentage points over 2 weeks or equivalent (Marsal 2009), was assessed over a minimum of 2 weeks so included data from follow-up scans. DV=ductus venosus, EDF=end-diastolic flow, $EFW_{HM}$=estimated fetal weight calculated using Hadlock 3 formula with z-score calculated using Marsal

reference chart, $EFW_I$=estimated fetal weight and z-score calculated using Intergrowth formulae, MCA=middle cerebral artery, OR=odds ratio, PI=pulsatility index, RR=risk ratio (estimated using Poisson regression), UmA=umbilical artery, UtA=uterine artery.

Table 5: Abbreviations and full names of proteins from main article figures.

| Abbreviation | Full protein name | Figure | | | | |
|---|---|---|---|---|---|---|
| | | 2 | 3 | 7A | 7B | 8 |
| ADAMTS13 | A disintegrin and metalloproteinase with thrombospondin motifs 13 | | x | | | |
| ADM | Pro-adrenomedullin | | x | x | | |
| ACE2 | Angiotensin-converting enzyme 2 | | x | | | |
| AGRP | Agouti-related protein | | x | | | |
| AMBP | Protein AMBP | | | | | x |
| ANG1 | Angiopoietin-1 | | x | | | |
| BOC | Brother of cysteine dioxygenase | x | x | | | |
| CA5A | Carbonic anhydrase 5A, mitochondrial | | x | | | x |
| CALCA | Calcitonin | | | | x | |
| CALCRL | Calcitonin gene-related peptide type 1 receptor | | | | x | |
| CD40L | CD40 ligand | | x | | | x |
| CD47 | Leucocyte surface antigen CD47 | | | | x | |
| CEACAM8 | Carcinoembryonic antigen related cell adhesion molecule 8 | | x | | | |
| CHD5 | Chromodomain-helicase-DNA-binding protein 5 | | | | x | |
| CSH2 | Chorionic somatomammotropin hormone 2 | | | | x | |
| CTRC | Chymotrypsin C | | x | | | |
| CTSL1 | Cathepsin L1 | | x | | | |
| DCN | Decorin | | x | | | x |
| DECR1 | 2,4-dienoyl-CoA reductase, mitochondrial | | x | | | |
| DKK1 | Dickkopf-related protein 1 | | x | | | |
| FABP2 | Fatty acid-binding protein, intestinal | x | x | | | |
| FGF21 | Fibroblast growth factor 21 | x | x | | | x |
| FGF23 | Fibroblast growth factor 23 | | x | | | |
| FN | Fibronectin | | | | | x |
| FS | Follistatin | x | | | | x |
| GAL9 | Galectin-9 | x | | | | x |
| GDF2 | Growth/differentiation factor 2 | | x | | | x |
| GH | Growth hormone | | x | x | | x |
| GHR | Growth hormone receptor | | | | x | |
| GT / FABP6 | Gastrotropin | | x | | | |
| HAOX1 | Hydroxyacid oxidase 1 | | | x | | x |
| HPL / CSH1 | Human placental lactogen / chorionic somatomammotropin hormone 1 | x | x | x | | x |
| IL1RA | Interleukin-1 receptor antagonist protein | x | | | | |
| IL1RL2 | Interleukin-1 receptor-like 2 | x | x | | | x |
| IL4RA | Interleukin-4 receptor subunit alpha | | | | | |
| IL6 | Interleukin-6 | x | | | | |
| IL17D | Interleukin-17D | | x | x | | x |
| IL18 | Interleukin-18 | | x | | | |
| IL27 | Interleukin-27 | | x | | | |
| JAK2 | Tyrosine-protein kinase JAK2 | | | | x | |
| KIM1 | Kidney injury molecule 1 | | x | | | |
| LEP | Leptin | | x | | | |
| LNPEP | Leucyl-cystinyl aminopeptidase | | x | | | |
| LOX1 | Lectin-like oxidized LDL receptor 1 | x | x | | | x |
| MARCO | Macrophage receptor MARCO | x | x | x | | |
| MMP12 | Matrix metalloproteinase-12 | x | x | | | x |
| NEMO | NF-kappa-B essential modulator | | x | | | |
| NP1 | Neuropilin 1 | x | x | | x | |
| OSCAR | Osteoclast-associated immunoglobulin-like receptor | | x | | | |
| PAPPA | Pappalysin-1 / pregnancy-associated plasma protein A | x | x | | | |
| PAR1 | Proteinase-activated receptor 1 | | x | | | |

| | | | | | | |
|---|---|---|---|---|---|---|
| PDGFB | Platelet-derived growth factor subunit B | x | x | | | |
| PDL2 | Programmed cell death 1 ligand 2 | x | x | | | x |
| PlGF | Placental growth factor | x | x | x | | x |
| PLXNA1 | Plexin-A1 | | | | x | |
| PRELP | Prolargin | | x | | | |
| PRSS27 | Serine protease 27 | | x | | | |
| PSG1 | Pregnancy-specific beta-glycoprotein 1 | x | | x | | x |
| PTX3 | Pentraxin-related protein PTX3 | | x | | | |
| RAGE | Receptor for advanced glycosylation end products | | x | | | |
| RAMP1 | Receptor activity-modifying protein 1 | | | | x | |
| RAMP2 | Receptor activity-modifying protein 2 | | | | x | |
| RAMP3 | Receptor activity-modifying protein 3 | | | | x | |
| REN | Renin | | x | | | |
| SEMA3A | Semaphorin-3A | | | | x | |
| SERPINA12 | Serpin A12 | | x | | | |
| sflt1 | Soluble fms-like tyrosine kinase 1 | | | | x | x |
| SH2D2A | SH2 domain-containing protein 2A | | | | x | |
| SIRPG | Signal-regulatory protein gamma | | | | x | |
| SPON2 | Spondin-2 | | x | | | |
| SRC | Proto-oncogene tyrosine-protein kinase Src | | x | | | |
| STK4 | Serine/threonine-protein kinase 4 | x | x | | | |
| TF | Tissue factor | | x | | | |
| TGM2 | Protein-glutamine gamma-glutamyltransferase 2 | | x | | | |
| THBS1 | Thrombospondin-1 | | | | x | |
| THBS2 | Thrombospondin-2 | x | x | x | | x |
| TNFRSF10A | Tumor necrosis factor receptor superfamily member 10A | | | | | x |
| TNFRSF11A | Tumor necrosis factor receptor superfamily member 11A | | x | | | |
| TRAIL-R2 | TNF-related apoptosis-inducing ligand receptor 2 | | x | | | x |
| VEGF-B | Vascular endothelial growth factor B | | | | x | |
| VEGF-D | Vascular endothelial growth factor D | | | | x | |
| VEGFR2 | Vascular endothelial growth factor receptor 2 | x | x | | x | |
| VSIG2 | V-set and immunoglobulin domain-containing protein 2 | | x | | | |

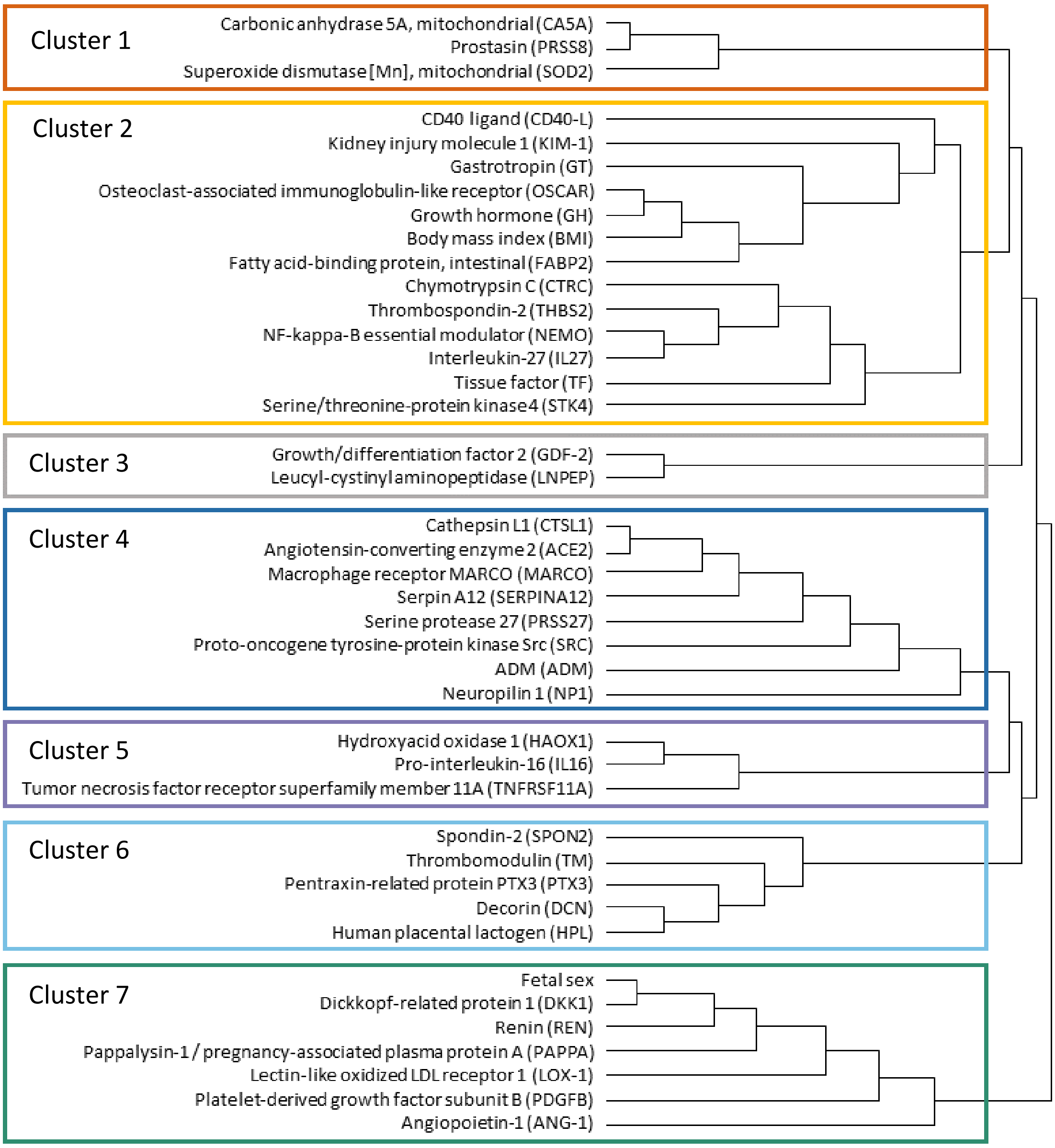

Figure 2: Dendrogram to accompany network analysis for pregnancies ending in fetal or neonatal death versus neonatal survival to 29 days of life, based on edge betweenness analysis.

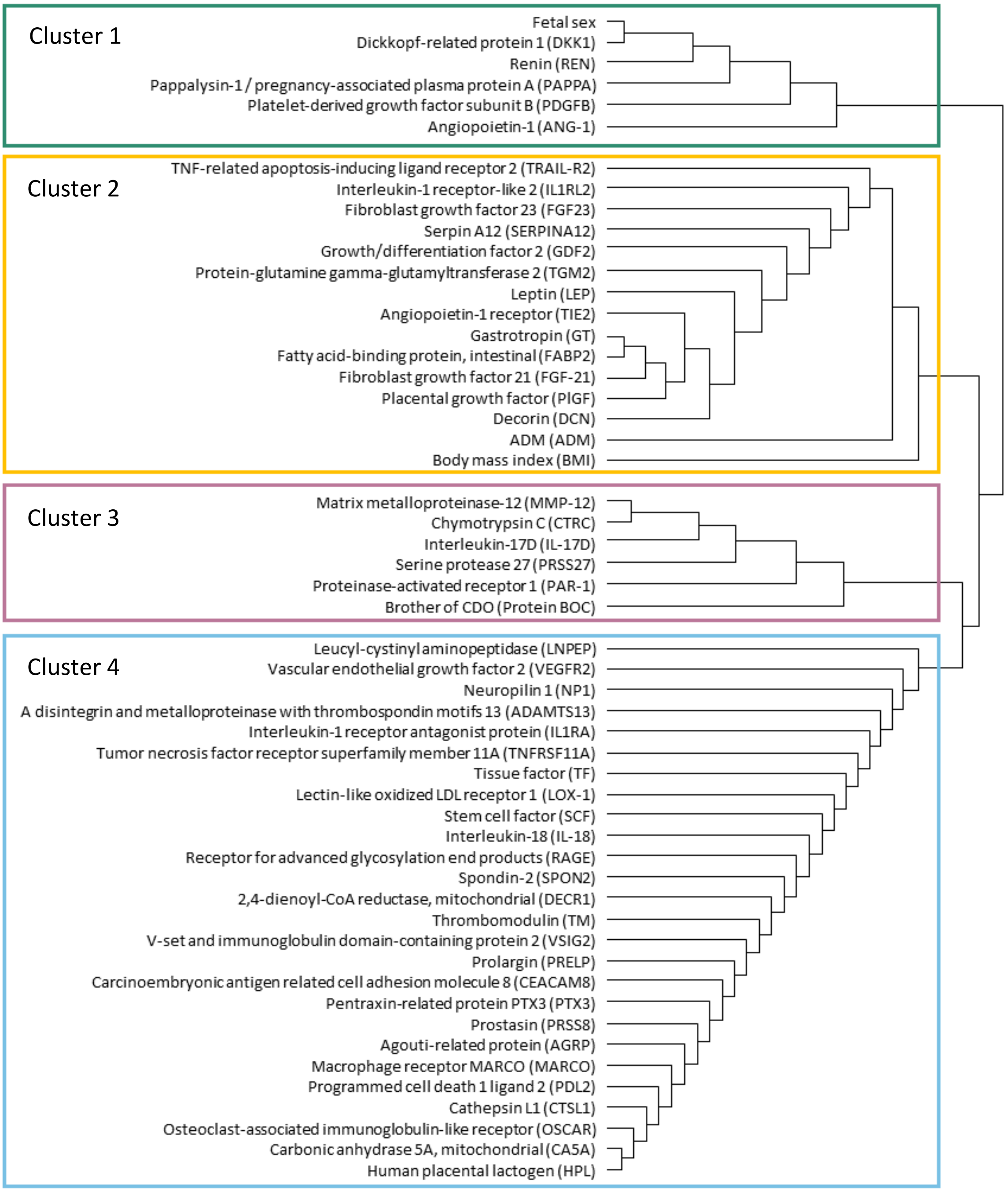

Figure 3: Dendrogram to accompany network analysis for pregnancies ending in fetal death or delivery ≤28+0 weeks of gestation versus continuation of pregnancy beyond 28+0 weeks, based on edge betweenness analysis.

(A)

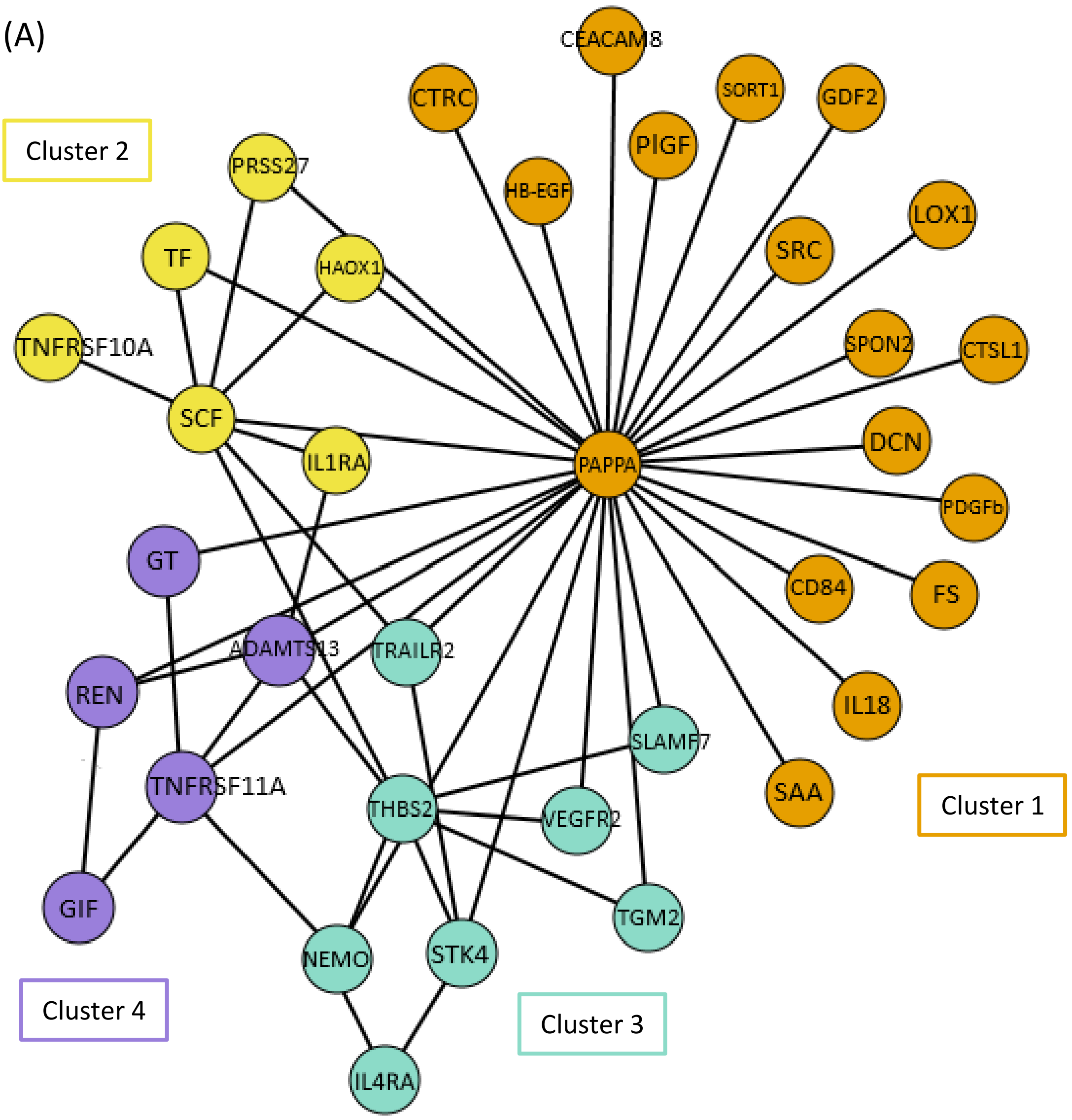

Cluster 2
CEACAM8
CTRC
SORT1
GDF2
PIGF
PRSS27
HB-EGF
LOX1
TF
HAOX1
SRC
TNFRSF10A
SPON2
CTSL1
SCF
DCN
IL1RA
PAPPA
PDGFb
GT
CD84
FS
ADAMTS13
TRAILR2
REN
IL18
SLAMF7
TNFRSF11A
SAA
Cluster 1
THBS2
VEGFR2
GIF
TGM2
NEMO
STK4
Cluster 4
Cluster 3
IL4RA

(B)

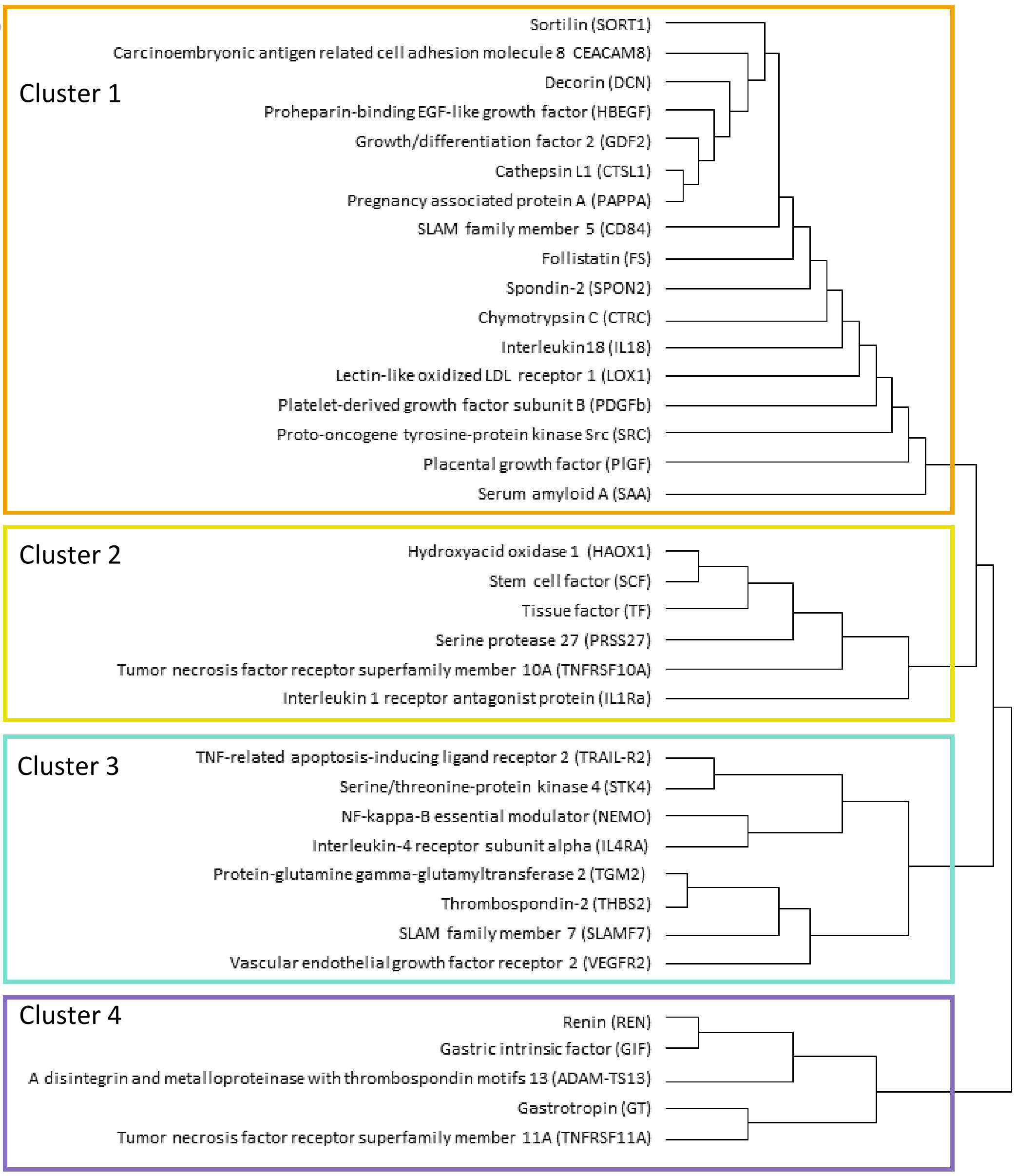

Figure 4: Network analysis for pregnancies that developed an abnormal UmA pulsatility index >95$^{th}$ centile versus those that did not. (A) Parenclitic protein network and clustering (B) Dendrogram, based on edge betweenness analysis.

(A)

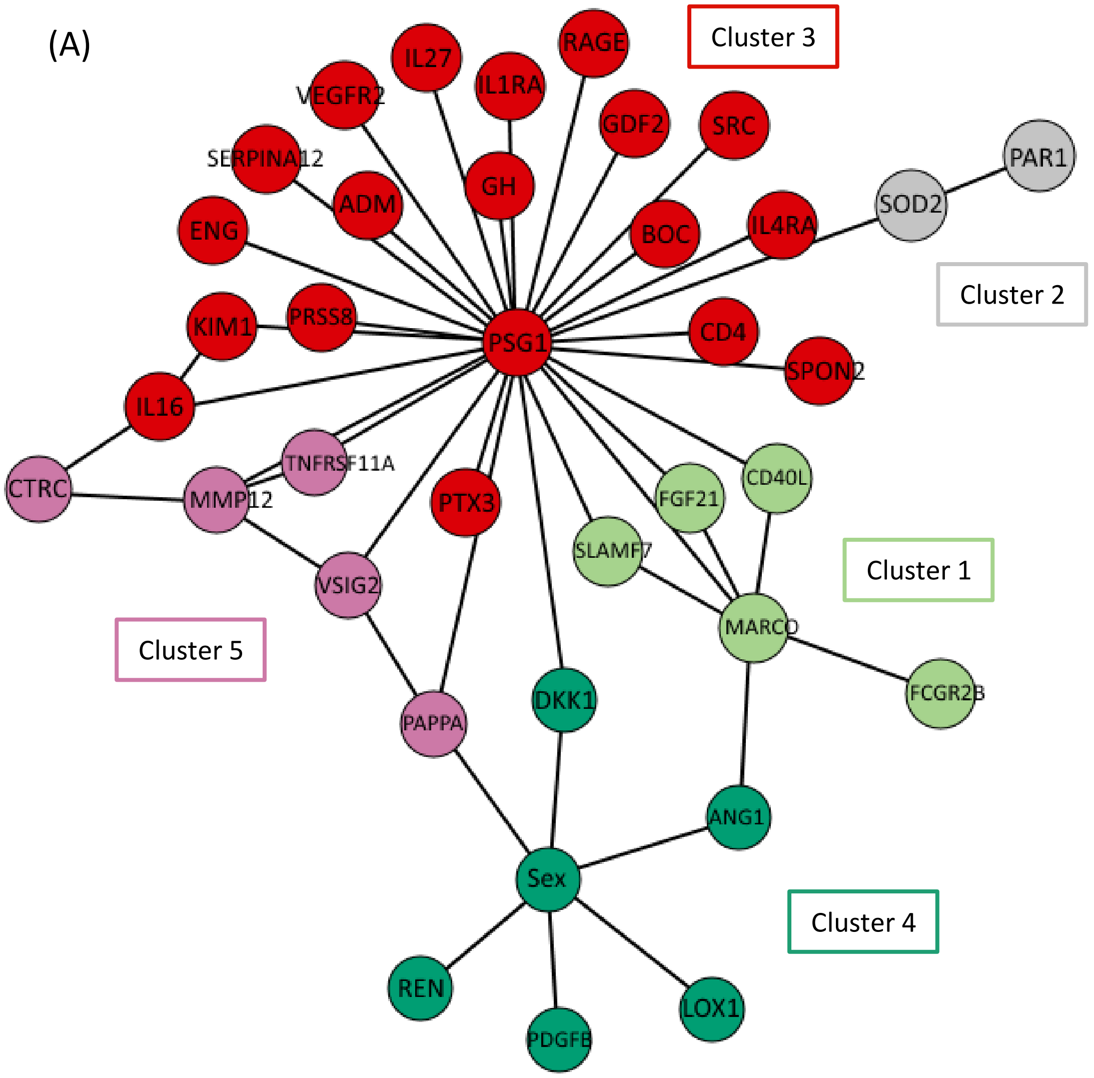
Cluster 3
Cluster 2
Cluster 1
Cluster 5
Cluster 4
IL27
RAGE
IL1RA
VEGFR2
GDF2
SRC
PAR1
SERPINA12
GH
ADM
SOD2
ENG
BOC
IL4RA
KIM1
PRSS8
PSG1
CD4
SPON2
IL16
CTRC
MMP12
TNFRSF11A
PTX3
FGF21
CD40L
SLAMF7
VSIG2
MARCO
FCGR2B
DKK1
PAPPA
ANG1
Sex
REN
PDGFB
LOX1

(B)

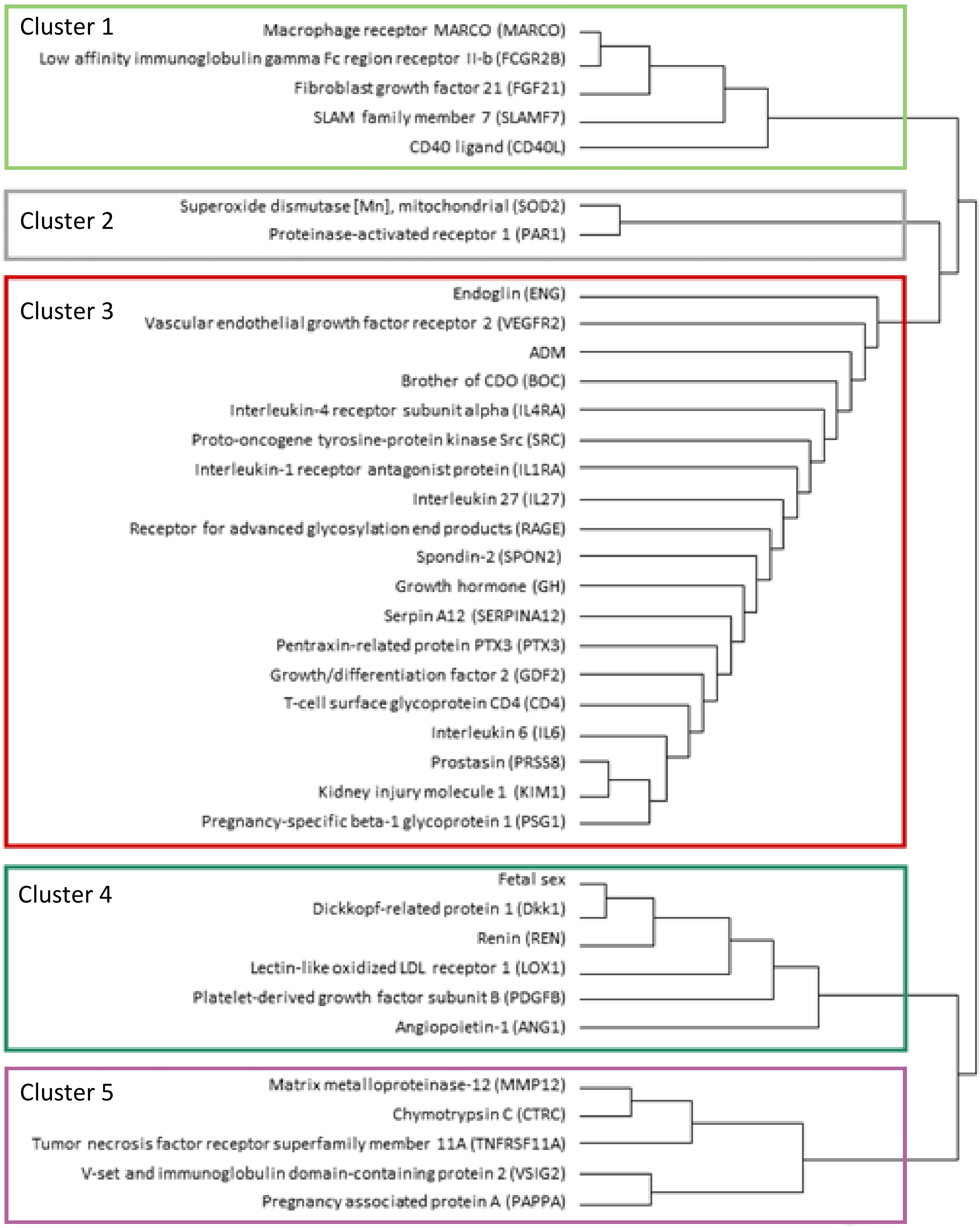

Figure 5: Network analysis for pregnancies with slow fetal growth versus those without slow fetal growth. (A) Parenclitic protein network and clustering (B) Dendrogram, based on edge betweenness analysis. Slow fetal growth was defined as a worsening of weight deviation of >10 percentage points over a two-week interval or equivalent trajectory over a longer period (Marsal 2009).

Table 6: Experience and geographical origin of the 45 clinicians completing the marker priority survey.

| **Specialty** | |
|---|---|
| | n (%) |
| Consultant in Maternal and Fetal Medicine | 31 (69) |
| Consultant in Obstetrics and Gynaecology | 8 (18) |
| Other | 6 (13) |

| **Location** | |
|---|---|
| | n (%) |
| Europe | 21 (47) |
| Australia and New Zealand | 6 (13) |
| Asia and Middle East | 5 (11) |
| North and South America | 4 (36) |

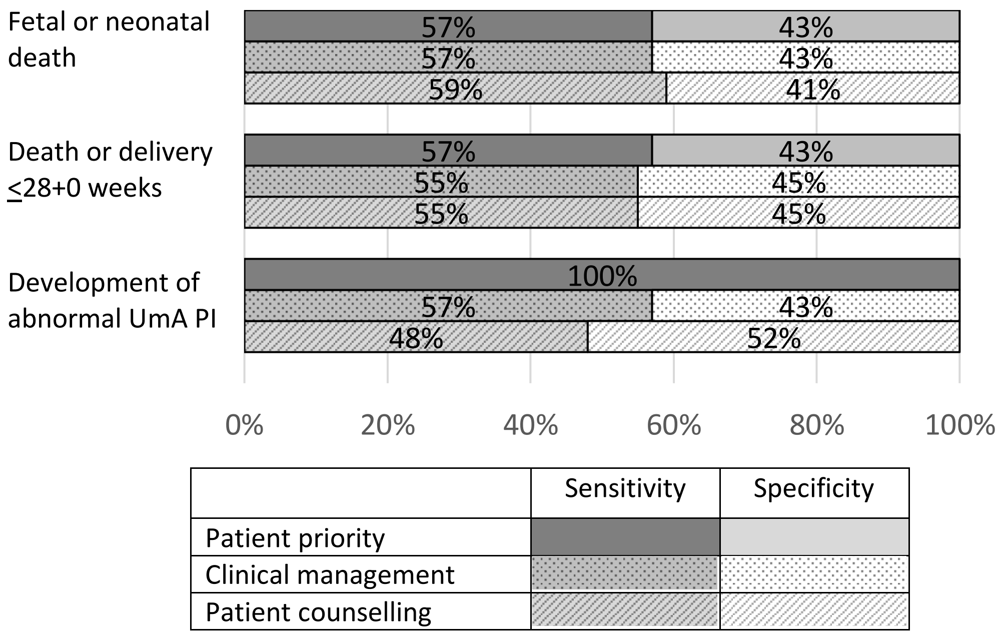

Figure 6: The proportion of patients who prioritised sensitivity or specificity for the three outcomes and the proportion of clinicians who prioritised sensitivity or specificity for clinical management and for patient counselling for the three outcomes. UmA PI=umbilical artery pulsatility index.

Table 7: Models significantly improved by the addition of gestational age or pre-eclampsia at enrolment. Likelihood ratio (LR) tests, Akaike information criteria (AIC) and Schwarz's Baysian information criteria (BIC) showing the effects of the added variables.

| Outcome | Variable(s) | LR test p value | AIC | BIC |
|---|---|---|---|---|
| Fetal or neonatal death | $EFW_{HM}$ z-score | 0.0001 | 111.2 | 116.9 |
| | $EFW_{HM}$ z-score & gestational age | | 97.2 | 105.6 |
| | $EFW_{HM}$ z-score & UmA category[1] | <0.00005 | 107.7 | 116.1 |
| | $EFW_{HM}$ z-score & UmA category & gestational age[1] | | 90.7 | 101.9 |
| Death or delivery ≤28+0 weeks of gestation | PlGF | 0.013 | 128.9 | 134.5 |
| | PlGF & pre-eclampsia | | 124.7 | 133.1 |
| | PlGF & PSG1* | 0.0042 | 119.3 | 127.7 |
| | PlGF, PSG1 & pre-eclampsia | | 113.1 | 124.3 |
| | UmA category[1] | 0.0002 | 131.5 | 137.1 |
| | UmA category & pre-eclampsia[1] | | 119.5 | 127.9 |
| | UmA category & PlGF[1] | 0.013 | 107.4 | 115.8 |
| | UmA category, PlGF & pre-eclampsia[1] | | 103.2 | 114.4 |

[1]n=1 missing from validation set. $EFW_{HM}$=estimated fetal weight calculated using Hadlock 3 formula (Hadlock et al. 1985) with z-score calculated using Marsal reference chart (Marsal et al. 1996), $EFW_{Int}$=estimated fetal weight and z-score calculated using Intergrowth formulae (Stirnemann et al. 2017), PlGF=placental growth factor, PSG1=pregnancy-specific glycoprotein 1, UmA=umbilical artery.

Table 8: Details of the validated models to predict fetal or neonatal death. Constants and coefficients for calculating log(odds) provided, along with variable cut points that give the maximum positive likelihood ratio (LR+), maximum correct classification and minimum negative likelihood ratio (LR-).

| Variable(s) | Constant (95% CI) | Coefficient(s) (95% CI) | Cut point | Value of the variable | Correctly classified | LR+ | LR- | Sensitivity | Specificity |
|---|---|---|---|---|---|---|---|---|---|
| PlGF | 4.20 (2.21 to 6.19) | ln(PlGF): -1.43 (-2.04 to -0.826) | Max LR+ | <14.2 pg/ml | 80% | 18.3 | 0.56 | 45% | 98% |
| | | | Max correct | | | | | | |
| | | | Min LR- | <240 pg/ml | 43% | 1.2 | 0.00 | 100% | 14% |
| PlGF & lymphotactin | 4.09 (2.07 to 6.10) | ln(PlGF): -1.41 (-2.03 to -0.795) lymphotactin: 0.442 (-0.061 to 0.945) | Max LR+ | | 70% | 11.6 | 0.87 | 14% | 99% |
| | | | Max correct | | 80% | 7.4 | 0.49 | 55% | 93% |
| | | | Min LR- | | 47% | 1.2 | 0.00 | 100% | 20% |
| $EFW_{HM}$ z-score | -8.02 (-10.83 to -5.20) | $EFW_{HM}$ z: -2.21 (-3.03 to -1.38) | Max LR+ | <-4.10 | 78% | 30.9 | 0.63 | 38% | 99% |
| | | | Max correct | <-3.46 | 81% | 5.6 | 0.35 | 69% | 88% |
| | | | Min LR- | <-2.25 | 44% | 1.2 | 0.00 | 100% | 15% |
| $EFW_{HM}$ z-score & gestational age at enrolment | 5.36 (-2.05 to 12.78) | $EFW_{HM}$ z: -2.69 (-3.70 to -1.68) GA: -0.915 (-0.141 to -0.042) | Max LR+ | | 78% | 30.9 | 0.63 | 38% | 99% |
| | | | Max correct | | 85% | 8.3 | 0.31 | 71% | 91% |
| | | | Min LR- | | 47% | 1.2 | 0.00 | 100% | 20% |
| $EFW_{Int}$ z-score | -5.72 (-7.73 to -3.72) | $EFW_{Int}$ z: -1.24 (-1.71 to -0.76) | Max LR+ | <-5.16 | 84% | 44.4 | 0.46 | 55% | 99% |
| | | | Max correct | | | | | | |
| | | | Min LR- | <-1.68 | 9% | 1.1 | 0.00 | 100% | 9% |
| $EFW_{HM}$ z-score & UmA category[1] | -7.30 (-10.16 to -4.45) | $EFW_{HM}$ z: -1.79 (-2.67 to -0.910) UmA: 0.56 (0.083 to 1.04) | Max LR+ | | 82% | 20.7 | 0.50 | 51% | 98% |
| | | | Max correct | | 84% | 6.8 | 0.27 | 76% | 89% |
| | | | Min LR- | | 42% | 1.1 | 0.00 | 100% | 12% |
| $EFW_{HM}$ z-score & UmA category & gestational age at enrolment[1] | 8.45 (0.264 to 16.63) | $EFW_{HM}$ z: -2.27 (-3.34 to -1.21) UmA: 0.784 (0.229 to 1.34) GA: -0.108 (-0.164 to -0.520) | Max LR+ | | 83% | 41.5 | 0.49 | 51% | 99% |
| | | | Max correct | | 85% | 5.7 | 0.17 | 85% | 85% |
| | | | Min LR- | | 48% | 1.3 | 0.00 | 100% | 21% |

[1]n=1 missing from validation set. PlGF=placental growth factor, UmA=umbilical artery. Models generated using the natural log of PlGF in pg/ml but cut points converted back to concentration. Models generated using centered and scaled values of lymphotactin normalised protein expression.

Table 9: Details of the validated models to predict death or delivery ≤28+0 weeks of gestation. Constants and coefficients for calculating log(odds) and variable cut points that give the maximum positive likelihood ratio (LR+), maximum correct classification and minimum negative likelihood ratio (LR-).

| Variable(s) | Constant (95% CI) | Coefficient(s) (95% CI) | Cut point | Value of variable | Correctly classified | LR+ | LR- | Sensitivity | Specificity |
|---|---|---|---|---|---|---|---|---|---|
| PlGF | 5.17 | ln(PlGF): -1.50 (-2.07 to -0.924) | Max LR+ | <14.5 pg/ml | 70% | 24.7 | 0.63 | 38% | 98% |
| | (3.17 to 7.17) | | Max correct | <34 pg/ml | 76% | 3.2 | 0.30 | 78% | 75% |
| | | | Min LR- | <240 pg/ml | 56% | 1.2 | 0.00 | 100% | 17% |
| PlGF & pre-eclampsia | 4.58 | ln(PlGF): -1.38 (-1.95 to -0.81) | Max LR+ | | 72% | 26.9 | 0.60 | 41% | 98% |
| | (2.57 to 6.58) | Pre-eclampsia: 2.13 (2.57 to 4.24) | Max correct | | 76% | 3.1 | 0.32 | 76% | 76% |
| | | | Min LR- | | 56% | 1.2 | 0.00 | 100% | 17% |
| PlGF & PSG1 | 4.47 | ln(PlGF): -1.30 (-1.88 to -0.714) | Max LR+ | | 67% | 21.3 | 0.68 | 33% | 98% |
| | (2.38 to 6.56) | PSG1: -0.846 (-1.37 to -0.318) | Max correct | | 84% | 4.4 | 0.15 | 88% | 80% |
| | | | Min LR- | | 53% | 1.1 | 0.00 | 100% | 11% |
| PlGF, PSG1 & pre-eclampsia | 3.66 | ln(PlGF): -1.13 (-1.71 to -0.55) | Max LR+ | | 74% | 30.3 | 0.54 | 47% | 98% |
| | (1.55 to 5.78) | PSG1: -0.95 (-1.51 to -0.40) | Max correct | | 83% | 4.3 | 0.17 | 86% | 80% |
| | | Pre-eclampsia: 2.91 (0.30 to 5.53) | Min LR- | | 53% | 1.1 | 0.00 | 100% | 11% |
| UmA category[1] | -1.42 | UmA: 1.26 (0.81 to 1.72) | Max LR+ | UmA PI >95th centile | 63% | 8.0 | 0.78 | 25% | 97% |
| | (-2.03 to -0.81) | | Max correct | Absent or reversed EDF | 77% | 5.9 | 0.41 | 63% | 89% |
| | | | Min LR- | Reversed EDF | 73% | 2.4 | 0.29 | 81% | 66% |
| UmA category & pre-eclampsia[1] | -1.76 | UmA: 1.32 (0.83 to 1.81) | Max LR+ | | 69% | 12.0 | 0.65 | 37% | 97% |
| | (-2.45 to -1.06) | Pre-eclampsia: 3.22 (1.02 to 5.43) | Max correct | | 81% | 6.0 | 0.30 | 74% | 88% |
| | | | Min LR- | | 75% | 2.4 | 0.22 | 86% | 65% |
| UmA category & PlGF[1] | 3.49 | UmA: 1.06 (0.58 to 1.54) | Max LR+ | | 71% | 26.2 | 0.61 | 40% | 98% |
| | (1.29 to 5.70) | ln(PlGF): -1.35 (-1.98 to -0.718) | Max correct | | 83% | 4.3 | 0.18 | 86% | 80% |
| | | | Min LR- | | 71% | 1.9 | 0.00 | 100% | 46% |
| UmA category, PlGF & pre-eclampsia[1] | 2.66 | UmA: 1.11 (0.61 to 1.61) | Max LR+ | | 75% | 30.8 | 0.53 | 47% | 98% |
| | (0.38 to 4.93) | ln(PlGF): -1.18 (-1.81 to -0.55) | Max correct | | 84% | 4.5 | 0.13 | 89% | 80% |
| | | Pre-eclampsia: 2.38 (0.11 to 4.65) | Min LR- | | 71% | 1.9 | 0.00 | 100% | 46% |

[1]n=1 missing from validation set. EDF=end-diastolic flow, PI=pulsatility index, PlGF=placental growth factor, PSG1=pregnancy-specific glycoprotein 1, UmA=umbilical artery. Models generated using the natural log of PlGF in pg/ml but cut points converted back to concentration. Models generated using and centered and scaled values of PSG1. UmA PI centile calculated using Schaffer and Staudach 1997.

Table 10: Details of the validated models to predict development of umbilical artery pulsatility index >95th centile. Constants and coefficients for calculating log(odds) provided, along with variable cut points that give the maximum positive likelihood ratio (LR+), maximum correct classification and minimum negative likelihood ratio (LR-).

| Protein(s) | Constant (95% CI) | Coefficient (95% CI) | Cut point | PlGF concentration (pg/ml) | Correctly classified | LR+ | LR- | Sensitivity | Specificity |
|---|---|---|---|---|---|---|---|---|---|
| PlGF | 4.86 (1.49 to 8.23) | -1.17 (-1.94 to -0.390) | Max LR+ | <24 | 63% | 5.9 | 0.79 | 24% | 96% |
| | | | Max correct | <60 | 74% | 3.0 | 0.38 | 71% | 76% |
| | | | Min LR- | <270 | 59% | 1.3 | 0.00 | 100% | 24% |
| PlGF & fibronectin | 7.09 (1.94 to 12.24) | ln(PlGF): -1.44 (-2.38 to -0.503) Fibronectin: -0.0035 (-0.0091 to 0.0021) | Max LR+ | | 63% | 5.6 | 0.79 | 24% | 96% |
| | | | Max correct | | 76% | 3.2 | 0.31 | 76% | 76% |
| | | | Min LR- | | 61% | 1.4 | 0.00 | 100% | 28% |

PlGF=placental growth factor. Models generated using the natural log of PlGF in pg/ml but cut points converted back to concentration.

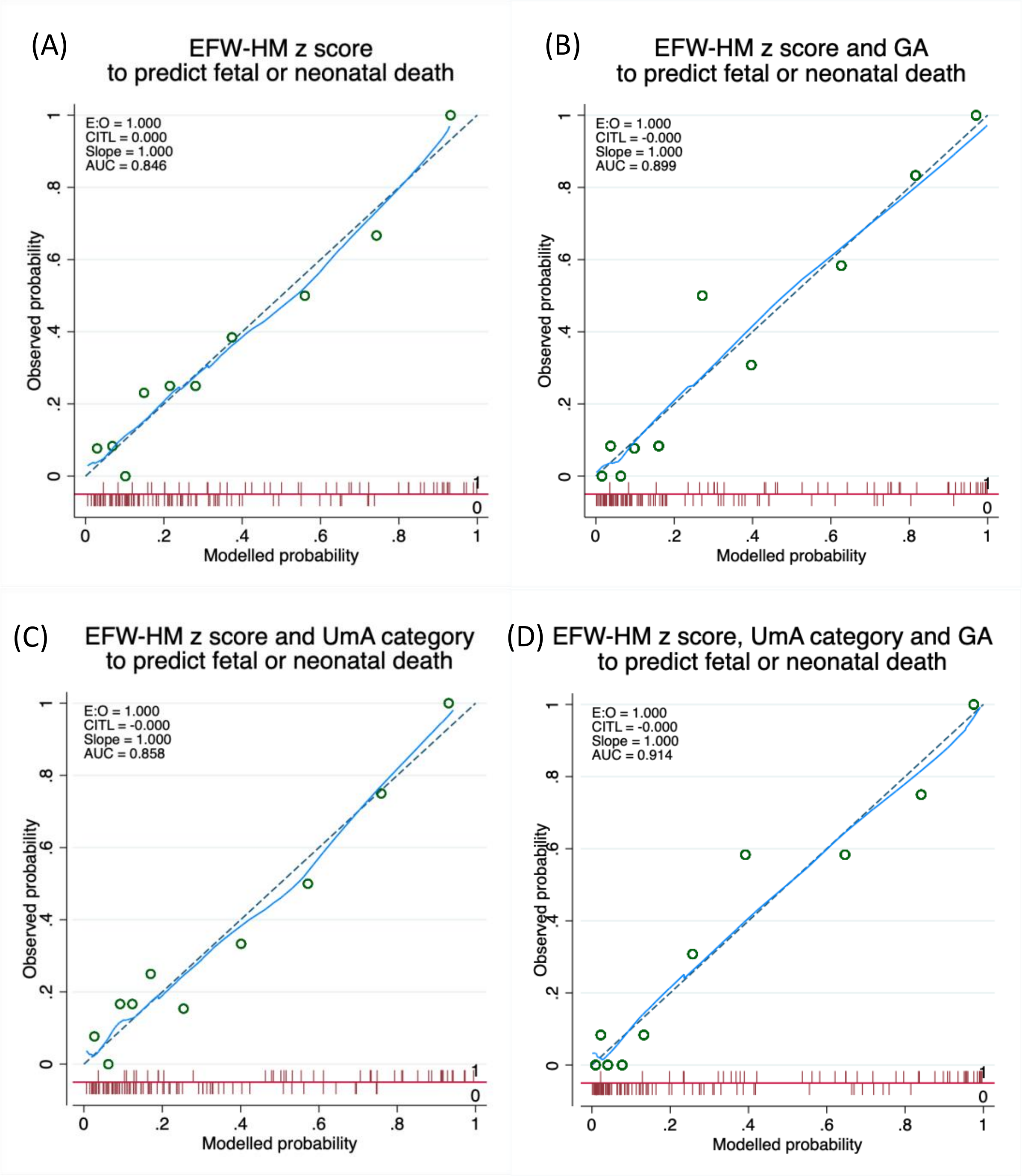

Figure 7: Calibration plots for the models in Tables 8-10. Green circles = grouped modelled probability plotted against grouped observed probability, solid blue line = locally weighted scatterplot smoothing (LOWESS), dashed grey line = line of unity. Spike plot (red) at the bottom of the chart area shows distribution of modelled probability by outcome (above line 1=fetal or neonatal death, below line 0=live birth and neonatal survival). CITL=calibration-in-the-large, EFW-HM=estimated fetal weight calculated using Hadlock 3 formula (Hadlock et al. 1985) with z-score calculated using Marsal reference chart (Marsal et al. 1996), E:O=ratio of expected to observed, GA=gestational age at study enrolment, UmA=umbilical artery.

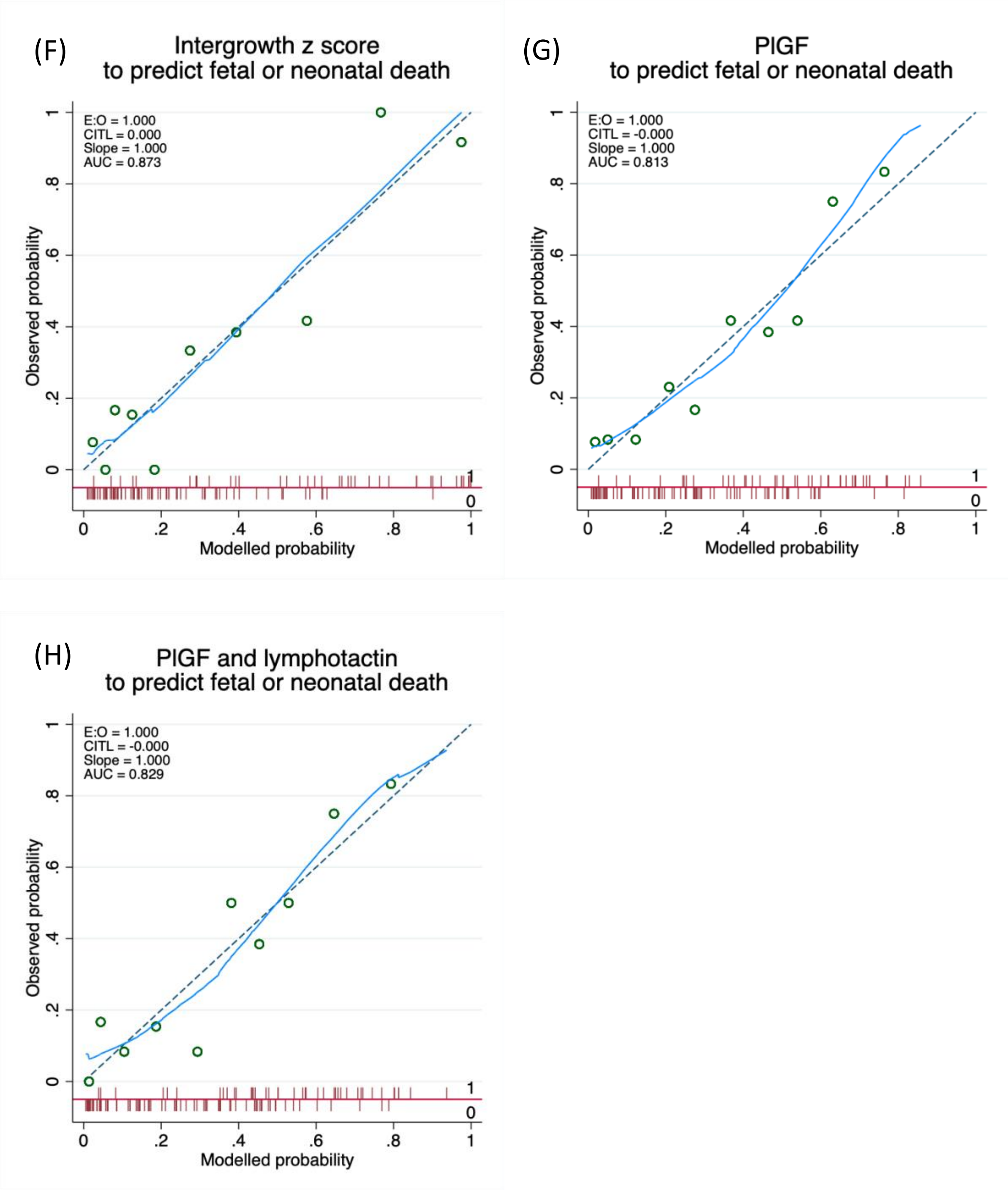

Figure 7 continued

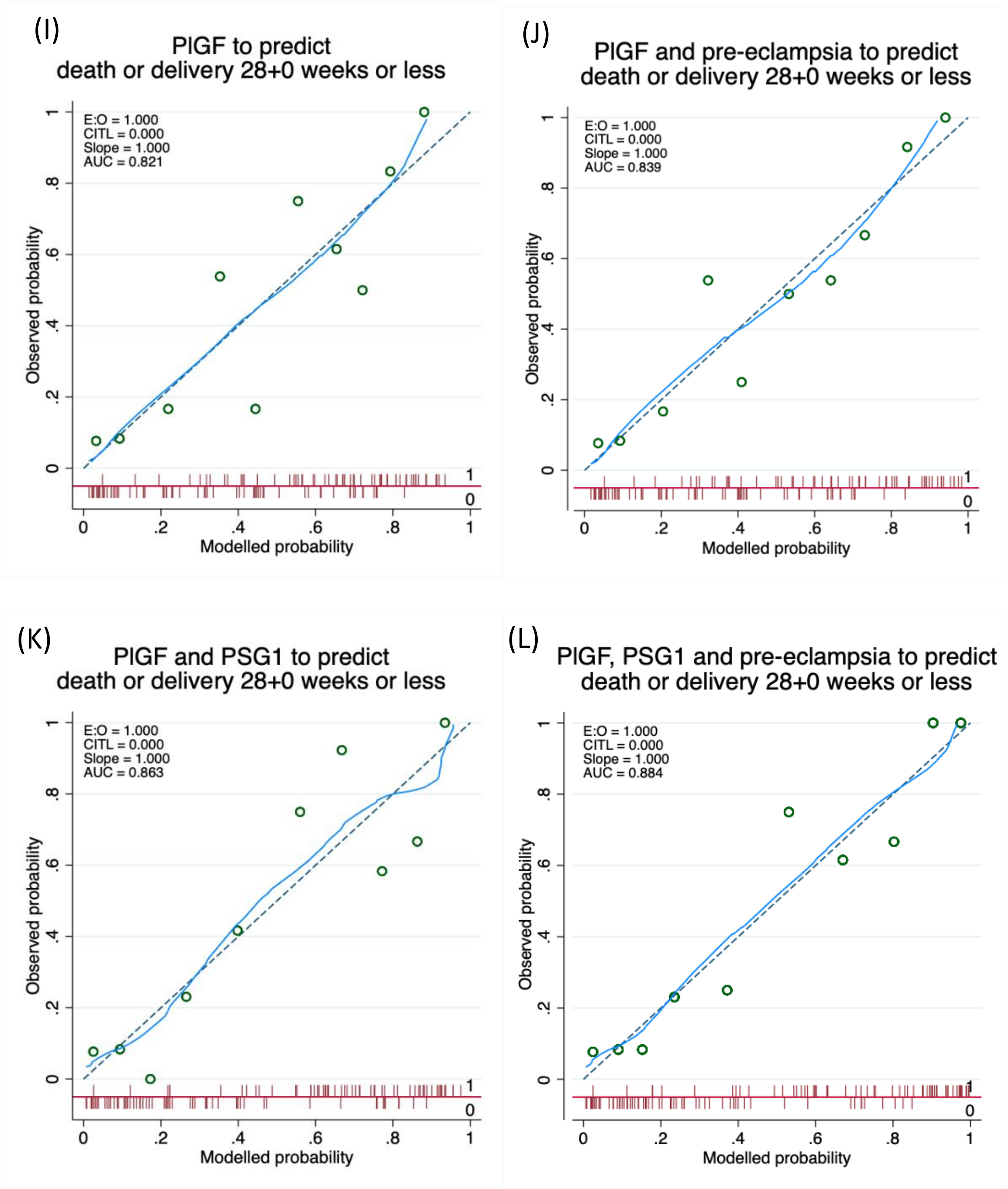

Figure 7 continued

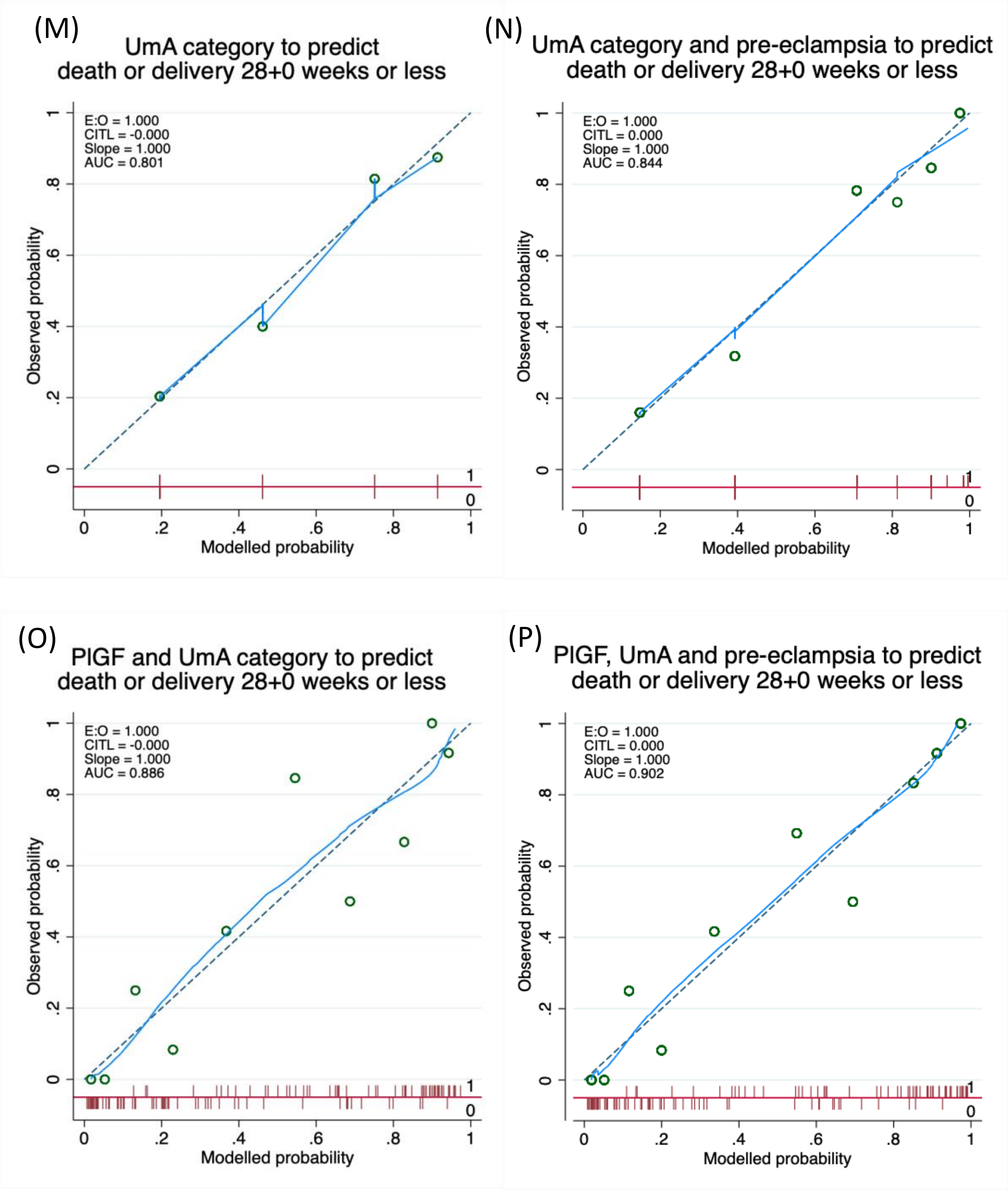

Figure 7 continued

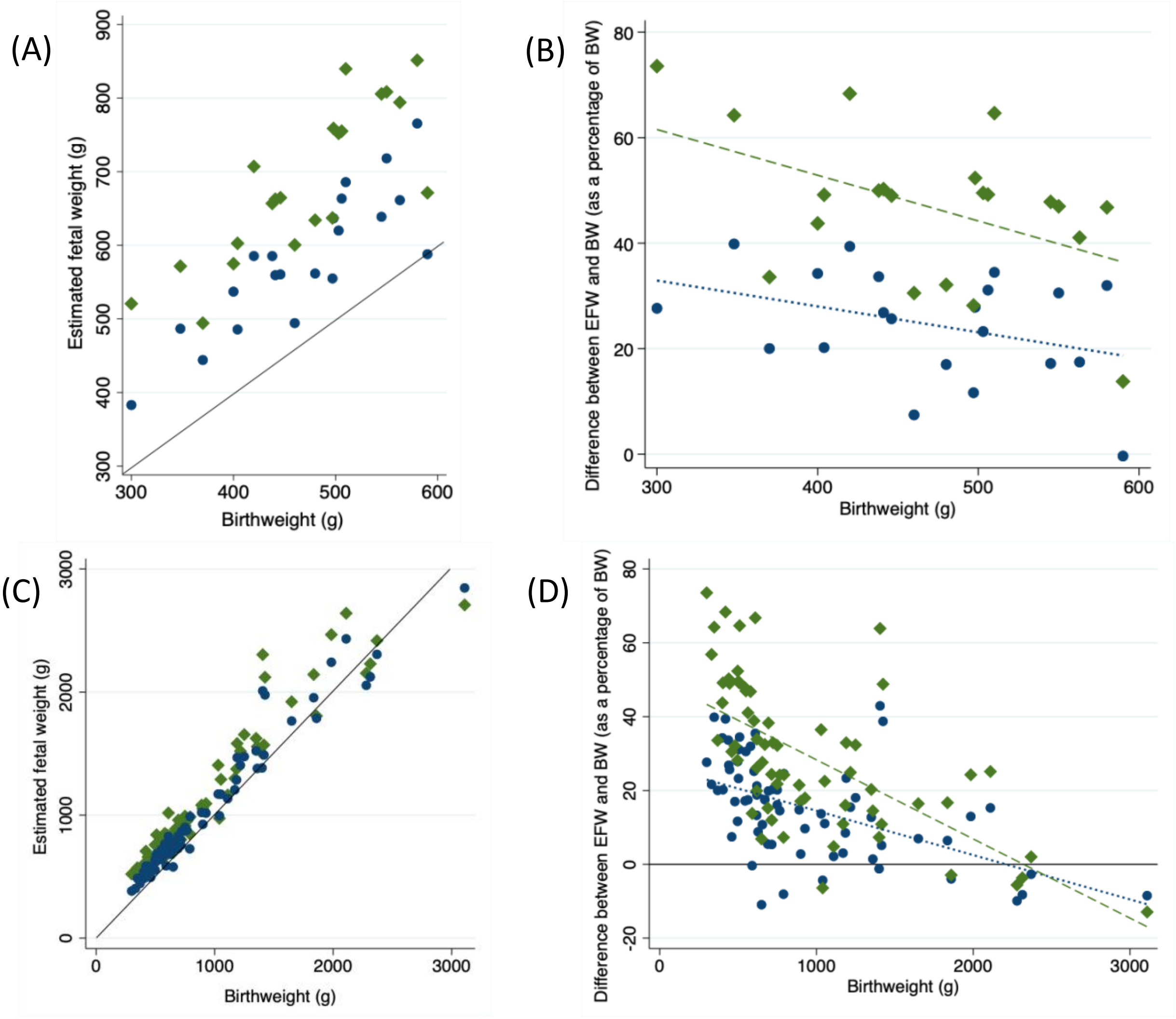

Figure 8: The differences between estimated fetal weights (EFWs) generated using the Intergrowth formula (green diamonds) (Stirnemann et al. 2017) and the Hadlock 3 formula (blue circles) (Hadlock et al. 1985) and birthweight (BW) for livebirths with an EFW performed within seven days of delivery. (A) Absolute differences for n=21 pregnancies with a BW <600g (B) percentage difference, as a percentage of BW, for n=21 pregnancies with a BW <600g (C) absolute differences for all n=67 pregnancies (D) percentage difference, as a percentage of BW, for all n=67 pregnancies. Solid black lines represent EFW=BW, green dashed lines= fitted association between BW and difference between Intergrowth EFW and BW as a percentage of BW, blue dotted line = fitted association between BW and difference between Hadlock 3 EFW and BW as a percentage of BW.

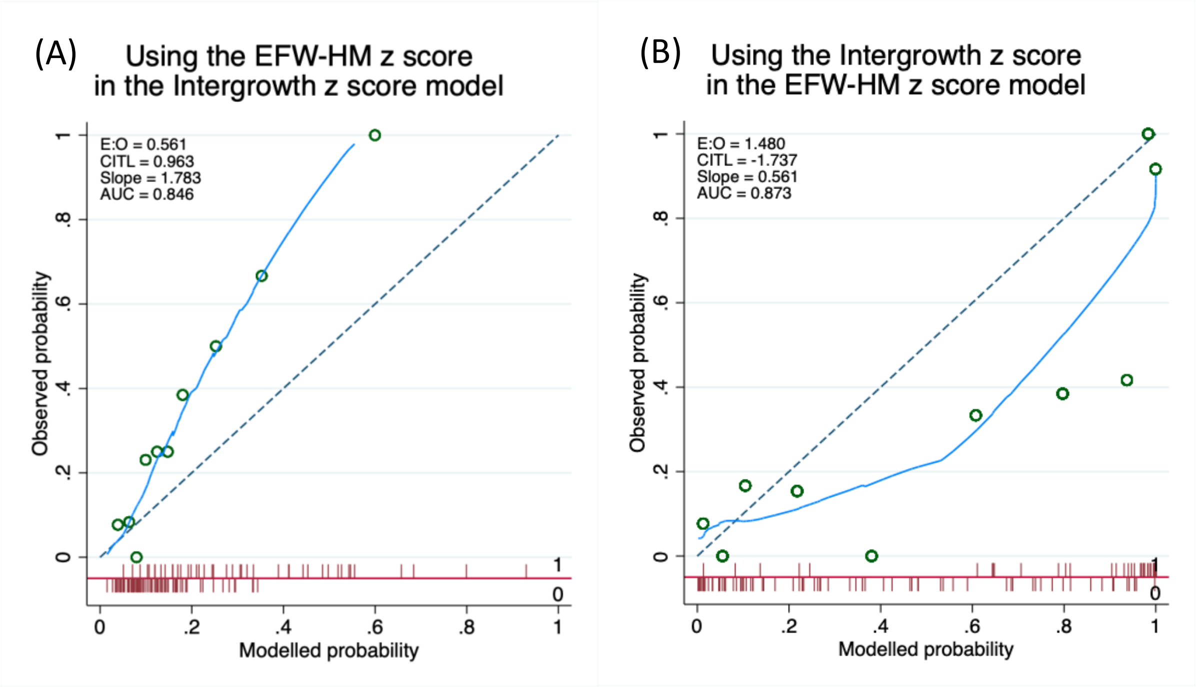

Figure 9: Calibration plots showing (A) the effect of using the estimated fetal weight calculated with the Hadlock 3 formula and z-score from the Marsal reference chart in the model generated from the Intergrowth z-score, and (B) the effect of using the Intergrowth z-score in the model generated from the Hadlock 3 formula estimated fetal weight z-score from the Marsal chart. Green circles = grouped modelled probability plotted against grouped observed probability, solid blue line = locally weighted scatterplot smoothing (LOWESS), dashed grey line = line of unity. Spike plot (red) at the bottom of the chart area shows distribution of modelled probability by outcome (above line 1=fetal or neonatal death, below line 0=live birth and neonatal survival). CITL=calibration-in-the-large, EFW-HM=estimated fetal weight calculated using Hadlock 3 formula (Hadlock et al. 1985) with z-score calculated using Marsal reference chart (Marsal et al. 1996), E:O=ratio of expected to observed. Intergrowth z-score from Stirnemann et al. 2017.

Table 11: Proteins showing an association with pregnancy outcomes at a Benjamini Hochberg 5% false discovery rate and Benjamini Hochberg 1% false discovery rate (over-shaded in blue) in centered and scaled data from the combined discovery and validation sets.

| **Protein** | **Abbreviation** | **Fetal or neonatal death -log10(p value)** | **Death or delivery <u><</u>28+0 weeks of gestation -log10(p value)** |
|---|---|---|---|
| Placental growth factor | PlGF | 7.86 | 10.52 |
| Human placental lactogen | HPL | 6.87 | 9.84 |
| Pro-adrenomedullin | ADM | 4.02 | |
| Pregnancy-specific beta-1 glycoprotein | PSG1 | 3.42 | 5.09 |
| Thrombospondin-2 | THBS2 | 3.37 | 5.04 |
| Hydroxyacid oxidase 1 | HAOX1 | 2.94 | 2.26 |
| Interleukin-17D | IL17D | 2.66 | 2.26 |
| Growth hormone | GH | 2.51 | 2.38 |
| Macrophage receptor MARCO | MARCO | 2.51 | |
| Programmed cell death 1 ligand 2 | PDL2 | | 4.71 |
| Matrix metalloproteinase 12 | MMP12 | | 4.31 |
| Soluble FMS-like tyrosine kinase 1 | sflt1 | | 3.31 |
| Galectin-9 | GAL9 | | 3.23 |
| Decorin | DCN | | 3.20 |
| Interleukin-1 receptor-like 2 | IL1RL2 | | 2.99 |
| Carbonic anhydrase 5A | CA5A | | 2.94 |
| TNF-related apoptosis-inducing ligand receptor 2 | TRAILR2 | | 2.75 |
| CD40 ligand | CD40L | | 2.59 |
| Fibroblast growth factor 21 | FGF21 | | 2.44 |
| Lectin-like oxidized LDL receptor 1 | LOX1 | | 2.44 |
| Growth/differentiation factor 2 | GDF2 | | 2.30 |
| Tumor necrosis factor receptor superfamily member 10A | TNFRSF10A | | 2.08 |
| Fibronectin | FN | | 2.06 |
| Protein AMBP | AMBP | | 2.05 |
| Follistatin | FS | | 2.02 |

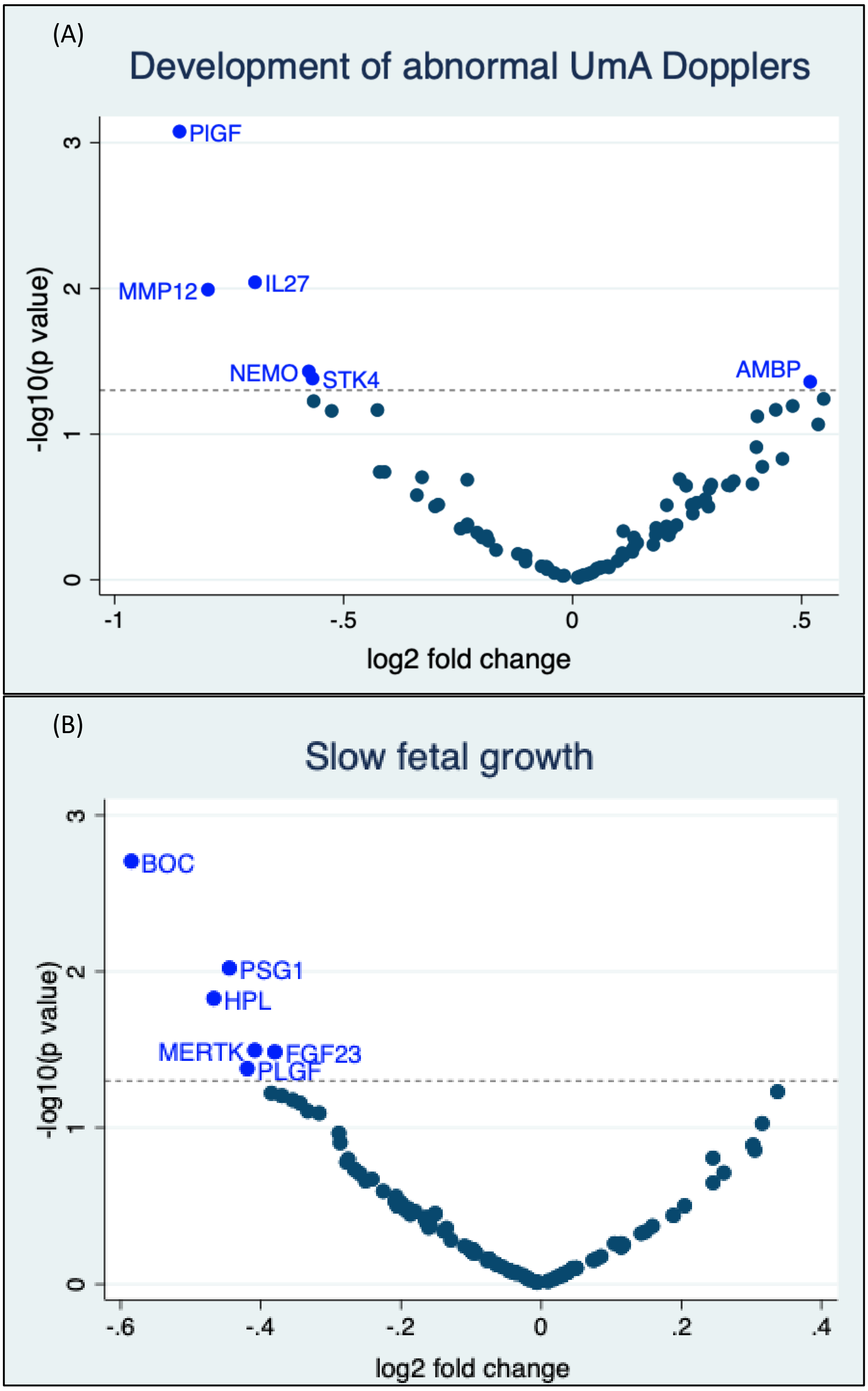

Figure 10: Volcano plots showing the statistical significance and magnitude of associations between the centred and scaled concentrations of the 93 proteins from the discovery and validation sets and (A) the development of abnormal umbilical artery (UmA) Dopplers (pulsatility index >95$^{th}$ centile) (B) slow fetal growth (a worsening of weight deviation of $\geq$10 percentage points over a two-week interval). Associations tested with 2-sided t tests. Dotted line indicates p=0.05. BOC=brother of cysteine dioxygenase, FGF23=fibroblast growth factor 23, HPL=human placental lactogen, PlGF=placental growth factor, PSG1=pregnancy-specific beta-glycoprotein 1, IL27=interleukin 27, MERTK=tyrosine-protein kinase Mer, MMP12=matrix metalloproteinase-12, NEMO=NF-kappa-B essential modulator, STK4=serine/threonine-protein kinase 4, AMBP=protein AMBP.

Table 12: Top three three-variable and top two-variable models for predicting fetal or neonatal death, death or delivery ≤28+0 weeks of gestation and the development of abnormal umbilical artery (UmA) Dopplers from the combined data set. Leave-one-out cross-validated models generated using centred and scaled values and evaluated on the basis of geometric mean of receiver operating characteristic curve area under the curve (AUC), precision-recall ROC area under the curve (PRROC), Matthews correlation coefficient (MCC) and $F_1$ rankings, excluding models with variance inflation factors (VIFs) of five or more (see Methods).

| Outcome | Variables | Proteins | AUC (95% CI) | PRROC | MCC | $F_1$ |
|---|---|---|---|---|---|---|
| Fetal or neonatal death | 3 | PlGF, IL16, ADM | 0.81 (0.72-0.89) | 0.70 | 0.51 | 0.65 |
| | | ADM, IL17D, PAPPA | 0.78 (0.69-0.87) | 0.65 | 0.53 | 0.63 |
| | | ADM, IL17D, PlGF | 0.81 (0.73-0.90) | 0.68 | 0.48 | 0.64 |
| | 2 | PlGF, sflt1 | 0.78 (0.69-0.87) | 0.62 | 0.45 | 0.59 |
| Death or delivery ≤28+0 weeks of gestation | 3 | PlGF, HPL, LOX1 | 0.87 (0.80-0.93) | 0.84 | 0.58 | 0.78 |
| | | PlGF, HPL, MARCO | 0.85 (0.79-0.92) | 0.83 | 0.63 | 0.81 |
| | | PlGF, HPL, ADAMTS13 | 0.86 (0.80-0.93) | 0.85 | 0.50 | 0.75 |
| | 2 | PlGF, PSG1 | 0.84 (0.77-0.92) | 0.80 | 0.57 | 0.78 |
| Development of abnormal UmA Dopplers | 3 | PDGFB, PSGL1, IL18 | 0.79 (0.65-0.94) | 0.75 | 0.60 | 0.78 |
| | | PDGFB, PSGL1, FGF23 | 0.80 (0.66-0.94) | 0.79 | 0.56 | 0.75 |
| | | PDGFB, CTRC, leptin | 0.78 (0.63-0.93) | 0.74 | 0.60 | 0.78 |
| | 2 | PDGFB, PSGL1 | 0.79 (0.64-0.93) | 0.73 | 0.60 | 0.78 |

NOTE: the model containing PlGF and sflt1 is not equivalent to a sflt1:PlGF ratio. ADAMTS13=A disintegrin and metalloproteinase with thrombospondin motifs 13, ADM=pro-adrenomedullin, CTRC=chymotrypsin C, FGF23=fibroblast growth factor 23, HPL=human placental lactogen, LOX1=lectin-like oxidised LDL receptor 1, MARCO=macrophage receptor MARCO, PAPPA=pregnancy-associated plasma protein A, PDGFB=platelet-derived growth factor subunit B, PlGF=placental growth factor, PSG1=pregnancy-specific beta-1 glycoprotein, PSGL1=p-selectin glycoprotein ligand 1, sflt1=soluble fms-like tyrosine kinase 1.

Table 13: Protein and ultrasound measurements at enrolment showing a significant association with gestational age at livebirth or diagnosis of fetal death and/or interval between enrolment and livebirth or diagnosis of fetal death at a Benjamini-Hochberg 1% false discovery rate.

| | Strength of association with gestational age at livebirth or diagnosis of fetal death -log10(p value) | Strength of association with interval between enrolment and livebirth or diagnosis of fetal death or livebirth -log10(p value) |
|---|---|---|
| PlGF | 22.0 | 17.2 |
| HPL | 11.1 | 8.0 |
| UmA category | 9.9 | 11.0 |
| Mean UtA PI | 7.2 | 6.5 |
| Thrombospondin 2 | 7.1 | 5.5 |
| sflt1 | 6.9 | 8.0 |
| MMP12 | 6.1 | 6.8 |
| Decorin | 5.7 | 4.7 |
| IL1RL2 | 5.1 | |
| PSG1 | 5.1 | |
| Growth hormone | 4.2 | |
| Protein AMBP | 4.2 | |

HPL=human placental lactogen, IL1RL2=interleukin-1 receptor-like 2, MMP12=matrix metalloproteinase 12, PlGF=placental growth factor, PSG1=pregnancy specific glycoprotein 1, sflt1=soluble fms-like tyrosine kinase 1, UmA=umbilical artery, UtA PI=uterine artery pulsatility index.

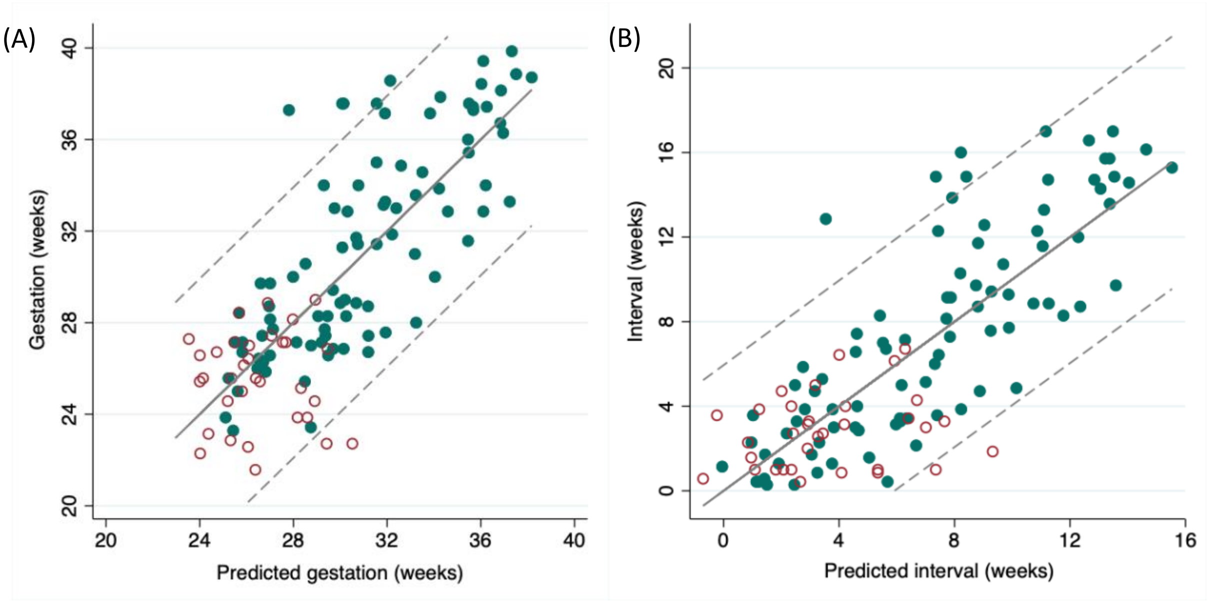

Figure 11: (A) Predicted versus actual gestational age of either livebirth or diagnosis of fetal death, based on the sparser model containing PlGF and sflt1 concentration and umbilical artery Doppler category (B) Predicted versus actual interval from enrolment to either livebirth or diagnosis of fetal death, based on the sparser model containing PlGF and sflt1 concentrations, umbilical artery Doppler category and gestational age at enrolment. Green filled circles=pregnancies ending in livebirth, red hollow circles=pregnancies ending in fetal death, dotted lines=95% prediction intervals.

Table 14: Comparison of the maternal and pregnancy characteristics of participants with and without samples available for placental histological classification.

| Characteristics and outcomes | Sample available (n=55) | No sample available (n=68) | p value |
|---|---|---|---|
| Maternal age (years, mean SD) | 33 (6.7) | 34 (5.9) | 0.18[1] |
| BMI (median IQR) | 24.9 (22.8-29.0) | 26.2 (22.8-30.0) | 0.87[2] |
| Ethnicity (n %) | | | |
| White | 33 (60) | 36 (54) | 0.92[3] |
| Black | 11 (20) | 14 (21) | |
| Asian | 10 (18) | 15 (22) | |
| Other | 1 (2) | 2 (3) | |
| Mean UtA PI >95$^{th}$ centile at enrolment (Schaffer and Staudach 1997) (n %) | 43 (80) | 55 (81) | 0.86[4] |
| Female fetus (n %) | 34 (63) | 26 (41) | 0.016[4] |
| Stillbirth (n %) | 14 (25) | 19 (28) | 0.76[4] |
| Gestational age at delivery (median IQR) | 28+3 (26+5 to 33+0) | 27+6 (26+2 to 34+1) | 0.81[2] |
| Caesarean delivery (n %) | 40 (73) | 39 (58) | 0.10[4] |

PI=pulsatility index, UmA=umbilical artery, UtA=uterine artery. [1]two-sided t test, [2]Wilcoxon rank sum, [3]Fisher's exact test, [4]chi square test. Missing: mean UtA PI for one pregnancy with a placental sample; fetal sex for one stillborn pregnancy with a placental sample; fetal sex for four stillborn pregnancies with no placental samples.

Table 15: Pregnancy outcome by Amsterdam consensus placental classification (Khong et al. 2016) for the 55 pregnancies with available placental histology samples.

| Classification | | Stillbirth | Neonatal death | Neonatal survival |
|---|---|---|---|---|
| Normal | n=10 | 1 | 0 | 9 |
| MVM | n=39 | 12 | 0 | 27 |
| VUE | n=3 | 1 | 1 | 1 |
| Dysmorphic villi | n=2 | 0 | 1 | 1 |
| FVM | n-1 | 0 | 1 | 0 |

FVM=fetal vascular malperfusion, MVM=maternal vascular malperfusion, VUE=villitis of unknown aetiology.

Table 16: Associations between ultrasound measurements and maternal serum protein concentrations at enrolment and subsequent placental classification of (1) any placental pathology (2) maternal vascular malperfusion (MVM) according to the Amsterdam consensus criteria (Khong et al. 2016).

| | | Any placental pathology (n=45) | No placental pathology identified (n=10) | p value | MVM (n=39) | non-MVM (n=16) | p value | AUC (95% CI) |
|---|---|---|---|---|---|---|---|---|
| $EFW_{HM}$ z-score (median IQR) (Hadlock et al. 1985, Marsal et al. 1996) | | -3.0 (-3.4 to -2.6) | -2.8 (-3.4 to -2.0) | 0.44 | -2.9 (-3.3 to -2.6) | -3.2 (-3.4 to -2.6) | 0.63 | |
| $EFW_{Int}$ z-score (median IQR) (Stirnemann et al. 2017) | | -3.4 (-4.3 to -2.6) | -3.0 (-4.9 to -2.6) | 0.72 | -3.4 (-4.2 to -2.6) | -3.4 (-4.9 to -2.6) | 0.78 | |
| UmA category at enrolment (n %) | PI ≤95th centile | 19 (42) | 6 (60) | 0.57# | 18 (46) | 7 (44) | 0.54# | |
| | PI >95th centile with positive EDF | 14 (31) | 1 (10) | | 12 (31) | 3 (19) | | |
| | Absent EDF | 6 (13) | 2 (20) | | 4 (10) | 4 (25) | | |
| | Reversed EDF | 6 (13) | 1 (10) | | 5 (13) | 2 (13) | | |
| Abnormal UmA PI (>95th centile) at any point before delivery (n %) (Schaffer and Staudach 1997) | | 33 (73) | 5 (50) | 0.15 | 28 (72) | 10 (63) | 0.50 | |
| Absent or reversed UmA end-diastolic flow at any point before delivery (n %) | | 27 (60) | 4 (40) | 0.30# | 22 (56) | 9 (56) | 0.99 | |
| Mean UtA PI at enrolment (mean SD)[1] | | 1.70 (0.67) | 1.59 (0.69) | 0.63 | 1.80 (0.63) | 1.40 (0.68) | 0.044 | 0.68 (0.51-0.84) |
| PlGF (pg/ml; median IQR) | | 27.8 (18.0-56.6) | 73.3 (22.6-279.6) | 0.06 | 26.7 (17.9-56.6) | 61.7 (29.8-195.8) | 0.043 | 0.68 (0.52-0.84) |
| HPL (mcg/ml; median IQR) | | 110 (77-141) | 134 (85-186) | 0.49 | 101 (74-140) | 135 (93-182) | 0.24 | |
| PAPPA (NPX; mean SD) | | 0.15 (0.88) | -0.28 (0.71) | 0.16 | 0.23 (0.76) | -0.31 (1.01) | 0.036 | 0.67 (0.51-0.84) |

$EFW_{HM}$=estimated fetal weight calculated using Hadlock 3 formula (Hadlock et al. 1985) with z-score calculated using Marsal reference chart (Marsal et al. 1996), $EFW_{Int}$=estimated fetal weight and z-score calculated using Intergrowth formula and chart (Stirnemann et al. 2017), HPL=human placental lactogen, NPX=normalised protein expression, PAPPA=pregnancy-associated plasma protein A, PI=pulsatility index, PlGF=placental growth factor, UmA=umbilical artery, UtA=uterine artery. PLGF analysed as a natural log for AUC, NPX centred and scaled. n=1 missing from MVM, #Fisher's exact test

### Methods

Equation 1: Hadlock 3 EFW formula (Hadlock et al. 1985)

$$log_{10}EFW = 1.326 - 0.00326 \times AC \times FL + \ 0.0107 \times HC + 0.0438 \times AC + 0.158 \times FL$$

Where AC = abdominal circumference, FL = femur length and HC = head circumference. EFW given in grams with measurements in centimetres.

Equation 2: Marsal EFW z-score (Marsal et al. 1996)

$$z\ score = \frac{EFW - \left(-2.278843 \times 10^{-6} {GA_d}^4 + 1.402168 \times 10^{-3} {GA_d}^3 - 2.008726 \times 10^{-1} {GA_d}^2 + 9.284121 GA_d - 41.25956\right)}{0.12\left(-2.278843 \times 10^{-6} {GA_d}^4 + 1.402168 \times 10^{-3} {GA_d}^3 - 2.008726 \times 10^{-1} {GA_d}^2 + 9.284121 GA_d - 41.25956\right)}$$

Where $GA_d$ = gestational age in days

Equation 3: Percentage weight deviation (Marsal 2009)

$$weight\ deviation = \frac{(EFW - 50th\ centile\ fetal\ weight\ for\ gestation)}{50th\ centile\ fetal\ weight\ for\ gestation} \times 100$$

Where 50$^{th}$ centile fetal weights for gestation are calculated using the formula for Marsal z-score with a z-score of 0.

Equation 4: Intergrowth EFW formula (Stirnemann et al. 2017)

$$\log(EFW) = 5.08482 - 54.06633\ \times \left(\frac{AC}{100}\right)^3 - 95.80076\ \times\ \left(\frac{AC}{100}\right)^3 \times \log\left(\frac{AC}{100}\right) + 3.136370 \times (\frac{HC}{100})$$

Where AC = abdominal circumference and HC = head circumference. EFW given in grams with measurements in centimetres.

Equation 5: Intergrowth z-score formula (Stirnemann et al. 2017)

$$\lambda(GA) = -4.257629 - 2162.234 \times GA^{-2} + 0.0002301829 \times GA^{3}$$
$$\mu(GA) = 4.956737 + 0.0005019687 \times GA^{3} - 0.0001227065 \times GA^{3} \times \log(GA)$$
$$\sigma(GA) = 10^{-4} \times (-6.997171 + 0.057559 \times GA^{3} - 0.01493946 \times GA^{3} \times \log(GA))$$

If λ(GA) = 0

$$z\ score = \sigma(GA)^{-1} \times \log\left(\frac{Y}{\mu(GA)}\right)$$

If λ(GA) ≠ 0

$$z\ score = (\sigma(GA) \times \lambda(GA))^{-1} \times \left(\left(\frac{Y}{\mu(GA)}\right)^{\lambda(GA)} - 1\right)$$

Where λ(GA) = skewness for given gestational age, GA = exact gestational age in weeks, μ(GA) = mean EFW for given gestational age, σ(GA) = coefficient of variation for given gestational age, Y = log(EFW), all logs are natural logs.

Table 17: Full list of proteins measured by the Olink Proseek® Multiplex cardiovascular disease (CVD) II proximity extension assay with reported intra-assay variation. Note PlGF is included in the panel, but the PlGF normalised protein expression (NPX) values were not used for our analysis. Instead, we used PlGF concentration, as measured by Elecsys® electrochemiluminescence immunoassays (Roche Diagnostics).

| Protein name (abbreviation) | UniProt No | Intra-assay variation |
|---|---|---|
| Pro-adrenomedullin (ADM) | P35318 | 13% |
| 2,4-dienoyl-CoA reductase, mitochondrial (DECR1) | Q16698 | 15% |
| A disintegrin and metalloproteinase with thrombospondin motifs 13 (ADAM-TS13) | Q76LX8 | 4.0% |
| Agouti-related protein (AGRP) | O00253 | 8.6% |
| Alpha-L-iduronidase (IDUA) | P35475 | 6.5% |
| Angiopoietin-1 (ANG-1) | Q15389 | 8.8% |
| Angiopoietin-1 receptor (TIE2) | Q02763 | 7.7% |
| Angiotensin-converting enzyme 2 (ACE2) | Q9BYF1 | 7.9% |
| Bone morphogenetic protein 6 (BMP-6) | P22004 | 21% |
| Brother of cysteine dioxygenase (Protein BOC) | Q9BWV1 | 9.6% |
| Carbonic anhydrase 5A, mitochondrial (CA5A) | P35218 | 8.8% |
| Carcinoembryonic antigen related cell adhesion molecule 8 (CEACAM8) | P31997 | 11% |
| Cathepsin L1 (CTSL1) | P07711 | 10% |
| C-C motif chemokine 17 (CCL17) | Q92583 | 12% |
| C-C motif chemokine 3 (CCL3) | P10147 | 8.6% |
| CD40 ligand (CD40-L) | P29965 | 9.1% |
| Chymotrypsin C (CTRC) | Q99895 | 9.8% |
| C-X-C motif chemokine 1 (CXCL1) | P09341 | 9.6% |
| Decorin (DCN) | P07585 | 7.5% |
| Dickkopf-related protein 1 (Dkk-1) | O94907 | 11% |

| | | |
|---|---|---|
| Fatty acid-binding protein, intestinal (FABP2) | P12104 | 8.5% |
| Fibroblast growth factor 21 (FGF-21) | Q9NSA1 | 12% |
| Fibroblast growth factor 23 (FGF-23) | Q9GZV9 | 14% |
| Follistatin (FS) | P19883 | 9.3% |
| Galectin-9 (Gal-9) | O00182 | 5.2% |
| Gastric intrinsic factor (GIF) | P27352 | 11% |
| Gastrotropin (GT) | P51161 | 16% |
| Growth hormone (GH) | P01241 | 7.4% |
| Growth/differentiation factor 2 (GDF-2) | Q9UK05 | 8.8% |
| Heat shock 27 kDa protein (HSP 27) | P04792 | 11% |
| Heme oxygenase 1 (HO-1) | P09601 | 8.1% |
| Hydroxyacid oxidase 1 (HAOX1) | Q9UJM8 | 8.7% |
| Interleukin-1 receptor antagonist protein (IL-1ra) | P18510 | 12% |
| Interleukin-1 receptor-like 2 (IL-1RL2) | Q9HB29 | 10% |
| Interleukin-17D (IL-17D) | Q8TAD2 | 13% |
| Interleukin-18 (IL-18) | Q14116 | 11% |
| Interleukin-27 (IL-27) | Q8NEV9<br>Q14213 | 7.2% |
| Interleukin-4 receptor subunit alpha (IL-4RA) | P24394 | 9.2% |
| Interleukin-6 (IL-6) | P05231 | 8.8% |
| Kidney injury molecule 1 (KIM-1) | Q96D42 | 11% |
| Lactoylglutathione lyase (GLO1) | Q04760 | 8.3% |
| Lectin-like oxidized LDL receptor 1 (LOX-1) | P78380 | 9.0% |
| Leptin (LEP) | P41159 | 6.2% |
| Lipoprotein lipase (LPL) | P06858 | 7.2% |
| Low affinity immunoglobulin gamma Fc region receptor II-b (FCGR2B) | P31994 | 9.1% |
| Lymphotactin (XCL1) | P47992 | 10% |
| Macrophage receptor MARCO (MARCO) | Q9UEW3 | 6.1% |
| Matrix metalloproteinase-12 (MMP-12) | P39900 | 11% |
| Matrix metalloproteinase-7 (MMP-7) | P09237 | 8.5% |
| Melusin (ITGB1BP2) | Q9UKP3 | 11% |
| Natriuretic peptides B (BNP) | P16860 | - |
| NF-kappa-B essential modulator (NEMO) | Q9Y6K9 | 9.1% |
| Osteoclast-associated immunoglobulin-like receptor (hOSCAR) | Q8IYS5 | 4.9% |
| Pappalysin-1 (PAPPA) | Q13219 | 13% |
| Pentraxin-related protein PTX3 (PTX3) | P26022 | 8.1% |
| Placenta growth factor (PlGF) | P49763 | 12% |
| Platelet-derived growth factor subunit B (PDGFB) | P01127 | 11% |
| Poly [ADP-ribose] polymerase 1 (PARP-1) | P09874 | 9.4% |
| Polymeric immunoglobulin receptor (PIgR) | P01833 | 3.4% |
| Programmed cell death 1 ligand 2 (PD-L2) | Q9BQ51 | 8.7% |
| Proheparin-binding EGF-like growth factor (HB-EGF) | Q99075 | 8.4% |
| Pro-interleukin-16 (IL-16) | Q14005 | 11% |
| Prolargin (PRELP) | P51888 | 6.9% |
| Prostasin (PRSS8) | Q16651 | 7.6% |
| Protein AMBP (AMBP) | P02760 | 5.6% |
| Proteinase-activated receptor 1 (PAR-1) | P25116 | 9.4% |
| Protein-glutamine gamma-glutamyltransferase 2 (TGM2) | P21980 | 8.0% |
| Proto-oncogene tyrosine-protein kinase Src (SRC) | P12931 | 9.7% |
| P-selectin glycoprotein ligand 1 (PSGL1) | Q14242 | 5.9% |

| | | |
|---|---|---|
| Receptor for advanced glycosylation end products (RAGE) | Q15109 | 8.8& |
| Renin (REN) | P00797 | 7.8% |
| Serine protease 27 (PRSS27) | Q9BQR3 | 9.3% |
| Serine/threonine-protein kinase 4 (STK4) | Q13043 | 7.2% |
| Serpin A12 (SERPINA12) | Q8IW75 | 10% |
| SLAM family member 5 (CD84) | Q9UIB8 | 9.0% |
| SLAM family member 7 (SLAMF7) | Q9NQ25 | 11% |
| Sortilin (SORT1) | Q99523 | 7.8% |
| Spondin-2 (SPON2) | Q9BUD6 | 5.1% |
| Stem cell factor (SCF) | P21583 | 7.3% |
| Superoxide dismutase [Mn], mitochondrial (SOD2) | P04179 | 5.9% |
| T-cell surface glycoprotein CD4 (CD4) | P01730 | 9.6% |
| Thrombomodulin (TM) | P07204 | 11% |
| Thrombopoietin (THPO) | P40225 | 9.3% |
| Thrombospondin-2 (THBS2) | P35442 | 5.4% |
| Tissue factor (TF) | P13726 | 8.2% |
| TNF-related apoptosis-inducing ligand receptor 2 (TRAIL-R2) | O14763 | 10% |
| Tumor necrosis factor receptor superfamily member 10A (TNFRSF10A) | O00220 | 11% |
| Tumor necrosis factor receptor superfamily member 11A (TNFRSF11A) | Q9Y6Q6 | 10% |
| Tumor necrosis factor receptor superfamily member 13B (TNFRSF13B) | O14836 | 9.5% |
| Tyrosine-protein kinase Mer (MERTK) | Q12866 | 10% |
| Vascular endothelial growth factor D (VEGF-D) | O43915 | 7.2% |
| V-set and immunoglobulin domain-containing protein 2 (VSIG2) | Q96IQ7 | 8.2% |

Table 18: The scoring system for ranking proteins identified through pooled mass spectrometry, using the following formula:

$$Score = \sum_{i=1}^{4} r_i v_i + t + p + u$$

| Category | Criteria | Score |
|---|---|---|
| Ratio score (r) | >1.33 or <0.75 | 5 |
| | >1.5 or <0.67 | 10 |
| | >2 or <0.5 | 15 |
| | >4 or <0.25 | 20 |
| Variability score (v) | >50% | 0 |
| | 31 to 50% | 1 |
| | 16 to 30% | 1.34 |
| | ≤15% | 1.50 |
| Trend score (t) | 2 ratios both >1.2 or both <0.83 | 5 |
| | 3 ratios all >1.2 or all <0.83 | 10 |
| | 4 ratios all >1.2 or all <0.83 | 20 |
| Peptide score (p) | 1 | -50 |
| | 2-4 | 0 |
| | ≥5 | 5 |
| Ubiquity score (u) | One of the 12 depleted proteins | -50 |
| | Haemoglobin protein | -20 |

Table 19: Top 50 highest scoring proteins identified from pooled serum liquid chromatography and tandem mass spectrometry, based on the scoring system above.

| UniProt code | Description |
|---|---|
| P02751 | Fibronectin OS=Homo sapiens GN=FN1 PE=1 SV=4 - [FINC_HUMAN] |
| P02741 | C-reactive protein OS=Homo sapiens GN=CRP PE=1 SV=1 - [CRP_HUMAN] |
| P0DJI9 | Serum amyloid A-2 protein OS=Homo sapiens GN=SAA2 PE=1 SV=1 - [SAA2_HUMAN] |
| P20848 | Putative alpha-1-antitrypsin-related protein OS=Homo sapiens GN=SERPINA2P PE=5 SV=1 - [A1ATR_HUMAN] |
| P0DML2 | Chorionic somatomammotropin hormone 1 OS=Homo sapiens GN=CSH1 PE=1 SV=1 - [CSH1_HUMAN] |
| P11464 | Pregnancy-specific beta-1-glycoprotein 1 OS=Homo sapiens GN=PSG1 PE=2 SV=1 - [PSG1_HUMAN] |
| Q13046 | Putative pregnancy-specific beta-1-glycoprotein 7 OS=Homo sapiens GN=PSG7 PE=5 SV=2 - [PSG7_HUMAN] |
| Q00887 | Pregnancy-specific beta-1-glycoprotein 9 OS=Homo sapiens GN=PSG9 PE=2 SV=2 - [PSG9_HUMAN] |
| P01591 | Immunoglobulin J chain OS=Homo sapiens GN=IGJ PE=1 SV=4 - [IGJ_HUMAN] |
| P0DJI8 | Serum amyloid A-1 protein OS=Homo sapiens GN=SAA1 PE=1 SV=1 - [SAA1_HUMAN] |
| P01834 | Ig kappa chain C region OS=Homo sapiens GN=IGKC PE=1 SV=1 - [IGKC_HUMAN] |
| Q6N069 | N-alpha-acetyltransferase 16, NatA auxiliary subunit OS=Homo sapiens GN=NAA16 PE=1 SV=2 - [NAA16_HUMAN] |
| Q8TDD5 | Mucolipin-3 OS=Homo sapiens GN=MCOLN3 PE=1 SV=1 - [MCLN3_HUMAN] |
| Q9UQ72 | Pregnancy-specific beta-1-glycoprotein 11 OS=Homo sapiens GN=PSG11 PE=2 SV=3 - [PSG11_HUMAN] |
| Q16557 | Pregnancy-specific beta-1-glycoprotein 3 OS=Homo sapiens GN=PSG3 PE=2 SV=2 - [PSG3_HUMAN] |
| Q00888 | Pregnancy-specific beta-1-glycoprotein 4 OS=Homo sapiens GN=PSG4 PE=2 SV=3 - [PSG4_HUMAN] |
| P02760 | Protein AMBP OS=Homo sapiens GN=AMBP PE=1 SV=1 - [AMBP_HUMAN] |
| Q9UIQ6 | Leucyl-cystinyl aminopeptidase OS=Homo sapiens GN=LNPEP PE=1 SV=3 - [LCAP_HUMAN] |
| P0CG05 | Ig lambda-2 chain C regions OS=Homo sapiens GN=IGLC2 PE=1 SV=1 - [LAC2_HUMAN] |
| P55196 | Afadin OS=Homo sapiens GN=MLLT4 PE=1 SV=3 - [AFAD_HUMAN] |
| P04196 | Histidine-rich glycoprotein OS=Homo sapiens GN=HRG PE=1 SV=1 - [HRG_HUMAN] |
| P04275 | von Willebrand factor OS=Homo sapiens GN=VWF PE=1 SV=4 - [VWF_HUMAN] |
| Q06033 | Inter-alpha-trypsin inhibitor heavy chain H3 OS=Homo sapiens GN=ITIH3 PE=1 SV=2 - [ITIH3_HUMAN] |
| Q13219 | Pappalysin-1 OS=Homo sapiens GN=PAPPA PE=1 SV=3 - [PAPP1_HUMAN] |
| O95445 | Apolipoprotein M OS=Homo sapiens GN=APOM PE=1 SV=2 - [APOM_HUMAN] |
| Q9UN37 | Vacuolar protein sorting-associated protein 4A OS=Homo sapiens GN=VPS4A PE=1 SV=1 - [VPS4A_HUMAN] |
| P11226 | Mannose-binding protein C OS=Homo sapiens GN=MBL2 PE=1 SV=2 - [MBL2_HUMAN] |
| O75636 | Ficolin-3 OS=Homo sapiens GN=FCN3 PE=1 SV=2 - [FCN3_HUMAN] |
| P48740 | Mannan-binding lectin serine protease 1 OS=Homo sapiens GN=MASP1 PE=1 SV=3 - [MASP1_HUMAN] |
| O14791 | Apolipoprotein L1 OS=Homo sapiens GN=APOL1 PE=1 SV=5 - [APOL1_HUMAN] |
| O75882 | Attractin OS=Homo sapiens GN=ATRN PE=1 SV=2 - [ATRN_HUMAN] |

| | |
|---|---|
| P00450 | Ceruloplasmin OS=Homo sapiens GN=CP PE=1 SV=1 - [CERU_HUMAN] |
| P00734 | Prothrombin OS=Homo sapiens GN=F2 PE=1 SV=2 - [THRB_HUMAN] |
| P00736 | Complement C1r subcomponent OS=Homo sapiens GN=C1R PE=1 SV=2 - [C1R_HUMAN] |
| P00740 | Coagulation factor IX OS=Homo sapiens GN=F9 PE=1 SV=2 - [FA9_HUMAN] |
| P00742 | Coagulation factor X OS=Homo sapiens GN=F10 PE=1 SV=2 - [FA10_HUMAN] |
| P00747 | Plasminogen OS=Homo sapiens GN=PLG PE=1 SV=2 - [PLMN_HUMAN] |
| P00748 | Coagulation factor XII OS=Homo sapiens GN=F12 PE=1 SV=3 - [FA12_HUMAN] |
| P00751 | Complement factor B OS=Homo sapiens GN=CFB PE=1 SV=2 - [CFAB_HUMAN] |
| P01008 | Antithrombin-III OS=Homo sapiens GN=SERPINC1 PE=1 SV=1 - [ANT3_HUMAN] |
| P01011 | Alpha-1-antichymotrypsin OS=Homo sapiens GN=SERPINA3 PE=1 SV=2 - [AACT_HUMAN] |
| P01019 | Angiotensinogen OS=Homo sapiens GN=AGT PE=1 SV=1 - [ANGT_HUMAN] |
| P01024 | Complement C3 OS=Homo sapiens GN=C3 PE=1 SV=2 - [CO3_HUMAN] |
| P01031 | Complement C5 OS=Homo sapiens GN=C5 PE=1 SV=4 - [CO5_HUMAN] |
| P01042 | Kininogen-1 OS=Homo sapiens GN=KNG1 PE=1 SV=2 - [KNG1_HUMAN] |
| P02649 | Apolipoprotein E OS=Homo sapiens GN=APOE PE=1 SV=1 - [APOE_HUMAN] |
| P02654 | Apolipoprotein C-I OS=Homo sapiens GN=APOC1 PE=1 SV=1 - [APOC1_HUMAN] |
| P02655 | Apolipoprotein C-II OS=Homo sapiens GN=APOC2 PE=1 SV=1 - [APOC2_HUMAN] |
| P02656 | Apolipoprotein C-III OS=Homo sapiens GN=APOC3 PE=1 SV=1 - [APOC3_HUMAN] |
| P02743 | Serum amyloid P-component OS=Homo sapiens GN=APCS PE=1 SV=2 - [SAMP_HUMAN] |

Table 20: Combinations of proteins included in different sections of the study

| Platform | Protein(s) | Tested in discovery set | Univariate association with outcomes in discovery set | Model generation from discovery set | Parenclitic network analysis | Tested in validation set | Univariate association with outcomes in combined set |
|---|---|---|---|---|---|---|---|
| Roche Elecsys | PlGF & sflt1 | ✓ | ✓ | ✓ | ✓ | ✓ | ✓ |
| ELISA | VEGF-A | ✓ | | | ✓ | | |
| | VEGF-D, endoglin, NP1, VEGFR2 | ✓ | ✓ | ✓ | ✓ | | |
| | FN, HPL, PSG1 | ✓ | ✓ | ✓ | ✓ | ✓ | ✓ |
| | LNPEP & SAA | ✓ | ✓ | ✓ | ✓ | | |
| Olink multiplex | PlGF | ✓ | | | | ✓ | |
| | VEGF-D | ✓ | | | | ✓ | ✓ |
| | BNP, melusin, PARP1 | ✓ | | | ✓ | ✓ | |
| | Remaining 87 proteins | ✓ | ✓ | ✓ | ✓ | ✓ | ✓ |
| Totals | | | 98 | 98 | 102 | | 93 |
